# Extended genome-wide association study employing the African Genome Resources Panel identifies novel susceptibility loci for Alzheimer’s Disease in individuals of African ancestry

**DOI:** 10.1101/2023.08.29.23294774

**Authors:** Nicholas R. Ray, Brian W. Kunkle, Kara Hamilton-Nelson, Jiji T. Kurup, Farid Rajabli, Mehmet I. Cosacak, Caghan Kizil, Melissa Jean-Francois, Michael Cuccaro, Dolly Reyes-Dumeyer, Laura Cantwell, Amanda Kuzma, Jeffery M. Vance, Sujuan Gao, Hugh C. Hendrie, Olusegun Baiyewu, Adesola Ogunniyi, Rufus O. Akinyemi, Alzheimer’s Disease Genetics Consortium, Wan-Ping Lee, Eden R. Martin, Li-San Wang, Gary W. Beecham, William S. Bush, Lindsay A. Farrer, Jonathan L. Haines, Goldie S. Byrd, Gerard D. Schellenberg, Richard Mayeux, Margaret A. Pericak-Vance, Christiane Reitz

## Abstract

**INTRODUCTION:** Despite a two-fold increased risk, individuals of African ancestry have been significantly underrepresented in Alzheimer’s Disease (AD) genomics efforts.

**METHODS:** GWAS of 2,903 AD cases and 6,265 cognitive controls of African ancestry. Within-dataset results were meta-analyzed, followed by gene-based and pathway analyses, and analysis of RNAseq and whole-genome sequencing data.

**RESULTS:** A novel AD risk locus was identified in *MPDZ* on chromosome 9p23 (rs141610415, MAF=.002, *P*=3.68×10^−9^). Two additional novel common and nine novel rare loci approached genome-wide significance at *P*<9×10^−7^. Comparison of association and LD patterns between datasets with higher and lower degrees of African ancestry showed differential association patterns at chr12q23.2 (*ASCL1*), suggesting that the association is modulated by regional origin of local African ancestry.

**DISCUSSION:** Increased sample sizes and sample sets from Africa covering as much African genetic diversity as possible will be critical to identify additional disease-associated loci and improve deconvolution of local genetic ancestry effects.

## 1. INTRODUCTION

Based on estimations by the World Health Organization (WHO), approximately 55 million people globally suffer from dementia, and forty million of these cases are thought to be due to Alzheimer’s disease (AD). As the proportion of older individuals increases in nearly every country, the number of AD cases is expected to rise exponentially to ∼78 million in 2030 and ∼139 million by 2050 (https://www.who.int/news-room/fact-sheets/detail/dementia), making identification of underlying factors one of the most urgent global public health concerns.

Recent large-scale genome-wide association studies (GWAS) by our group and others identified over 75 loci associated with risk of AD and related dementias[1–5]. While these findings have significantly advanced the field by providing invaluable insights into underlying mechanistic pathways, AD genomic studies were almost exclusively conducted in individuals of European ancestry. This limits our ability to identify ancestry-specific causative genetic variants, loci, and mechanistic pathways underlying the disease, and substantially hampers our progress towards identification of effective druggable targets to address this devastating disease in underrepresented populations.

With a two-fold increased disease risk compared to non-Hispanic Whites[6] (NHW), a ∼64% higher rate of progression to AD and related dementias (ADRD) compared to NHW[7, 8], a higher degrees of dementia risk factors, and greater cognitive impairment and neuropsychiatric symptom severity[9], individuals of African ancestry are at a particularly high risk of AD and its sub-phenotypes. To identify genetic risk factors associated with AD in African Americans, we previously conducted GWAS in 8,006 subjects of African American ancestry and identified 16 common and rare loci associated with AD, most of which appear ancestry-specific[10, 11]. To identify additional risk variants, genes, and mechanistic pathways, we reanalyzed these data with a 14.5% increase in sample size. To start to deconvolute regional African ancestry effects we included West African individuals with a high degrees of African ancestry. Analysis of RNAseq and whole-genome sequencing data was performed for the most interesting loci. These analyses identified 12 novel susceptibility loci potentially associated with AD in individuals of African ancestry that have not been identified in other ancestry groups.

## 2. METHODS

### 2.1 Samples

To identify additional risk loci associated with AD in individuals of African ancestry, we both increased the sample size by 14.5% in this updated analysis and included, in addition to African Americans, ∼800 individuals from West Africa, yielding in total 9,168 subjects of African ancestry in the analysis (2,903 cases, 6,265 controls). Summary demographics of all datasets are shown in **Supplementary Table 1** and additional detailed information about each dataset is provided in the **Description of Cohorts** in the **Supplementary Materials**. Written informed consent was obtained from all participants or, for those with cognitive impairment, from a caregiver, legal guardian, or other proxy. Study protocols for all cohorts were reviewed and approved by the appropriate institutional review boards.

### 2.2 Diagnosis of AD and age of onset

Participants were diagnosed for AD according to the National Institute of Neurological and Communicative Disorders and Stroke–Alzheimer’s Disease and Related Disorders Association criteria[12, 13]. Age at onset for AD patients and age at examination or death for healthy controls was available for most datasets. When not available, other information was used instead, such as age at diagnosis (Chicago Health and Aging Project [CHAP], Minority Aging Research Study/Clinical Minority Core [MARS/CORE]) or age at ascertainment (Indiana University). To restrict the analyses to cases with late-onset AD, individuals with age < 60 years at symptom onset, last examination, or death were excluded.

### 2.3 Genotyping

The platforms used for genome-wide genotyping in the individual datasets are shown in **Supplementary Table 2**. For all datasets, samples were randomly plated to minimize potential batch effects.

### 2.4 *APOE* genotyping

For the Alzheimer Disease Centers, Adult Changes in Thought, National Institute in Aging– LOAD/National Cell Repository for Alzheimer Disease (NIA-FBS/NCRAD), UM/VU, CHAP, Columbia University, Mayo Clinic, and REAAADI cohorts, *APOE* genotypes were based on haplotypes derived from single-nucleotide polymorphisms SNPs rs7412 and rs429358. Whole-genome sequencing genotypes of these two SNPs were used for the Ibadan cohort. For the MIRAGE and GenerAAtions cohorts, *APOE* genotypes were determined using the Roche Diagnostics LightCycler 480 instrument (Roche Diagnostics) and LightMix Kit ApoE C112R R158 (TIB MOLBIOL); for the University of Pittsburgh, Washington Heights Columbia Aging Project, and Indianapolis cohorts, they were determined by pyrosequencing or analysis of restriction fragment length polymorphisms; for the Religious Orders Study/Rush Memory and Aging Project (ROS/MAP) and MARS/CORE they were determined by high-throughput sequencing of codons 112 and 158 in *APOE* by Agencourt Bioscience Corporation; for the Washington University samples they were determined using a taqman-based assay from Applied Biosystems.

### 2.5 Genotype quality control

Standard quality control was performed separately on each dataset’s genotype and sample-level data. Single-nucleotide polymorphisms with call rates less than 98% or not in Hardy-Weinberg equilibrium at *P* < 10^−6^ in controls were excluded. In addition, subjects with non-African American ancestry according to principal components (PCs) analysis of ancestry informative markers, and study participants whose reported sex did not equal the sex designation determined by analysis of the X-chromosome SNPs were removed. We identified latent relatedness among participants using the estimated proportion of alleles (π) shared identical by descent (IBD) and included from each duplicate pair (π > 0.95) or relative pair (0.4 ≤ π < 0.95) one participant prioritizing first samples with non-missing disease status followed by samples with higher SNP call rate. Relationships among individuals in family-based cohorts (MIRAGE) were validated employing pairwise genome-wide estimates of IBD allele sharing.

### 2.6 Genotype imputation

Each dataset was independently phased and imputed to the African Genome Resource (AGR) reference panel utilizing the Sanger genotype imputation and phasing service (https://imputation.sanger.ac.uk/), which employs EAGLE2 for phasing and PBWT (Positional Burrows-Wheeler Transform) for genotype imputation. The AGR reference panel provides information on 93,421,145 autosomal bi-allelic markers and is based on 4,956 samples that includes – in addition to all of the African and non-African populations from the 1000 Genomes Phase 3 reference panel – also ∼2000 samples from Uganda (Baganda, Banyarwanda, Barundi and others) and ∼100 samples each from Ethiopia (Gumuz, Wolayta, Amhara, Oromo, Somali), Egypt, Namibia (Nama/Khoesan), and South Africa (Zulu). Data were filtered to exclude common variants (MAF ≥ 0.01) with imputation quality score < 0.4, rare variants (MAF < 0.01) with imputation quality < 0.7, and variants present in less than 30% of AD cases and 30% of controls. The final SNP set for analysis included 33,089,606 genotyped and imputed variants.

### 2.7 Association analysis

Genome-wide single variant association analyses of common and rare variants were performed individually on each dataset using SNPTEST[14–16]. Age, sex, and population stratification (as determined by Principal Components Analysis calculated individually on each dataset; PCs) were entered as covariates in Model 1; APOEe4 allele dosage (coded as 0,1,2) was entered as an additional covariate in Model 2. Logistic regression was used for case-control datasets and generalized estimating equations (GEE) as implemented in GWAF[17] were used for family-based datasets (i.e. MIRAGE). Associations with extreme beta coefficients (|β| > 5) were filtered out. Within-study results were subsequently meta-analyzed with METAL[18] employing an inverse-variance based model with genomic control. Variants showing significant heterogeneity between studies (*I^2^* > 75%) were removed. The GenABEL package[19] was used to estimate genomic inflation (λ).

### 2.8 Gene-based analysis

Genome-wide gene-based analyses were performed employing MAGMA implemented in the FUMA software[20, 21]. Setting a 35kb window upstream and a 10kb window downstream of the genes and including only variants with MAF > .001 and present in >30% of cases and controls, these analyses were first adjusted for PCs, age, and sex and subsequently in addition for APOEe4 allele dosage. Genome-wide significance was determined using Bonferroni correction.

### 2.9 Pathway analysis

Pathway analyses were performed with MAGMA[20], which performs SNP-wise gene analysis of summary statistics with correction for LD between variants and genes to test whether sets of genes are jointly associated with a phenotype (i.e. AD), compared to other genes across the genome. 9,988 gene-sets from GO[22] pathways were used in the analyses. These analyses were performed using the same parameters as described above in the gene-based analysis section.

### 2.10 Local ancestry

To estimate local ancestry within African American datasets, we initially merged each array dataset with the Human Genome Diversity Project (HGDP) reference panel individually, utilizing PLINK v2 software[23, 24]. This integration involved 98 African and 109 European individuals from the HGDP reference populations. Subsequently, the merged datasets were phased employing the SHAPEIT tool version 2 with default configurations using the 1000 Genomes Phase 3 reference panel[25, 26]. Finally, we inferred the local ancestries using the discriminative modeling approach implemented in RFMix with the PopPhased option and a minimum node size of 5[27].

### 2.11 Bulk RNA sequence analysis in zebrafish

Amyloid toxicity was induced as previously described[28, 29] in the adult telencephalon zebrafish brain. At three days after cerebroventricular injection, the brains were dissected and deep sequencing for bulk RNA was performed[28]. Data can be accessed at GSE74326 at GEO (https://www.ncbi.nlm.nih.gov/geo).

## 3. RESULTS

### 3.1 Single-variant meta-analysis

The results of the single-variant meta-analyses are summarized in **Table 1** and **Figure 1**. Single marker analyses identified one novel genome-wide significant disease-associated locus, and eleven novel loci approaching genome-wide significance at P ≤ 9×10^−7^. The novel genome-wide significant locus, associated with a rare variant, is located within *MPDZ* on chromosome 9p23 (rs141610415, MAF = 0.002, *P* = 3.68×10^−9^) and has strong regional support by variants in LD (**Supplementary Figure 1**). The eleven novel loci approaching genome-wide significance include two common loci centered at 2p25.3 at *LINC01250/TSSC1* and 3p25 at *SRGAP3*, and nine rare loci centered at 2p25 (*KIDINS220*), 2q22 (*TEX41*), 4q22 (*UNC5C*), 6q21 (*IYD/PLEKHG1*), 7p22 (*SDK1*), 8q21 (*MMP16*), 12q23 (*ASCL1*), 16q23 (*CNTNAP4*), and 17q23 (*TANC2*) (see **Table 1**). Eight of these eleven loci have strong regional support by variants in LD (see **Supplementary Figure 1**) and all have consistent directions of effect across most individual datasets (see **Supplementary Figure 2**). There was no genomic inflation in either model (Model 1: λ = 0.95; Model 2: λ = 0.96; see **Supplementary Figure 3** for QQ-plots). Besides US-based datasets of African Americans, the present analyses also included samples of Yoruba from Ibadan with a high degrees of African ancestry (see **Supplementary Figure 4** for principal component plots of all datasets). Comparison of effect sizes of identified loci between the datasets with high (ie. Ibadan dataset) and lower degrees of African ancestry (all other (US-based African American) datasets), showed comparable effects between datasets with higher and lower degrees of African ancestry at all loci except for chr2:3070309, chr2:145902343, and chr12:103570373. These three loci showed a higher effect size in the Yoruba samples (see **Supplementary Figure 2**), indicating that degrees of African ancestry at these loci may modify the effect. Closer examination of the local association and LD patterns in these three regions showed comparable association and LD patterns between African and African American datasets at the two chr2 loci, but differential association and LD patterns at the chr12 locus (see **Supplementary Figure 5**), suggesting that this locus may show differential genetic ancestry. While in the African American datasets the strongest association at the chromosome 12 locus is observed at 103,538kb, the high African ancestry Ibadan dataset shows genome-wide significant association at *P* = 3.8 x 10^−8^ (rs547590324) 180kb upstream within the *ASCL1* gene. Analysis of local ancestry at the two meta-analysis top locus markers from both models (Model 1: rs556001137; Model 2: rs138206541; see **Table 1**) showed that both markers appear on African and Amerindian but not European local ancestry backgrounds (**Supplementary table 3**). Comparison of the allele frequencies of the three top markers only observed with significant association in the African Ibadan dataset (rs547590324, rs144730555, rs148789350; see **Supplementary Figure 5C**) across the African American and Ibadan datasets, as well as gnomAD, 1000 Genomes and the Human Genome Diversity Project (HGDP), suggest that these markers are present in West Africans but less frequent in Hispanics and virtually absent in other African regions (ie. Kenya, South Africa), Europeans, and Asians (https://gnomad.broadinstitute.org/variant/12-102960732-G-A?dataset=gnomad_r3; https://gnomad.broadinstitute.org/variant/12-102957658-C-G?dataset=gnomad_r3, https://gnomad.broadinstitute.org/variant/12-102956366-C-A?dataset=gnomad_r3).

**Figure 1.**
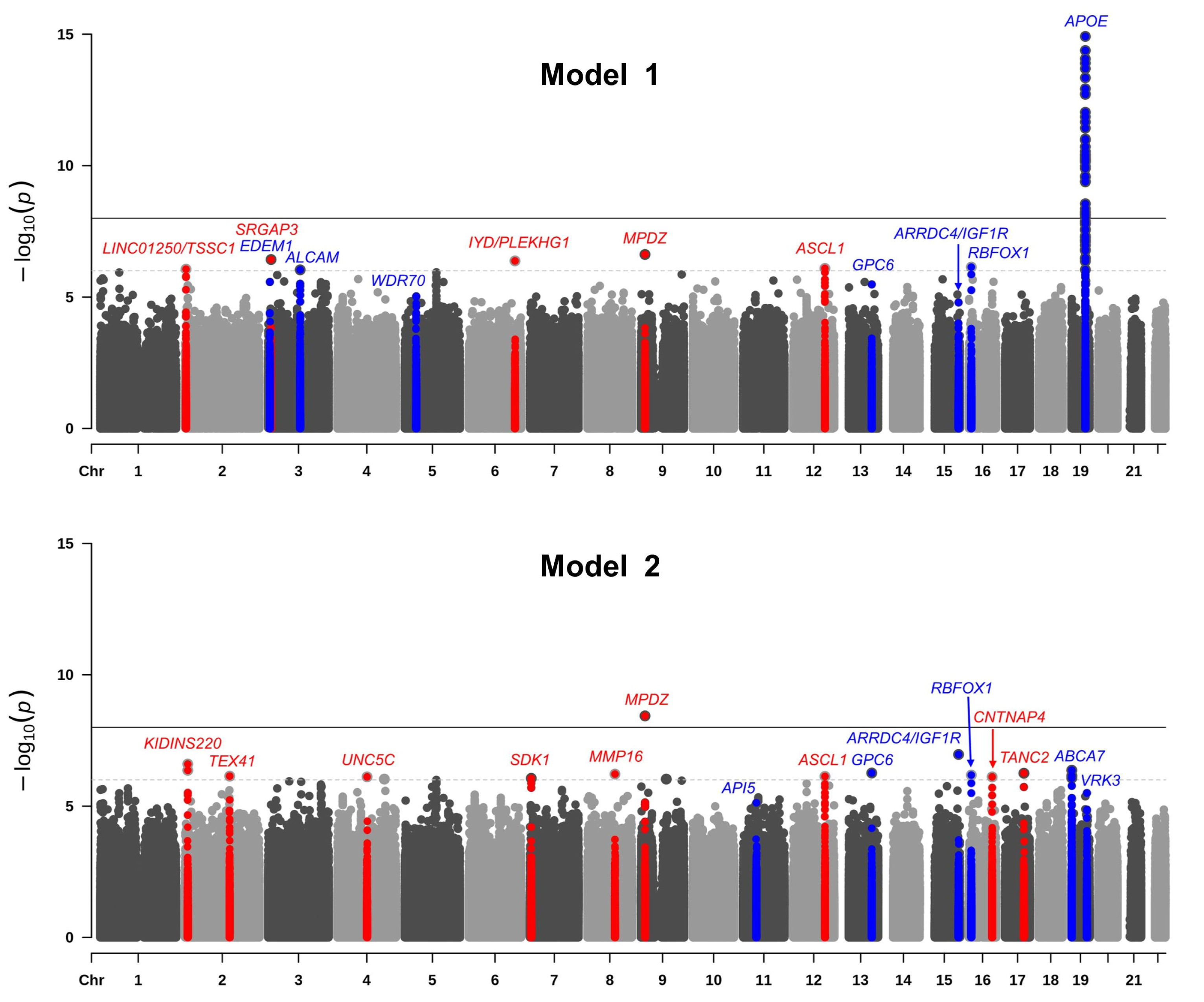
Manhattan plots showing negative log_10_-transformed *P* values from the single-variant meta-analysis adjusted for age, sex, and population stratification (Model 1) and age, sex, population stratification, and *APOE* (Model 2).The solid black horizontal line represents a genome-wide significance threshold of *P* = 5 × 10^−8^ and the dotted line represents a suggestive threshold of threshold of *P* = 5 × 10^−6^. All novel loci signficant at *P* < 9 × 10^−7^ are shown in red and all previously reported loci significant at *P* < 9 × 10^−6^ are shown in blue. The y-axis has been truncated, the lowest *P* value on chromosome 19 is 3.11 × 10^−65^.

**Table 1.**
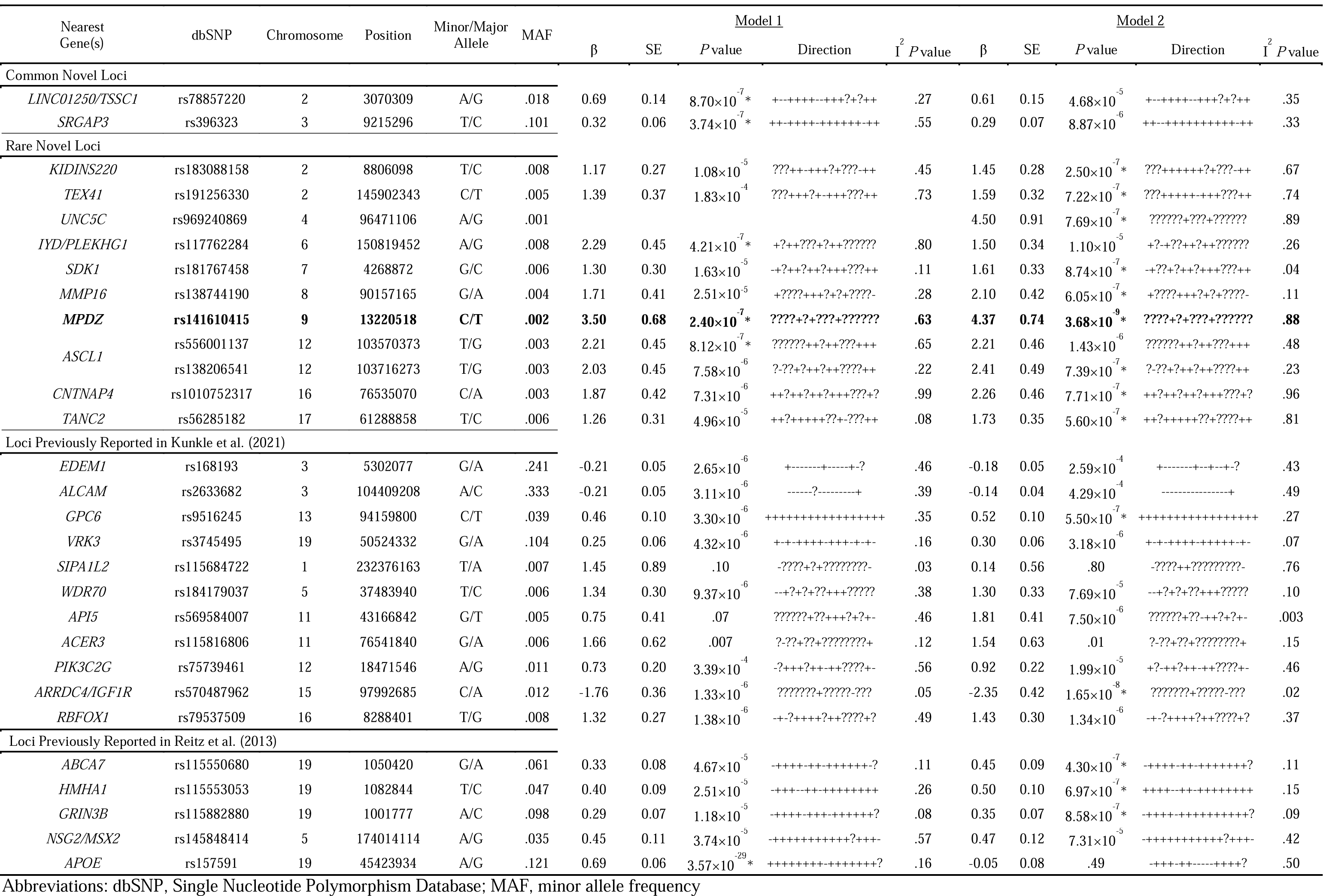
Results of Single-variant Meta-analysis.

Of the 16 loci identified in our previous analyses[10, 11] all but two (*SIPA1L2* and *ACER3,* both associated with rare variants) replicated at a P-value of ≤ 9 × 10^−5^. Of the variants previously implicated in either AD, ADRD, or AD by proxy in African Americans by other studies[30–35], rs112404845 in *COBL* (*P* = 6.13×10^−5^), rs2234258 in *TREM2* (*P* = 1.74×10^−5^), and rs73505251 in *ABCA7* (*P* = 2.13×10^−5^) showed suggestive significance, and variants in *SLC4A1AP* (rs17006206), *POLN* (rs1923775), *RP11-785H20.1* (rs956225), *RP11-116D17.2* (rs10850408), *ENOX1* (rs17460623), *AKAP9* (rs144662445; rs149979685), *TREM2* (rs2234256; rs73427293), *BZRAP1-AS1* (rs263251), *AC010967.2/SCARNA16* (rs58443395), *RP11-157D6.1/CD2AP* (rs7738720), and *RP11-192P9.1/TRPS1* (rs76427927) were replicated with nominal significance (see **Supplementary Table 4**). Of the GWAS loci implicated in NHW individuals[1, 3] besides *APOE* and *ABCA7*, only the variants in *BIN1*, *IDUA*, *UNC5CL*, *UMAD1*, *USP6NL*, *FERMT2*, and *SCIMP* showed nominal association (see **Supplementary Table 5**).

### 3.2 Gene-based and pathway analyses

Gene-based analyses identified two novel loci (*C9orf139* and *SNX31*) that are significantly associated with AD in individuals of African ancestry at a significance threshold of *P* < 9×10^−5^ (see **Table 2** and **Supplementary Figures 6 and 7**), in addition to five regions (*APOE*, *LARP1B*, *TREM2*, *SERPINB13*, and *ABCA7*) that we previously reported[10, 11]. Gene-based results for all previously reported genes in NHW[1, 3] and African ancestry[30–35] are reported in **Supplementary Tables 6 and 7**. Gene-set analyses identified 16 pathways at *P* < 5 x 10^−4^ (see **Table 3**). Notably, besides lipid metabolism, immune response, transcription/DNA repair and intracellular trafficking that are also associated with AD in NHW[1, 3], sodium transport emerged as a novel prominent pathway that has not been reported before in analyses restricted to individuals of African ancestry or in analyses in other ethnic groups.

**Table 2.**
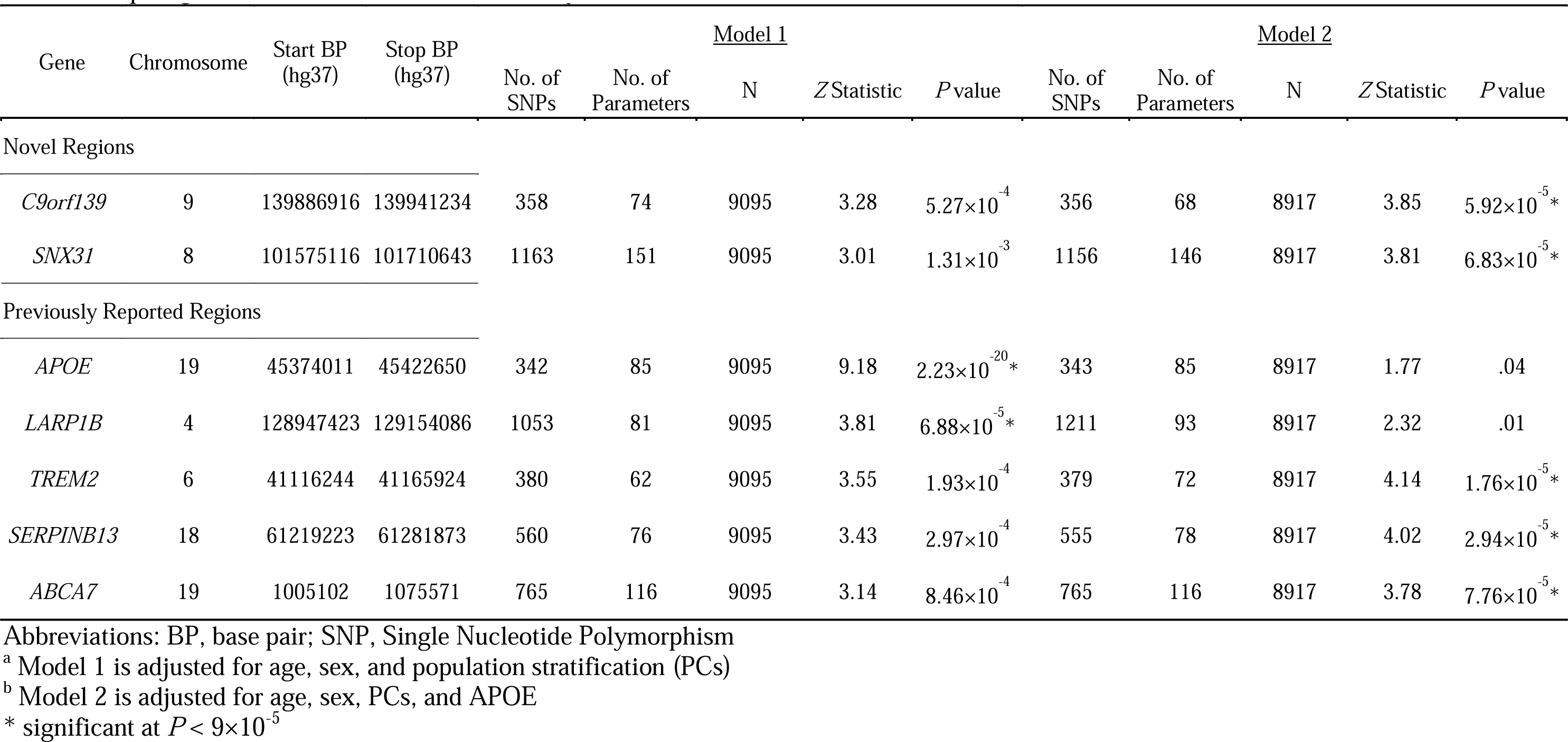
Top Regions Identified in Gene-Based Analysis.

**Table 3.**
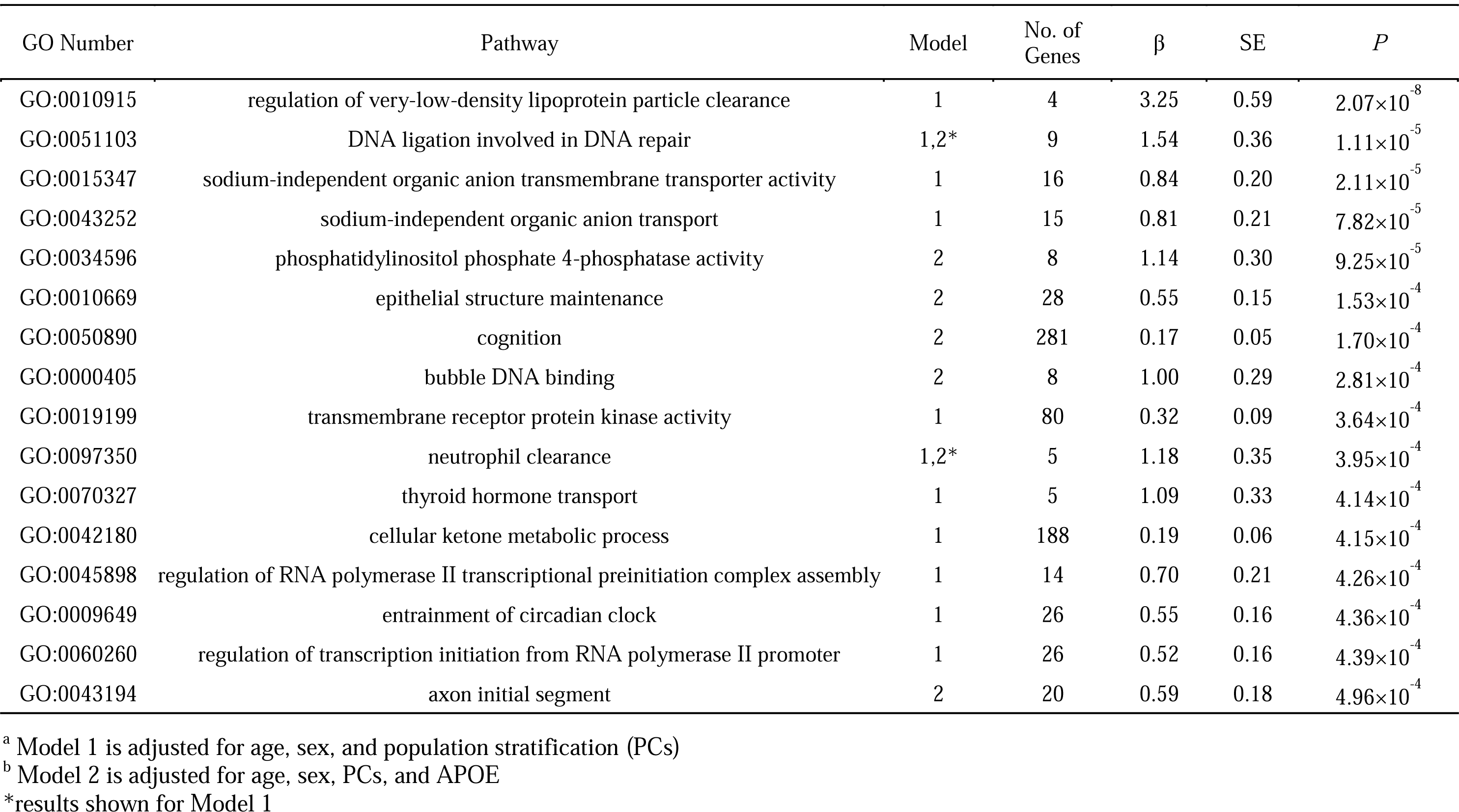
Results of Pathway Analysis.

### 3.3 RNAseq analyses from Human and zebrafish brain tissue

In brain RNAseq data from over 2,100 samples from post-mortem brains of more than 1,100 individuals from the ROSMAP study, the Mount Sinai Brain Bank (MSBB) and the Mayo Clinic (https://agora.adknowledgeportal.org/), all nearest genes at all novel loci identified in single marker association or gene-based analyses are differentially expressed between AD cases and controls (see **Supplementary Figure 8**). In brain RNA-seq analyses in zebrafish, orthologs for *MPDZ, SRGAP3, SDK1, TANC2, MMP16* and *UNC5C* were downregulated after inducing amyloid toxicity, while the ortholog of *ASCL1* was upregulated (see **Supplementary Figure 9**).

### 3.4 Analysis of whole-genome sequencing data at top loci

To identify potential causal variants we analyzed whole-genome sequence data from the ADSP in 1Mb regions around the novel variants listed in Table 1. This analysis revealed nine missense and one stop-gain variant with CADD_PHRED scores ≥ 20 in 3,009 Hispanic AD cases and controls with African ancestry (see **Supplementary Table 8**). For three of these variants pairwise LD information is available from 1000 Genomes data that demonstrate strong LD (D’=1) with the respective GWAS top variants. Notably, this includes a stop-gain variant (chr9:12709125) with a CADD_Phred score of 37.0 that is 500kb upstream of our novel genome-wide significant locus within *MPDZ* on chromosome 9, a missense variant ∼900kb downstream of the chromosome 6 locus (chr6:151738529, CADD-PHRED = 24.4), and a missense variant (chr17:62049954, CADD-PHRED = 23.0) that is ∼700kb downstream of the chromosome 17 GWAS locus (see **Supplementary Table 8**).

## 4. DISCUSSION

The largest GWAS on AD in individuals of African ancestry conducted to date and the first to include individuals from continental Africa nominated one novel locus at *P* = 3.68×10^−9^ and eleven novel loci at *P* ≤ 9×10^−7^. At least one of these loci appears influenced by regional origin of African ancestry. Gene-based analyses nominated two additional loci with associations of *P*IJ≤IJ5IJ×IJ10^−5^. Of over 80 known loci implicated in NHW[1, 3], only eight were associated at nominal significance level or stronger besides *APOE*.

Most of the novel loci identified in this study cluster in known AD pathways. The novel top locus identified in our analyses, associated with a rare variant with strong regional support, is located within the *MPDZ* gene on chromosome 9p23. *MPDZ* (also known as *MUPP1*) is highly expressed in brain and encodes a modular scaffold protein (*MPDZ*)[36, 37] that is localized near the junctions of neuronal synapses[38]. It is involved in regulation of synaptic transmission by connecting neurotransmitter receptors to cytoskeletal proteins[39] and is a vital component of the NMDAR signaling complex in excitatory synapses of hippocampal neurons critical for learning and memory[38]. According to GTEx data from brain tissue[40], *MPDZ* contains six splice site variants, three of which are in a haplotype block with the top variant identified in our analyses (see **Supplementary Figure 10**). As described above, the top variant is also in LD with a functional stop-gain variant identified in ADSP sequence data.

The loci at chromosomes 2p25 and 3p25 are common and robustly present in at least 15 of the 17 contributing datasets. The top variant at 2p25 is located within a long intergenic non-coding RNA (lincRNA). lincRNAs play an essential role in RNA transcription, translation, and regulation of gene expression, as well as chromatin remodeling and genomic imprinting, significantly impacting regulation of cell proliferation, survival, and differentiation[41]. Human and animal studies link lincRNAs to the onset and pathogenesis of various neurodegenerative diseases, including AD[42, 43]. Notably, LINC01250 identified in our study also emerged as a top hit in a recent genome-wide association study of brain amyloid deposition[44]. The locus on chromosome 3p25 is located within the *SRGAP3* gene involved in nervous system development[45, 46] and has been implicated in cognitive functioning[47] and intellectual disability[48, 49]. *SRGAP3* is highly expressed in brain and is downregulated in the parahippocampal gyrus and temporal cortex in AD cases compared to cognitively healthy individuals[50]. In line with this finding, in our study *SRGAP3* expression was downregulated in zebrafish brain after inducing amyloid toxicity.

The additional identified loci, all associated with rare variants, also cluster near/in genes involved in biologically plausible pathways. *KIDINS220* encodes a multi-functional scaffold protein that is expressed in the nervous system and plays a role in neuronal survival, neurite outgrowth, differentiation into axons and dendrites, synaptic transmission, and synaptic plasticity[51, 52]. In rodent models, knockdown of Kidins220 leads to severe learning and memory deficits[53]. *TEX41* encodes a long non-coding RNA that, as described above, can lead to disease through their roles in RNA transcription, translation, regulation of gene expression, and chromatin remodeling[41]. *UNC5C* codes for a netrin receptor protein that directs axon extension and cell migration during neural development[54–56]. Notably, variants in *UNC5C* have been identified in multiplex families with AD from the NIA-FBS study, an association that has been replicated in independent case-control studies[57], and the gene encoding UNC5C-like (UNC5CL) protein is an implicated risk locus in NHW[1] that has also been shown in a methylation study of Alzheimer’s disease to be associated with key AD neuropathologic changes[58]. The present study is the first to report association at this locus in individuals of African ancestry. *PLEKHG1* is a protein coding gene involved in Rho GTPase signaling that has been associated with cognitive decline[59], white matter hyperintensities[60], and also high blood pressure in African Americans[61]. White matter hyperintensities and high blood pressure have a high prevalence in individuals of African ancestries and are associated with memory impairment and AD[62]. *SDK1* encodes an immunoglobulin superfamily cell adhesion protein required for appropriate synaptic connectivity between neurons[63]. *MMP16* encodes a matrix metalloproteinase family protein involved in the breakdown of extracellular matrix. MMPs have been implicated in several neuropathological processes including inflammation, bloodIJbrain barrier damage, and neuronal cell death[64]. *ASCL1* encodes a protein that regulates neurogenesis and neuronal differentiation[65] by directly mediating the conversion of fibroblasts into neurons[66]. Comparison of association and LD patterns of identified loci between datasets with high and lower degrees of African ancestry showed differential association patterns at this locus suggesting an influence by origin of local African ancestry. *CNTNAP4* encodes a neurexin superfamily protein involved in neural development and synaptic transmission, [67, 68] and variants in this gene have been associated with AD and cognitive impairment in NHW[69]. *TANC2* encodes a scaffolding protein highly expressed in brain that inhibits mTOR signaling controlling long-term synaptic efficacy and memory storage[70]. The two loci identified in gene-based analyses encode a member of the SNX-FERM family involved in intracellular trafficking and endocytosis (*SNX31*)[71] and a long non-coding RNA (*C9orf139/LINC02908*). In brain RNAseq data from over 1,100 individuals all these genes are differentially expressed between AD cases and controls, while in brain RNA-seq analyses in zebrafish orthologs for *MPDZ, SRGAP3, SDK1, TANC2, MMP16* and *UNC5C* are downregulated and the ortholog of *ASCL1* is upregulated after inducing amyloid toxicity, further supporting the involvement of these genes in AD etiology. Imputation quality for all novel identified disease-associated variants was excellent, and there was no evidence of inflation, thus minimizing the possibility that the observed associations are spurious.

The results of our pathway analyses also support the notion that at least a subset of the principal causative molecular pathways (immunity, lipid processing, intracellular trafficking, DNA repair, and transcription) overlap with those in NHW, although largely with different disease-associated genes within these pathways. However, a novel AD pathway emerging from our analysis is sodium-independent organic anion transmembrane transporter activity. Organic anion transporters (OATs) are transporter proteins primarily expressed in the liver, brain, proximal tubule of the kidney, and placenta. While in the kidneys they control the excretion of common drugs, toxins, and endogenous metabolites into the urine, they transport in the brain a broad range of hydrophobic organic anions including neurotransmitter and amyloid beta metabolites across the blood brain barrier (BBB). There is significant evidence that BBB breakdown and dysfunction, and dysregulation of BBB transporters, is involved in etiology of AD[72].

This study has limitations. First, given the paucity of available African American samples for genomic research on AD and the need to maximize statistical power, we combined all samples into one discovery set and relied on data from NHW, and RNAseq and ADSP whole-genome sequence data for replication and functional validation. Additional validation will likely need to be derived from experimental studies. Second, while this is the largest GWAS on directly assessed AD in African ancestry individuals to date, our sample size was underpowered to detect associations with very rare single variants or rare variants exerting very small effects. Consequently, it is possible that there remain unidentified disease-associated variants.

In summary, while our findings support the notion that the principal molecular pathways implicated in AD etiology in individuals of African ancestry largely overlap with those in NHW individuals, they suggest that the specific disease-associated loci within these pathways differ. They further suggest, that – even within individuals of African ancestry – genetic association with AD differs and is influenced by local genetic ancestry. These observations have critical implications for our quest to fully disentangle the genetic influences on the biology of AD and to develop more effective, population-specific druggable targets. First, our findings provide significant support for the importance of lipid metabolism, native immune response, intracellular trafficking, nervous system development, and synaptic plasticity in AD etiology and suggest that these pathways are critical in disease etiology across ethnic groups. At the same time, they suggest that there are also pathways whose contributions to disease differ in individuals of African ancestry compared to NHW and other populations. Sodium-independent organic anion transmembrane transporter activity was identified as a novel, plausible disease mechanism in these individuals of African ancestry, while amyloid and tau pathology are not represented among the top pathways. It is important to note however that associations with amyloid and tau pathways may be revealed as larger studies of AD in African ancestry populations are conducted, as was the case with GWAS analyses of AD in non-Hispanic White populations[3]. The observation of differential association patterns within individuals modulated by regional origin of African ancestry underscores the importance of comprehensive analysis of local ancestry at disease-associated genetic loci, and the urgent need to assemble larger sample sets from Africa that cover as much of the African genetic diversity as possible[73]. It is likely that both an overall increase of the total sample size as well as higher and broader representation of individuals of high African ancestry – as is currently underway through the ADGC, ADSP-FUS and READD-ADSP efforts – will facilitate the identification of additional disease-associated loci and significantly help to disentangle local genetic ancestry effects.

## Data Availability

All data produced will be available online at https://www.niagads.org/

## Acknowledgements

This study was supported by NIH grants: U19AG074865 (MPV, CR, JLH, WB, BK, GB, AO, RA), R01AG072547 (MPV, CR, GB, JLH, RA), U01AG058654 (JLH, MPV, WSB, ERM, LAF, FR, MC, JMV), R01AG048927 (LAF), R01AG064614 (CR), U24AG056270 (RM, CR), P30AG066462 (CR), AG057659 (MPV), AG062943 (MPV)

The National Institutes of Health, National Institute on Aging (NIH-NIA) supported this work through the following grants: ADGC, U01 AG032984, RC2 AG036528; Samples from the National Cell Repository for Alzheimer’s Disease (NCRAD), which receives government support under a cooperative agreement grant (U24 AG21886) awarded by the National Institute on Aging (NIA), were used in this study. We thank contributors who collected samples used in this study, as well as patients and their families, whose help and participation made this work possible; Data for this study were prepared, archived, and distributed by the National Institute on Aging Alzheimer’s Disease Data Storage Site (NIAGADS) at the University of Pennsylvania (U24-AG041689); GCAD, U54 AG052427; NACC, U01 AG016976; NIA FBS (Columbia University), U24 AG026395, U24 AG026390, R01AG041797; Banner Sun Health Research Institute P30 AG019610; Boston University, P30 AG013846, U01 AG10483, R01 CA129769, R01 MH080295, R01 AG017173, R01 AG025259, R01 AG048927, R01AG33193, R01 AG009029; Columbia University, P50 AG008702, R37 AG015473, R01 AG037212, R01 AG028786; Duke University, P30 AG028377, AG05128; Emory University, AG025688; Group Health Research Institute, UO1 AG006781, UO1 HG004610, UO1 HG006375, U01 HG008657; Indiana University, P30 AG10133, R01 AG009956, RC2 AG036650; Johns Hopkins University, P50 AG005146, R01 AG020688; Massachusetts General Hospital, P50 AG005134; Mayo Clinic, P50 AG016574, R01 AG032990, KL2 RR024151; Mount Sinai School of Medicine, P50 AG005138, P01 AG002219; New York University, P30 AG08051, UL1 RR029893, 5R01AG012101, 5R01AG022374, 5R01AG013616, 1RC2AG036502, 1R01AG035137; North Carolina A&T University, P20 MD000546, R01 AG28786-01A1; Northwestern University, P30 AG013854; Oregon Health & Science University, P30 AG008017, R01 AG026916; Rush University, P30 AG010161, R01 AG019085, R01 AG15819, R01 AG17917, R01 AG030146, R01 AG01101, RC2 AG036650, R01 AG22018; TGen, R01 NS059873; REAADI study is supported by NIA grant AG052410; University of Alabama at Birmingham, P50 AG016582; University of Arizona, R01 AG031581; University of California, Davis, P30 AG010129; University of California, Irvine, P50 AG016573; University of California, Los Angeles, P50 AG016570; University of California, San Diego, P50 AG005131; University of California, San Francisco, P50 AG023501, P01 AG019724; University of Kentucky, P30 AG028383, AG05144; University of Michigan, P50 AG008671; University of Pennsylvania, P30 AG010124; University of Pittsburgh, P50 AG005133, AG030653, AG041718, AG07562, AG02365; University of Southern California, P50 AG005142; University of Texas Southwestern, P30 AG012300; University of Miami, R01 AG027944, AG010491, AG027944, AG021547, AG019757; University of Washington, P50 AG005136, R01 AG042437; University of Wisconsin, P50 AG033514; Vanderbilt University, R01 AG019085; and Washington University, P50 AG005681, P01 AG03991, P01 AG026276. The Kathleen Price Bryan Brain Bank at Duke University Medical Center is funded by NINDS grant # NS39764, NIMH MH60451 and by Glaxo Smith Kline. Support was also from the Alzheimer’s Association (LAF, IIRG-08-89720; MP-V, IIRG-05-14147), the US Department of Veterans Affairs Administration, Office of Research and Development, Biomedical Laboratory Research Program, and BrightFocus Foundation (MP-V, A2111048). P.S.G.-H. is supported by Wellcome Trust, Howard Hughes Medical Institute, and the Canadian Institute of Health Research. Genotyping of the TGEN2 cohort was supported by Kronos Science. The TGen series was also funded by NIA grant AG041232 to AJM and MJH, The Banner Alzheimer’s Foundation, The Johnnie B. Byrd Sr. Alzheimer’s Institute, the Medical Research Council, and the state of Arizona and also includes samples from the following sites: Newcastle Brain Tissue Resource (funding via the Medical Research Council, local NHS trusts and Newcastle University), MRC London Brain Bank for Neurodegenerative Diseases (funding via the Medical Research Council),South West Dementia Brain Bank (funding via numerous sources including the Higher Education Funding Council for England (HEFCE), Alzheimer’s Research Trust (ART), BRACE as well as North Bristol NHS Trust Research and Innovation Department and DeNDRoN), The Netherlands Brain Bank (funding via numerous sources including Stichting MS Research, Brain Net Europe, Hersenstichting Nederland Breinbrekend Werk, International Parkinson Fonds, Internationale Stiching Alzheimer Onderzoek), Institut de Neuropatologia, Servei Anatomia Patologica, Universitat de Barcelona. ADNI data collection and sharing was funded by the National Institutes of Health Grant U01 AG024904 and Department of Defense award number W81XWH-12-2-0012. ADNI is funded by the National Institute on Aging, the National Institute of Biomedical Imaging and Bioengineering, and through generous contributions from the following: AbbVie, Alzheimer’s Association; Alzheimer’s Drug Discovery Foundation; Araclon Biotech; BioClinica, Inc.; Biogen; Bristol-Myers Squibb Company; CereSpir, Inc.; Eisai Inc.; Elan Pharmaceuticals, Inc.; Eli Lilly and Company; EuroImmun; F. Hoffmann-La Roche Ltd and its affiliated company Genentech, Inc.; Fujirebio; GE Healthcare; IXICO Ltd.; Janssen Alzheimer Immunotherapy Research & Development, LLC.; Johnson & Johnson Pharmaceutical Research & Development LLC.; Lumosity; Lundbeck; Merck & Co., Inc.; Meso Scale Diagnostics, LLC.; NeuroRx Research; Neurotrack Technologies; Novartis Pharmaceuticals Corporation; Pfizer Inc.; Piramal Imaging; Servier; Takeda Pharmaceutical Company; and Transition Therapeutics. The Canadian Institutes of Health Research is providing funds to support ADNI clinical sites in Canada. Private sector contributions are facilitated by the Foundation for the National Institutes of Health (www.fnih.org). The grantee organization is the Northern California Institute for Research and Education, and the study is coordinated by the Alzheimer’s Disease Cooperative Study at the University of California, San Diego. ADNI data are disseminated by the Laboratory for Neuro Imaging at the University of Southern California. The results published here are in whole or in part based on data obtained from Agora, a platform initially developed by the NIA-funded AMP-AD consortium that shares evidence in support of AD target discovery. Agora is available at: https://agora.adknowledgeportal.org/

The RNAseq results published here are in whole or in part based on data obtained from the AD Knowledge Portal (https://adknowledgeportal.org). Study data were provided by the Rush Alzheimer’s Disease Center, Rush University Medical Center, Chicago. Data collection was supported through funding by NIA grants P30AG10161 (ROS), R01AG15819 (ROSMAP; genomics and RNAseq), R01AG17917 (MAP), R01AG30146, R01AG36042 (5hC methylation, ATACseq), RC2AG036547 (H3K9Ac), R01AG36836 (RNAseq), R01AG48015 (monocyte RNAseq) RF1AG57473 (single nucleus RNAseq), U01AG32984 (genomic and whole exome sequencing), U01AG46152 (ROSMAP AMP-AD, targeted proteomics), U01AG46161(TMT proteomics), U01AG61356 (whole genome sequencing, targeted proteomics, ROSMAP AMP-AD), the Illinois Department of Public Health (ROSMAP), and the Translational Genomics Research Institute (genomic). Additional phenotypic data can be requested at www.radc.rush.edu. The data from Mount Sinai Brain Bank (MSBB) were generated from postmortem brain tissue collected through the Mount Sinai VA Medical Center Brain Bank and were provided by Dr. Eric Schadt from Mount Sinai School of Medicine.

## DEI Statement

This study was approved by all appropriate university institutional review boards and was performed in accordance with the ethical standards described in the 1964 Declaration of Helsinki and its later amendments. This study specifically addresses the topic of diversity, equity, and inclusion by targeting individuals of African descent, who are vastly underrepresented in AD research.

