## Supplemental Materials for "Extended genome-wide association study employing the African Genome Resources Panel identifies novel susceptibility loci for Alzheimer’s Disease in individuals of African ancestry"

**SUPPLEMENTARY MATERIALS**

[References	5](#_Toc133318727)6

### Description of Cohorts

The study included subjects from the Adult Changes in Thought (ACT) Study^1^, the National Institute on Aging (NIA) Alzheimer‘s Disease Centers (ADCs)^2^, the University of Miami/Vanderbilt University (UM/VU)^3,4^, the Mount Sinai School of Medicine (MSSM) Brain Bank^5^, the Washington Heights Inwood Columbia Aging Project (WHICAP)^6^, The African American Alzheimer's Disease Genetics (AAG) Study^7^, the MIRAGE Study^8^, NIA-FBS/NCRAD^9^, the Mayo Clinic^10^, the Rush University Alzheimer’s disease Center (ROS/MAP, MARS/CORE)^11-14^, the Chicago Health and Aging Project (CHAP)^15,16^, the Indianapolis Ibadan Dementia Study (Indianapolis)^17^, the Genetic and Environmental Risk Factors for Alzheimer’s Disease Among African Americans (GenerAAtions) Study^18,19^, the University of Pittsburgh (UP)^18,20^, and Washington University (WU)^21-24^. As described in the main text, the analyses were restricted to individuals of African American ancestry. All subjects were recruited under protocols approved by the appropriate Institutional Review Boards.

**The Adult Changes in Thought Study (ACT).** The ACT cohort^1^ is an urban and suburban elderly population from a stable HMO. The original cohort of 2,581 cognitively intact participants age ≥ 65 were enrolled between 1994 and 1998; of these 4% were African American. An additional 811 participants were enrolled in 2000-2002 using the same methods except oversampling clinics with more minorities, resulting in an overall rate of 5% African Americans. More recently, a continuous enrollment strategy was initiated in which new participants are contacted, screened, and enrolled to keep 2,000 active at-risk person-years accruing in each calendar year. This resulted in an overall enrollment of 4,729 participants as of June 2012, of whom 193 (4.1%) were African American. All clinical data are reviewed at a consensus conference. Dementia onset is assigned halfway between the prior biennial and the exam that diagnosed dementia. Enrollment for the eMERGE Study began in 2007. A waiver of consent was obtained from the IRB to enroll deceased ACT participants, and consent for data sharing was obtained from living participants. In total, ACT/eMERGE contributed data on 32 individuals with probable or possible AD and on 65 cognitively normal elders (CNEs) who were included in analyses.

**The African American Alzheimer's Disease Genetics (AAG) Study.** Participants of the multisite AAG study^7^ that contributed to this study were recruited between 2008 and 2011 from communities surrounding four locations: Columbia University in New York City, NY, North Carolina State A&T University in Greensboro, NC, University of Miami, FL, and Vanderbilt University in Nashville, TN. Participants were recruited from various sources, including naturally occurring retirement communities, churches, Black fraternal and other organizations, community centers, health fairs, physician’s offices, newspaper ads, and word-of-mouth. All participants were age 60 and older and described themselves as non-Hispanic and Black. A one-time in-person evaluation included a comprehensive neuropsychological test battery, a medical and neurological examination, an assessment of memory complaints, and an informant interview assessing functional status and possible change in cognitive performance and daily activities.

These data were evaluated in a consensus conference and diagnoses were based on standard research criteria^25^ and categorized according to National Alzheimer’s Coordinating Center (NACC) criteria. Blood was drawn and sent to the National Cell Repository for Alzheimer’s Disease (NCRAD). The current study included 624 people with AD and 161 controls from the AAG cohort. DNA was prepared by NCRAD for genotyping and sent to the genotyping site at Children’s Hospital of Philadelphia.

**The NIA ADC Samples (ADC).** The NIA ADC cohort^12^ included subjects ascertained and evaluated by the clinical and neuropathology cores of the 29 NIA-funded ADCs. Data collection is coordinated by the NACC. NACC coordinates collection of phenotype data from the 29 ADCs, cleans all data, coordinates implementation of definitions of AD cases and controls, and coordinates collection of samples. The ADC cohort consists of 228 autopsy-confirmed or clinically-confirmed African American AD cases, and 189 autopsy-confirmed or clinically-confirmed CNEs who were older than 60 years at death or at assessment. Based on the data collected by NACC, the ADGC Neuropathology Core Leaders Subcommittee derived inclusion and exclusion criteria for AD and control samples. The clinical evaluation was made using the Uniform dataset (UDS) protocol.

AD cases were demented according to DSM-IV criteria or Clinical Dementia Rating (CDR) ≥ 1. Neuropathologic stratification of cases followed NIA/Reagan criteria explicitly or used a similar approach when NIA/Reagan criteria were coded as not done, missing, or unknown. Cases were intermediate or high likelihood by NIA/Reagan criteria with moderate to frequent amyloid plaques and neurofibrillary tangle (NFT) Braak stage of III-VI. Persons with Down’s syndrome, non-AD tauopathies, and synucleinopathies were excluded. All autopsied controls had a clinical evaluation within two years of death. Controls did not meet DSM-IV criteria for dementia, did not have a diagnosis of mild cognitive impairment (MCI), and had a CDR of 0, if performed. Controls also did not meet or were low-likelihood AD by NIA/Reagan criteria, had sparse or no amyloid plaques, and a Braak NFT stage of 0 – II. ADCs sent frozen tissue from autopsied subjects and DNA samples from some autopsied subjects and from living subjects to the ADCs to NCRAD. DNA was prepared by NCRAD for genotyping and sent to the genotyping site at the Children’s Hospital of Philadelphia. ADC samples were genotyped and analyzed in separate batches. The subjects included in this study were genotyped in three waves. While most neuropathologically and clinically characterized cases and CNEs were part of the first two waves (ADC1 and ADC2, n=62 cases and 77 CNEs), the third wave consisted of clinically-identified living cases and CNEs (ADC3, n=186 cases and 112 CNEs.

**The Chicago Health and Aging Project (CHAP).** A longitudinal cohort study^15,16^ of all participating residents 65-years-of-age and older of a geographically defined biracial community located on the southwest side of Chicago. At each of six data collection cycles (every three years), all subjects underwent brief cognitive testing and a stratified random sample of about 500-600 subjects (aggregate 2844) underwent detailed clinical evaluation. The subjects provided for analysis were diagnosed with prevalent or incident Alzheimer’s disease at these clinical evaluations.

**The Genetic and Environmental Risk Factors for Alzheimer’s Disease Among African Americans (GenerAAtions) Study.** Participants of the GenerAAtions Study^18,19^ were identified through the electronic claims database of the Henry Ford Health System. Community-dwelling African Americans aged 65 and older who had at least one encounter with the Henry Ford Health System in the three years prior to their recruitment and who had an available proxy informant were eligible for this study. Cases met NINCDS-ADRDA criteria for possible or probable AD, determined in a consensus conference which included a behavioral neurologist, psychiatrist, neuropsychologist, and a behavioral neurology nurse practitioner. Phenotypic and GWAS data were available for 242 AD cases and 204 cognitively normal controls. GWAS genotyping of this sample was performed using the Illumina 660 chip as previously described^18,19^.

**Indianapolis Cohort of the Indianapolis Ibadan Dementia Study, Indiana University (IU).** The African American participants that were included in this study (173 cases, 1002 controls)^17^ were part of the community-based longitudinal comparative epidemiological study of African Americans in Indianapolis, and Yoruba Nigerians living in the city of Ibadan. In 1992, enrollment staff employed home visits to randomly sampled residential addresses in 29 contiguous U.S. Census tracts. Entry criteria were: age ≥ 65, self-identified African American, and living at sampled address. At that time 2,212 participants were enrolled. In 2001 new participants were enrolled using random sampling from Medicare rolls, with entry criteria: age ≥ 70, and self-identified African American. At that time 1,892 participants were enrolled. Participants were evaluated every two to three years with the Community Screening Interview for Dementia (CSI- D). Based on CSI-D scores individuals were selected for a full diagnostic clinical assessment including: CERAD neuropsychological battery, physical and neurological exam, and informant interview. Diagnoses were made by a panel of clinicians using standard criteria^17^. As part of the ADSP Follow-up study (ADSP-FUS) Nigerian samples from this cohort were whole-genome sequenced (“Ibadan” dataset in the present analyses).

**Mayo Clinic.** Included from the Mayo Clinic were 64 cases and 195 CNEs^10^. All subjects were diagnosed by a neurologist at the Mayo Clinic in Jacksonville, Florida or Rochester, Minnesota. The neurologist confirmed a Clinical Dementia Rating score of 0 for all controls; cases had diagnoses of possible or probable AD made according to NINCDS-ADRDA criteria^25^.

**The MIRAGE Study (MIRAGE).** The MIRAGE study^8^ is a family-based genetic epidemiological study of AD that enrolled AD cases and unaffected sibling controls at 17 clinical centers in the United States, Canada, Germany, and Greece. MIRAGE contributed 51 African American cases and 65 CNEs that were genotyped on the Illumina 300k chip, and 188 African American cases and 236 CNEs that were genotyped on the Illumina 660k chip. In brief, families were ascertained through a proband meeting the NINCDS-ADRDA criteria for definite or probable AD. Unaffected sibling controls were verified as cognitively healthy based on a Modified Telephone Interview of Cognitive Status score ≥ 86.

**Mount Sinai School of Medicine (MSSM).** The MSSM dataset^11^ contains 29 African American AD cases (all neuropathologically confirmed) and 14 CNEs (all neuropathologically confirmed), recruited to the Mount Sinai Brain Bank. Subjects had been residents of the Jewish Home and Hospital in Manhattan and The Bronx, NY and were participants in a longitudinal study of aging and dementia^11^. Brains were donated by the next of kin of deceased residents. AD diagnoses were based on clinical assessment including neuropathological assessments and subjects met CERAD criteria for definite AD or probable AD. CDR assessments, based on cognitive and functional status during the last 6 months of life, had been carried for every subject.

**NIA-FBS/NCRAD.** The NIA-FBS Family Study^9^ recruited families with two or more affected siblings with LOAD and unrelated CNEs similar in age and ethnic background. A total of 35 African American familial cases and 61 unaffected individuals were recruited through the NIA-FBS study, NCRAD, and the University of Kentucky and included for analysis. One case per family was selected after determining the individual with the strictest diagnosis (definite> probable > possible LOAD). If there were multiple individuals with the strictest diagnosis, then the individual with the earliest age of onset was selected. The controls included only those samples that were neurologically evaluated to be normal and were not related to a study participant.

**The Research in African American Alzheimer Disease Initiative (REAAADI).** Study participants for the REAAADI were ascertained as part of a family-based study of Alzheimer’s Disease through the John P. Hussman Institute for Human Genomics Outreach and Diversity Core at the University of Miami, Columbia University Alzheimer Disease Research Center, Case Western Reserve University, and Maya Angelou Center for Health Equity at Wake Forest School of Medicine.. Affected individuals met National Institute of Neurological and Communicative Disorders and Stroke-Alzheimer's Disease and Related Disorders Association criteria for AD^25,26^.

**The Rush Studies (ROS/MAP/MARS/CORE).** ROS/MAP are two community-based cohort studies^11-13^. The ROS has been on-going since 1993, with a rolling admission. Through July of 2010, 1,147 older nuns, priests, and brothers from across the United States initially free of dementia who agreed to annual clinical evaluation and brain donation at the time of death completed their baseline evaluation. Of these, 89 self-reported African Americans were included in the current study. The MAP has been on-going since 1997, also with a rolling admission.

Through July of 2010, 1,392 older persons from across northeastern Illinois initially free of dementia who agreed to annual clinical evaluation and organ donation at the time of death completed their baseline evaluation and 97 self-reported African Americans were included in this meta-analysis. Details of the clinical and neuropathologic evaluations have been previously reported^11-13^. A total of 130 persons passed genotyping QC. Of these, 30 met clinical criteria for AD at the time of their last clinical evaluation or time of death and met neuropathologic criteria for AD for those on whom neuropathologic data were available, and 100 were without dementia or MCI at the time of their last clinical evaluation or time of death and did not meet neuropathologic criteria for AD for those on whom neuropathologic data were available. MARS^14^ is a community-based cohort study of older African Americans with a rolling admission. Through July of 2010, 356 self-reported African Americans without known dementia who agreed to annual clinical evaluation completed their baseline evaluation. CORE^14^ is a community-based cohort study of older African Americans with and without dementia at baseline. Through July 2010, CORE has enrolled 218 older Africans without dementia at baseline.

**University of Miami/Vanderbilt University (UM/VU).** The UM/VU dataset^3,4^ contains 110 African American cases and 189 CNEs ascertained at the University of Miami and Vanderbilt University. Each affected individual met NINCDS-ADRDA criteria for probable or definite AD with age at onset greater than 60 years as determined from specific probe questions within the clinical history provided by a reliable family informant or from documentation of significant cognitive impairment in the medical record. Cognitively healthy controls were unrelated individuals from the same catchment areas, were frequency matched by age and gender, and had a documented MMSE or 3MS score in the normal range.

**University of Pittsburgh**. The UPITT dataset^20^ contains 114 African American AD cases (of which 6 were autopsy-confirmed) recruited by the University of Pittsburgh Alzheimer’s Disease Research Center, and 79 African American CNEs ages 60 and older (2 were autopsy-confirmed). All AD cases met NINCDS/ADRDA criteria for probable or definite AD^25^.

**The Washington Heights Inwood Aging Project (WHICAP).** The African American participants that were included in the present study (170 cases, 299 controls) were part of a longitudinal cohort study enrolled by a random sampling of Medicare recipients 65 years or older residing in northern Manhattan, New York^6,27^. Each participant underwent an interview of general health and function, medical history, a neurological examination, and a neuropsychological battery. Baseline data were collected from 1999 through 2001. Follow-up data were collected at sequential intervals of 18 months. Diagnosis of dementia etiology was made based on standard criteria^25^, and severity of dementia was assessed using the Clinical Dementia Rating scale.

**Washington University (WU).** An African American LOAD case-control dataset consisting of 87 cases and 30 healthy elderly controls was used in analyses for this study^21-24^. Participants were recruited as part of a longitudinal study of healthy aging and dementia. Diagnosis of dementia etiology was made in accordance with standard criteria and methods^25^ . Severity of dementia was assessed using the Clinical Dementia Rating scale.

### Supplementary Methods

### Supplementary Table 1. Demographic characteristics of datasets

| **Dataset** | **Cases**  **N (%)** | **Controls**  **N (%)** | **Females**  **N (%)** | **Age**  **Mean (SD)** | **APOE Genotype - N (%)** | | | |
| --- | --- | --- | --- | --- | --- | --- | --- | --- |
|  |  |  |  |  | **-/-** | **-/4** | **4/4** | **missing** |
| ACT | 32 (32.99) | 65 (67.01) | 62 (63.92) | 80.53 (6.1) | 57 (58.76) | 32 (32.99) | 4 (4.12) | 4 (4.12) |
| ADC12 | 53 (42.4) | 72 (57.6) | 89 (71.2) | 73.94 (7.46) | 57 (45.6) | 55 (44) | 8 (6.4) | 5 (4) |
| ADC3 | 147 (58.57) | 104 (41.43) | 194 (77.29) | 77.4 (7.82) | 90 (35.86) | 107 (42.63) | 21 (8.37) | 33 (13.15) |
| ADC8 | 283 (40.72) | 412 (59.28) | 514 (73.96) | 73.96 (7.23) | 357 (51.37) | 287 (41.29) | 51 (7.34) | 0 |
| ADC11 | 79 (24.76) | 240 (75.24) | 245 (76.8) | 73.41 (7.47) | 172 (53.92) | 130 (40.75) | 17 (5.33) | 0 |
| CHAP | 112 (20.66) | 430 (79.34) | 359 (66.24) | 78.8 (6.68) | 325 (59.96) | 193 (35.61) | 17 (3.14) | 0 |
| Indianapolis | 172 (14.68) | 1000 (85.32) | 769 (65.61) | 83 (5.55) | 747 (63.74) | 371 (31.66) | 54 (4.61) | 0 |
| NIA-FBS/NCRAD | 34 (37.36) | 57 (62.64) | 66 (72.53) | 73.88 (6.9) | 43 (47.25) | 37 (40.66) | 11 (12.09) | 0 |
| ADGC (2013)^a^ | 811 (33.46) | 1613 (66.54) | 1801 (74.3) | 75.55 (8.54) | 1302 (53.71) | 779 (32.14) | 129 (5.32) | 214 (8.83) |
| ADGC (2018a)^b^ | 25 (32.47) | 52 (67.53) | 58 (75.32) | 79.54 (6.81) | 48 (62.34) | 24 (31.17) | 4 (5.19) | 1 (1.3) |
| Mirage 300k | 65 (57.02) | 49 (42.98) | 80 (70.18) | 70.87 (9.29) | 41 (35.96) | 61 (53.51) | 12 (10.53) | 0 |
| Mirage 600k | 188 (44.76) | 232 (55.24) | 302 (71.9) | 71.41 (9.43) | 188 (44.76) | 181 (43.1) | 49 (11.67) | 2 (0.48) |
| GenerAAtions | 240 (54.79) | 198 (45.21) | 254 (57.99) | 79.42 (6.71) | 200 (45.66) | 174 (39.73) | 32 (7.31) | 32 (7.31) |
| ADGC (2018b)^c^ | 371 (45.92) | 437 (54.08) | 615 (76.11) | 73.9 (8.85) | 426 (52.72) | 301 (37.25) | 67 (8.29) | 14 (1.73) |
| WHICAP | 162 (27.74) | 422 (72.26) | 401 (68.66) | 78.42 (7.36) | 373 (63.87) | 191 (32.71) | 19 (3.25) | 1 (0.17) |
| REAAADI | 52 (16.99) | 254 (83.01) | 251 (82.03) | 70.72 (7.6) | 172 (56.21) | 113 (36.93) | 21 (6.86) | 0 |
| Ibadan | 77 (10.92) | 628 (89.08) | 452 (64.11) | 82.1 (5.49) | 428 (60.71) | 244 (34.61) | 32 (4.54) | 1 (0.14) |

^a^ includes samples from the AAG Study, ADCs, CHAP, Mayo Clinic, MSSM, NIA-FBS/NCRAD, ROS/MAP/MARS/CORE, UM/VU, UP, WHICAP and WU

^b^ includes samples from Mayo Clinic, Kamboh, WU, WHICAP, CHAP, AAG Study

^c^ includes samples from UM/VU, North Carolina A&T, MIRAGE, ROS/MAP/MARS/CORE

### Supplementary Table 2. Genotyping Platforms used in the individual datasets.

| **Dataset** | **Platform** |
| --- | --- |
| ADC1/2 | Human660W-Quad_v1 |
| ADC3 | HumanOmniExpress-12v1 |
| ADC4 | HumanOmniExpress-12v1 |
| ADC5 | HumanOmniExpress-12v1 |
| ADC6 | HumanOmniExpress-12v1 |
| ADC7 | HumanOmniExpressExome-8 v1.2 |
| ADC8 | HumanOmniExpressExome-8 v1.2 |
| CHOP_2017 (ADC9) | Global Screening Array v1 |
| ACT | Illumina 660k |
| CHAP | Illumina 1 M |
| Indianapolis | Illumina 1 M |
| NIA-FBS/NCRAD | Illumina 610k and 370k |
| ADGC [2013]^a^ | Illumina 1Mduo (v3) |
| ADGC [2018a]^b^ | Illumina 1Mduo (v3) |
| Mirage 300k | Illumina 300k |
| Mirage 660k | Illumina 660k |
| GenerAAtions | Illumina 660k |
| ADGC [2018b]^c^ | Global Screening Array v1 |
| WHICAP | Illumina MEGA and Illumina OmniExpress-24v1 |
| REAAADI | Illumina GSAv2 chip |
| ADC11 | Global Screening Array v1 |
| Ibadan^d^ | NovaSeq platform |

^a^includes samples from AAG, ADCs, CHAP, Mayo Clinic, MSSM, NIA-FBS/NCRAD,ROS/MAP/MARS/CORE, UM/VU, UP, WHICAP and WU

^b^includes samples from Mayo Clinic, Kamboh, WU, WHICAP, CHAP, AAG Study

^c^includes samples from UM/VU, North Carolina A&T, MIRAGE, ROS/MAP/MARS/CORE

^c^whole genome sequencing

### Supplemental Table 3. Local ancestry at the chromosome 12 locus in admixed African American datasets.

|  | **rs556001137 (Model 1)** | | | | | **rs138206541 (Model 2)** | | | | |
| --- | --- | --- | --- | --- | --- | --- | --- | --- | --- | --- |
|  | AF\|AF | AF\|AI | AF\|EU | EU\|AI | EU\|EU | AF\|AF | AF\|AI | AF\|EU | EU\|AI | EU\|EU |
| **Cases** |  | | | | |  | | | | |
| GG | 1376 | 28 | 664 | 11 | 116 | 1385 | 29 | 668 | 11 | 112 |
| TG | 8 | 3 | 6 | 0 | 0 | 6 | 2 | 6 | 0 | 0 |
| **Controls** |  | | | | |  | | | | |
| GG | 2870 | 62 | 1452 | 14 | 238 | 2880 | 65 | 1451 | 15 | 236 |
| TG | 7 | 0 | 3 | 0 | 0 | 4 | 0 | 3 | 0 | 0 |

### ^a^ AF: African ancestry haplotype; AI: Amerindian ancestry haplotype; EU: European ancestry haplotype

### Supplementary Table 4. Single-marker meta-analysis results for previously reported variants in African Americans^18,28-32^. Beta coefficients are reported with respect to the minor allele.

| Variant | Gene | Chr:Pos  (GRCh37) | Minor/Major  Alleles | MAF | Model 1 | | | Model 2 | | |
| --- | --- | --- | --- | --- | --- | --- | --- | --- | --- | --- |
|  |  |  |  |  | *β* | SE | *P* | *β* | SE | *P* |
| Previously reported in Logue et al. (2011)^c^ | | | | |  |  |  |  |  |  |
| rs340849 | *PROX1-AS1* | 1:214118090 | A/C | .19 | -0.04 | 0.06 | .46 | -0.06 | 0.05 | .24 |
| rs11889338 | *AC008069.2* | 2:17427585 | A/G | .28 | 0.04 | 0.04 | .34 | 0.03 | 0.04 | .44 |
| rs17006206 | *SLC4A1AP* | 2:27907473 | G/A | .09 | -0.12 | 0.06 | .05 | -0.13 | 0.07 | .04 |
| rs2221154 | *RBMS3* | 3:28928860 | T/G | .18 | -0.05 | 0.05 | .27 | -0.05 | 0.05 | .34 |
| rs1923775 | *POLN* | 4:2103096 | T/C | .24 | 0.14 | 0.04 | .001 | 0.14 | 0.05 | .003 |
| rs10273775 | *CNTNAP2* | 7:146897403 | G/A | .41 | 0.05 | 0.04 | .17 | 0.05 | 0.04 | .19 |
| rs956225 | *RP11-785H20.1* | 8:122909687 | G/A | .04 | -0.35 | 0.10 | 8.08×10^-4^ | -0.33 | 0.11 | .003 |
| rs3750208 | *ZC3H3* | 8:144621035 | A/G | .05 | -0.05 | 0.09 | .59 | -0.01 | 0.09 | .96 |
| rs3888908 | *RP11-632K5.3* | 11:74033066 | A/G | .20 | 0.04 | 0.05 | .38 | 0.03 | 0.05 | .51 |
| rs302318 | *TMTC1* | 12:29921667 | C/T | .27 | -0.04 | 0.04 | .33 | -0.03 | 0.04 | .55 |
| rs10850408 | *RP11-116D17.2* | 12:115380393 | T/C | .34 | -0.10 | 0.04 | .007 | -0.11 | 0.04 | .006 |
| rs17511627 | *ATP8A2P3/RNF6* | 13:26724328 | C/A | .17 | 0.09 | 0.05 | .06 | 0.10 | 0.05 | .06 |
| rs17460623 | *ENOX1* | 13:44166019 | C/T | .09 | -0.16 | 0.06 | .01 | -0.18 | 0.07 | .006 |
| rs912330 | *STK24* | 13:99131294 | T/C | .13 | -0.05 | 0.05 | .33 | -0.08 | 0.06 | .19 |
| Previously reported in Logue et al. (2014) | | | | |  |  |  |  |  |  |
| rs144662445 | *AKAP9* | 7:91709085 | G/A | .01 | 0.60 | 0.24 | .01 | 0.64 | 0.25 | .01 |
| rs149979685 | *AKAP9* | 7:91732110 | T/C | .01 | 0.59 | 0.26 | .02 | 0.62 | 0.28 | .03 |
| Previously reported in Jin et al. (2015) | | | | |  |  |  |  |  |  |
| rs2234258 | *TREM2* | 6:41126429 | T/C | .04 | 0.35 | 0.10 | 3.33×10^-4^ | 0.45 | 0.10 | 1.74×10^-5^ |
| rs2234256 | *TREM2* | 6:41126655 | G/A | .13 | 0.17 | 0.05 | .002 | 0.19 | 0.06 | .001 |
| Previously reported in Jun et al. (2017)^d^ | | | | |  |  |  |  |  |  |
| rs11168036 | *PFDN1/HBEGF* | 5:139707439 | G/T | .46 | -0.05 | 0.04 | .15 | -0.06 | 0.04 | .12 |
| rs792072 | *USP6NL/ECHDC3* | 2:5593302 | G/A | .17 | -0.02 | 0.06 | .66 | 0.01 | 0.06 | .85 |
| rs263251 | *BZRAP1-AS1* | 8:131821922 | A/G | .05 | -0.20 | 0.09 | .02 | -0.25 | 0.09 | .007 |
| rs9749589 | *NFIC* | 19:3405592 | A/T | .21 | 0.04 | 0.05 | .45 | 0.03 | 0.05 | .59 |
| Previously reported in Mez et al. (2017) | | | | |  |  |  |  |  |  |
| rs112404845 | *COBL* | 7:51578022 | T/A | .01 | 0.73 | 0.21 | 5.34×10^-4^ | 0.87 | 0.22 | 6.13×10^-5^ |
| rs16961023 | *SLC10A2* | 13:103663945 | G/C | .02 | 0.15 | 0.16 | .35 | 0.11 | 0.17 | .51 |

**Supplementary Table 4 (continued)**. Single-marker meta-analysis results for all previously reported variants in African Americans^18,28-32^. Beta results are reported with respect to the minor allele.

| Variant | Gene | Chr:Pos  (GRCh37) | Minor/Major  Alleles | MAF | Model 1 | | | Model 2 | | |
| --- | --- | --- | --- | --- | --- | --- | --- | --- | --- | --- |
|  |  |  |  |  | *β* | SE | *P* | *β* | SE | *P* |
| Previously reported in Sherva et al. (2022)^e^ | | | | |  |  |  |  |  |  |
| rs116329346 | *AC016995.3* | 2:38743886 | A/G | .01 | -0.12 | 0.25 | .62 | -0.12 | 0.26 | .65 |
| rs58443395 | *AC010967.2/SCARNA16* | 2:53482134 | G/A | .11 | 0.12 | 0.06 | .04 | 0.10 | 0.06 | .09 |
| rs10197243 | *EML6* | 2:55052493 | T/C | .16 | 0.06 | 0.06 | .28 | 0.03 | 0.05 | .53 |
| rs567572378 | *KCNH8/EFHB* | 3:19633465 | - | - | - | - | - | - | - | - |
| rs11919682 | *ROBO1* | 3:78770102 | - | - | - | - | - | - | - | - |
| rs192764155 | *RP11-506N2.1* | 4:59794471 | - | - | - | - | - | - | - | - |
| rs148433063 | *RP11-340A13.2* | 4:59886649 | C/T | .02 | -0.07 | 0.15 | .64 | -0.11 | 0.14 | .41 |
| rs28377689 | *ANTXR2* | 4:80967671 | G/T | .29 | -0.03 | 0.05 | .50 | -0.03 | 0.04 | .47 |
| rs74852218 | *RNU6-374P* | 5:25701222 | A/G | .01 | 0.08 | 0.17 | .62 | 0.07 | 0.18 | 0.68 |
| rs114681435 | *EGFLAM* | 5:38361541 | G/T | .03 | 0.16 | 0.12 | .17 | 0.13 | 0.13 | .31 |
| rs2234253 | *TREM2* | 6:41129105 | - | - | - | - | - | - | - | - |
| rs73427293 | *TREM2/TREML2* | 6:41136611 | T/A | .13 | 0.17 | 0.06 | .002 | 0.19 | 0.06 | .001 |
| rs16894668 | *AL136967.1/RP11-328M4.2* | 6:41388910 | T/C | .03 | -0.05 | 0.11 | .63 | -0.09 | 0.12 | .46 |
| rs7738720 | *RP11-157D6.1/CD2AP* | 6:47395399 | T/C | .08 | -0.18 | 0.07 | .007 | -0.19 | 0.07 | .008 |
| rs4607615 | *MSRA* | 8:10279626 | G/C | .24 | 0.08 | 0.04 | .07 | 0.08 | 0.04 | .06 |
| rs73581622 | *RP11-566H8.3/RP11-566H8.1* | 8:31258426 | A/G | .01 | -0.12 | 0.23 | .62 | -0.11 | 0.25 | .66 |
| rs112395375 | *RP11-127H5.1* | 8:105698769 | G/A | .02 | -0.13 | 0.15 | .41 | -0.06 | 0.16 | .72 |
| rs76427927 | *RP11-192P9.1/TRPS1* | 8:116123287 | C/A | .06 | 0.22 | 0.08 | .005 | 0.27 | 0.08 | .001 |
| rs509334 | *BRP11-142I2.1/SORL1* | 11:121277918 | A/G | .22 | 0.02 | 0.05 | .61 | 0.02 | 0.05 | .65 |
| rs145008711 | *ITGA11* | 15:68726103 | T/A | .01 | -0.19 | 0.27 | .49 | -0.30 | 0.29 | .30 |
| rs116620371 | *FHOD3* | 18:34179992 | A/G | .01 | 0.24 | 0.20 | .23 | 0.28 | 0.22 | .20 |
| rs73505251 | *ABCA7* | 19:1068095 | A/T | .14 | 0.23 | 0.05 | 2.13×10^-5^ | 0.25 | 0.06 | 2.32×10^-5^ |

^a^MAF: minor allele frequency from Model 1

^b^“-” indicates that the variant was not present in QCed results for the specified model

^c^Results in this study were reported at P < 5×10^-5^

^d^This GWAS was conducted on a transethnic population

^e^This study used an expanded phenotype of AD and related dementia (ADRD) and AD by proxy

### Supplementary Table 5. Single-marker meta-analysis results for previously reported variants in non-Hispanic Whites (NHW)^33^. Beta coefficients are reported with respect to the minor allele.

| Variant | Gene | Chr:Pos  (GRCh38) | Chr:Pos  (GRCh37) | Minor/Major  Alleles | MAF | Model 1 | | | Model 2 | | |
| --- | --- | --- | --- | --- | --- | --- | --- | --- | --- | --- | --- |
|  |  |  |  |  |  | *β* | SE | *P* | *β* | SE | *P* |
| rs141749679 | *SORT1* | 1:109345810 | 1:109888432 | - | - | - | - | - | - | - | - |
| rs679515 | *CR1* | 1:207577223 | 1:207750568 | T/C | .037 | 0.07 | 0.13 | .61 | 0.17 | 0.11 | .13 |
| rs72777026 | *ADAM17* | 2:9558882 | 2:9699011 | G/A | .343 | 0.04 | 0.04 | .37 | 0.06 | 0.04 | .12 |
| rs17020490 | *PRKD3* | 2:37304796 | 2:37531939 | C/T | .170 | 0.02 | 0.05 | .74 | -0.01 | 0.05 | .85 |
| rs143080277 | *NCK2* | 2:105749599 | 2:106366056 | C/T | .003 | 0.48 | 0.99 | .63 | 0.84 | 0.90 | .35 |
| rs6733839 | *BIN1* | 2:127135234 | 2:127892810 | T/C | .393 | 0.11 | 0.05 | .02 | 0.14 | 0.04 | .001 |
| rs139643391 | *WDR12* | 2:202878717 | 2:203743440 | - | - | - | - | - | - | - | - |
| rs10933431 | *INPP5D* | 2:233117202 | 2:233981912 | C/G | .392 | -0.01 | 0.05 | .92 | 0.03 | 0.04 | .55 |
| rs16824536 | *MME* | 3:155069722 | 3:154787511 | A/G | .244 | 0.02 | 0.05 | .66 | 0.01 | 0.05 | .76 |
| rs61762319 | *MME* | 3:155084189 | 3:154801978 | G/A | .007 | 0.54 | 0.45 | .23 | 0.50 | 0.48 | .30 |
| rs3822030 | *IDUA* | 4:993555 | 4:987343 | T/G | .247 | 0.09 | 0.04 | .04 | 0.08 | 0.05 | .06 |
| rs6846529 | *CLNK* | 4:11023507 | 4:11025131 | C/T | .387 | 0.04 | 0.04 | .32 | 0.03 | 0.04 | .39 |
| rs2245466 | *RHOH* | 4:40197226 | 4:40198846 | G/C | .258 | 0.04 | 0.05 | .37 | 0.04 | 0.05 | .45 |
| rs112403360 | *ANKH* | 5:14724304 | 5:14724413 | A/T | .100 | 0.11 | 0.06 | .09 | 0.13 | 0.07 | .05 |
| rs62374257 | *COX7C* | 5:86927378 | 5:86223195 | C/T | .050 | 0.01 | 0.09 | .91 | 0.05 | 0.09 | .57 |
| rs871269 | *TNIP1* | 5:151052827 | 5:150432388 | T/C | .399 | 0.03 | 0.04 | .37 | 0.05 | 0.04 | .16 |
| rs113706587 | *RASGEF1C* | 5:180201150 | 5:179628150 | A/G | .037 | 0.12 | 0.11 | .27 | 0.13 | 0.12 | .26 |
| rs6605556 | *HLA-DQA1* | 6:32615322 | 6:32583099 | - | - | - | - | - | - | - | - |
| rs10947943 | *UNC5CL* | 6:41036354 | 6:41004093 | A/G | .032 | -0.22 | 0.12 | .06 | -0.29 | 0.12 | .02 |
| rs143332484 | *TREM2* | 6:41161469 | 6:41129207 | T/C | .003 | -0.47 | 0.72 | .51 | -0.53 | 0.76 | .48 |
| rs75932628 | *TREM2* | 6:41161514 | 6:41129252 | T/C | .001 | -0.63 | 1.17 | .59 | -0.02 | 1.45 | .99 |
| rs60755019 | *TREML2* | 6:41181270 | 6:41149008 | G/A | .186 | 0.09 | 0.05 | .05 | 0.09 | 0.05 | .06 |
| rs7767350 | *CD2AP* | 6:47517390 | 6:47485126 | T/C | .191 | 0.07 | 0.05 | .15 | 0.08 | 0.05 | .08 |
| rs785129 | *HS3ST5* | 6:114291731 | 6:114612895 | T/C | .226 | -0.07 | 0.05 | .17 | -0.04 | 0.05 | .34 |
| rs6943429 | *UMAD1* | 7:7817263 | 7:7856894 | T/C | .485 | 0.08 | 0.04 | .02 | 0.08 | 0.04 | .04 |
| rs10952097 | *ICA1* | 7:8204382 | 7:8244012 | C/T | .474 | 0.01 | 0.04 | .72 | 0.03 | 0.04 | .47 |
| rs13237518 | *TMEM106B* | 7:12229967 | 7:12269593 | C/A | .297 | 0.07 | 0.04 | .09 | 0.05 | 0.04 | .20 |

**Supplementary Table 5 (continued)**. Single-marker meta-analysis results for previously reported variants in NHW^33^. Beta coefficients are reported with respect to the minor allele.

| Variant | Gene | Chr:Pos  (GRCh38) | Chr:Pos  (GRCh37) | Minor/Major  Alleles | MAF | Model 1 | | | Model 2 | | |
| --- | --- | --- | --- | --- | --- | --- | --- | --- | --- | --- | --- |
|  |  |  |  |  |  | *β* | SE | *P* | *β* | SE | *P* |
| rs1160871 | *JAZF1* | 7:28129131 | 7:28168750 | - | - | - | - | - | - | - | - |
| rs6966331 | *EPDR1* | 7:37844191 | 7:37883793 | C/T | .313 | 0.00 | 0.04 | .93 | 0.01 | 0.04 | .90 |
| rs76928645 | *SEC61G* | 7:54873635 | 7:54941328 | T/C | .019 | 0.05 | 0.14 | .74 | 0.01 | 0.15 | .96 |
| rs7384878 | *SPDYE3* | 7:100334426 | 7:99932049 | C/T | .143 | -0.03 | 0.05 | .53 | -0.02 | 0.06 | .67 |
| rs11771145 | *EPHA1* | 7:143413669 | 7:143110762 | G/A | .450 | 0.06 | 0.04 | .12 | 0.06 | 0.04 | .09 |
| rs1065712 | *CTSB* | 8:11844613 | 8:11702122 | C/G | .010 | 0.20 | 0.21 | .35 | 0.08 | 0.22 | .72 |
| rs73223431 | *PTK2B* | 8:27362470 | 8:27219987 | T/C | .257 | 0.05 | 0.04 | .21 | 0.05 | 0.04 | .25 |
| rs11787077 | *CLU* | 8:27607795 | 8:27465312 | C/T | .445 | 0.02 | 0.04 | .67 | 0.02 | 0.04 | .69 |
| rs34173062 | *SHARPIN* | 8:144103704 | 8:145158607 | - | - | - | - | - | - | - | - |
| rs1800978 | *ABCA1* | 9:104903697 | 9:107665978 | G/C | .033 | -0.16 | 0.11 | .14 | -0.19 | 0.11 | .09 |
| rs7912495 | *USP6NL* | 10:11676714 | 10:11718713 | G/A | .321 | 0.08 | 0.04 | .04 | 0.08 | 0.04 | .05 |
| rs7068231 | *ANK3* | 10:60025170 | 10:61784928 | T/G | .250 | 0.00 | 0.04 | .99 | 0.00 | 0.04 | .93 |
| rs6586028 | *TSPAN14* | 10:80494228 | 10:82253984 | C/T | .046 | -0.12 | 0.09 | .19 | -0.08 | 0.09 | .40 |
| rs6584063 | *BLNK* | 10:96266650 | 10:98026407 | G/A | .055 | 0.01 | 0.08 | .94 | 0.00 | 0.09 | .99 |
| rs7908662 | *PLEKHA1* | 10:122413396 | 10:124172912 | G/A | .381 | 0.00 | 0.04 | .98 | 0.01 | 0.04 | .72 |
| rs10437655 | *SPI1* | 11:47370397 | 11:47391948 | A/G | .319 | 0.06 | 0.04 | .10 | 0.06 | 0.04 | .14 |
| rs1582763 | *MS4A4A* | 11:60254475 | 11:60021948 | A/G | .103 | 0.03 | 0.06 | .61 | 0.05 | 0.07 | .43 |
| rs3851179 | *EED* | 11:86157598 | 11:85868640 | T/C | .155 | -0.06 | 0.05 | .21 | -0.04 | 0.05 | .44 |
| rs74685827 | *SORL1* | 11:121482368 | 11:121353077 | G/T | .005 | -0.39 | 0.72 | .59 | -0.31 | 0.75 | .68 |
| rs11218343 | *SORL1* | 11:121564878 | 11:121435587 | C/T | .083 | -0.07 | 0.07 | .28 | -0.07 | 0.07 | .33 |
| rs6489896 | *TPCN1* | 12:113281983 | 12:113719788 | C/T | .165 | 0.03 | 0.05 | .49 | 0.03 | 0.05 | .61 |
| rs17125924 | *FERMT2* | 14:52924962 | 14:53391680 | G/A | .069 | -0.14 | 0.07 | .05 | -0.21 | 0.08 | .01 |
| rs7401792 | *SLC24A4* | 14:92464917 | 14:92931261 | A/G | .253 | 0.03 | 0.04 | .41 | 0.05 | 0.05 | .29 |
| rs12590654 | *SLC24A4* | 14:92472511 | 14:92938855 | A/G | .359 | 0.02 | 0.04 | .66 | 0.02 | 0.04 | .65 |
| rs7157106 | *IGH* | 14:105761758 | 14:106228095 | G/A | .173 | -0.01 | 0.06 | .83 | -0.01 | 0.06 | .88 |
| rs10131280 | *IGH* | 14:106665591 | 14:107121607 | A/G | .208 | 0.03 | 0.05 | .53 | 0.03 | 0.05 | .59 |
| rs8025980 | *SPPL2A* | 15:50701814 | 15:50994011 | G/A | .416 | 0.04 | 0.04 | .29 | 0.02 | 0.04 | .59 |
| rs602602 | *MINDY2* | 15:58764824 | 15:59057023 | A/T | .243 | 0.07 | 0.04 | .08 | 0.07 | 0.05 | .10 |

**Supplementary Table 5 (continued)**. Single-marker meta-analysis results for previously reported variants in NHW^33^. Beta coefficients are reported with respect to the minor allele.

| Variant | Gene | Chr:Pos  (GRCh38) | Chr:Pos  (GRCh37) | Minor/Major  Alleles | MAF | Model 1 | | | Model 2 | | |
| --- | --- | --- | --- | --- | --- | --- | --- | --- | --- | --- | --- |
|  |  |  |  |  |  | *β* | SE | *P* | *β* | SE | *P* |
| rs117618017 | *APH1B* | 15:63277703 | 15:63569902 | T/C | .027 | 0.07 | 0.14 | .61 | 0.02 | 0.15 | .92 |
| rs3848143 | *SNX1* | 15:64131307 | 15:64423506 | G/A | .391 | -0.03 | 0.04 | .47 | -0.04 | 0.04 | .29 |
| rs12592898 | *CTSH* | 15:78936857 | 15:79229199 | A/G | .228 | -0.03 | 0.04 | .43 | -0.03 | 0.05 | .51 |
| rs1140239 | *DOC2A* | 16:30010081 | 16:30021402 | T/C | .182 | -0.02 | 0.05 | .60 | -0.02 | 0.05 | .75 |
| rs889555 | *BCKDK* | 16:31111250 | 16:31122571 | T/C | .404 | 0.01 | 0.04 | .70 | 0.02 | 0.04 | .59 |
| rs4985556 | *IL34* | 16:70660097 | 16:70694000 | A/C | .027 | -0.15 | 0.12 | .21 | -0.24 | 0.13 | .06 |
| rs450674 | *MAF* | 16:79574511 | 16:79608408 | C/T | .254 | 0.06 | 0.04 | .17 | 0.07 | 0.04 | .10 |
| rs12446759 | *PLCG2* | 16:81739398 | 16:81773003 | A/G | .264 | -0.06 | 0.04 | .19 | -0.06 | 0.05 | .21 |
| rs72824905 | *PLCG2* | 16:81908423 | 16:81942028 | G/C | .002 | 0.18 | 0.70 | .79 | -0.06 | 0.75 | .94 |
| rs16941239 | *FOXF1* | 16:86420604 | 16:86454210 | A/T | .183 | -0.03 | 0.05 | .48 | -0.04 | 0.05 | .47 |
| rs56407236 | *PRDM7* | 16:90103687 | 16:90170095 | A/G | .079 | 0.04 | 0.07 | .56 | 0.05 | 0.07 | .48 |
| rs35048651 | *WDR81* | 17:1728056 | 17:1631350 | - | - | - | - | - | - | - | - |
| rs7225151 | *SCIMP* | 17:5233752 | 17:5137047 | A/G | .224 | 0.08 | 0.04 | .04 | 0.08 | 0.04 | .06 |
| rs2242595 | *MYO15A* | 17:18156140 | 17:18059454 | A/G | .146 | 0.05 | 0.05 | .32 | 0.05 | 0.05 | .32 |
| rs5848 | *GRN* | 17:44352876 | 17:42430244 | C/T | .349 | -0.05 | 0.04 | .16 | -0.07 | 0.04 | .08 |
| rs199515 | *WNT3* | 17:46779275 | 17:44856641 | G/C | .120 | -0.06 | 0.06 | .27 | -0.06 | 0.06 | .31 |
| rs616338 | *ABI3* | 17:49219935 | 17:47297297 | - | - | - | - | - | - | - | - |
| rs2526377 | *TSPOAP1* | 17:58332680 | 17:56410041 | A/G | .431 | 0.06 | 0.04 | .13 | 0.05 | 0.04 | .22 |
| rs4277405 | *ACE* | 17:63471557 | 17:61548918 | C/T | .362 | -0.04 | 0.04 | .25 | -0.05 | 0.04 | .19 |
| rs12151021 | *ABCA7* | 19:1050875 | 19:1050874 | A/G | .399 | 0.06 | 0.04 | .13 | 0.06 | 0.04 | .16 |
| rs149080927 | *KLF16* | 19:1854254 | 19:1854254 | - | - | - | - | - | - | - | - |
| rs9304690 | *SIGLEC11* | 19:49950060 | 19:50453317 | T/C | .102 | 0.04 | 0.06 | .56 | 0.08 | 0.06 | .21 |
| rs587709 | *LILRB2* | 19:54267597 | 19:54771451 | - | - | - | - | - | 0.02 | 0.04 | .67 |
| rs1358782 | *RBCK1* | 20:413334 | 20:393978 | A/G | .167 | -0.08 | 0.05 | .13 | -0.04 | 0.05 | .42 |
| rs6014724 | *CASS4* | 20:56423488 | 20:54998544 | G/A | .103 | 0.02 | 0.06 | .72 | 0.03 | 0.06 | .65 |
| rs6742 | *SLC2A4RG* | 20:63743088 | 20:62374441 | T/C | .307 | 0.03 | 0.04 | .50 | 0.03 | 0.05 | .50 |
| rs2154481 | *APP* | 21:26101558 | 21:27473875 | T/C | .118 | 0.09 | 0.06 | .13 | 0.08 | 0.06 | .18 |
| rs2830489 | *ADAMTS1* | 21:26775872 | 21:28148191 | T/C | .065 | -0.04 | 0.08 | .66 | -0.06 | 0.09 | .52 |

^a^MAF: minor allele frequency from Model 1

^b^“-” indicates that the variant was not present in QCed results for the specified model

Supplementary Table 6. Gene-based results for AD genes previously identified in non-Hispanic Whites^33^.

| Gene | ID | Chr | Start BP  (hg37) | Stop BP  (hg37) | Model 1 | | | | Model 2 | | | |
| --- | --- | --- | --- | --- | --- | --- | --- | --- | --- | --- | --- | --- |
|  |  |  |  |  | No. of SNPs | No. of Parameters | *Z* | *P* | No. of SNPs | No. of Parameters | *Z* | *P* |
| *SORT1* | ENSG00000134243 | 1 | 109842192 | 109975573 | 557 | 57 | -0.42 | .66 | 857 | 80 | -0.55 | .71 |
| *CR1* | ENSG00000203710 | 1 | 207634492 | 207823992 | 953 | 118 | -1.13 | .87 | 1093 | 125 | -0.34 | .63 |
| *ADAM17* | ENSG00000151694 | 2 | 9618615 | 9730921 | 808 | 93 | -1.16 | .88 | 809 | 97 | -0.05 | .52 |
| *PRKD3* | ENSG00000115825 | 2 | 37467645 | 37586951 | 1086 | 89 | -0.75 | .77 | 1084 | 91 | -0.70 | .76 |
| *NCK2* | ENSG00000071051 | 2 | 106326354 | 106520730 | 1328 | 108 | -0.69 | .75 | 1530 | 152 | -0.92 | .82 |
| *BIN1* | ENSG00000136717 | 2 | 127795603 | 127899931 | 816 | 99 | 0.87 | .19 | 940 | 125 | 1.10 | .14 |
| *WDR12* | ENSG00000138442 | 2 | 203729505 | 203914521 | 818 | 73 | -0.71 | .76 | 1005 | 84 | 0.26 | .40 |
| *INPP5D* | ENSG00000168918 | 2 | 233889677 | 234126549 | 1517 | 208 | 0.83 | .20 | 1718 | 238 | -1.20 | .88 |
| *MME* | ENSG00000196549 | 3 | 154706913 | 154911497 | 1136 | 169 | -1.02 | .85 | 1395 | 189 | -0.84 | .80 |
| *IDUA* | ENSG00000127415 | 4 | 945785 | 1008316 | 563 | 134 | -0.27 | .61 | 562 | 139 | 0.18 | .43 |
| *CLNK* | ENSG00000109684 | 4 | 10478019 | 10721489 | 2368 | 195 | -0.03 | .51 | 2368 | 207 | -0.42 | .66 |
| *RHOH* | ENSG00000168421 | 4 | 40157673 | 40258587 | 797 | 173 | -1.26 | .90 | 797 | 162 | -0.42 | .66 |
| *ANKH* | ENSG00000154122 | 5 | 14694910 | 14906887 | 1626 | 207 | 0.35 | .36 | 1623 | 211 | 0.35 | .36 |
| *COX7C* | ENSG00000127184 | 5 | 85878721 | 85926779 | 370 | 51 | -0.74 | .77 | 383 | 56 | 0.23 | .41 |
| *TNIP1* | ENSG00000145901 | 5 | 150399506 | 150508138 | 1049 | 167 | 0.10 | .46 | 1047 | 170 | 1.15 | .13 |
| *RASGEF1C* | ENSG00000146090 | 5 | 179517795 | 179671153 | 1348 | 220 | 1.18 | .12 | 1347 | 225 | 1.53 | .06 |
| *HLA-DQA1* | ENSG00000196735 | 6 | 32560956 | 32624839 | 1442 | 49 | -0.27 | .61 | 1441 | 51 | 0.07 | .47 |
| *UNC5CL* | ENSG00000124602 | 6 | 40984772 | 41041928 | 417 | 62 | 2.21 | .01 | 418 | 57 | 2.34 | .01 |
| *TREM2* | ENSG00000095970 | 6 | 41116244 | 41165924 | 380 | 62 | 3.55 | 1.93×10^-4^ | 379 | 72 | 4.14 | 1.76×10^-5^ |
| *TREML2* | ENSG00000112195 | 6 | 41148015 | 41203932 | 607 | 97 | 2.68 | .004 | 607 | 100 | 2.98 | .001 |
| *CD2AP* | ENSG00000198087 | 6 | 47410525 | 47604999 | 1419 | 86 | 0.96 | .17 | 1423 | 88 | 1.40 | .08 |
| *HS3ST5* | ENSG00000249853 | 6 | 114366750 | 114699209 | 1800 | 194 | -1.66 | .95 | 2201 | 236 | -0.28 | .61 |
| *UMAD1* | ENSG00000219545 | 7 | 7645342 | 8053689 | 4231 | 422 | 0.13 | .45 | 4217 | 419 | -0.21 | .58 |
| *ICA1* | ENSG00000003147 | 7 | 8142814 | 8337317 | 1823 | 293 | -1.02 | .85 | 1820 | 291 | -0.59 | .72 |
| *TMEM106B* | ENSG00000106460 | 7 | 12215867 | 12292993 | 988 | 101 | 1.25 | .11 | 985 | 86 | 0.62 | .27 |
| *JAZF1* | ENSG00000153814 | 7 | 27860192 | 28255362 | 3189 | 276 | -1.57 | .94 | 3175 | 296 | -0.57 | .71 |
| *EPDR1* | ENSG00000086289 | 7 | 37688446 | 38001543 | 2723 | 191 | -0.29 | .62 | 2711 | 192 | 0.65 | .26 |
| *SEC61G* | ENSG00000132432 | 7 | 54809943 | 54862667 | 510 | 62 | -1.13 | .87 | 509 | 63 | -1.14 | .87 |
| *SPDYE3* | ENSG00000214300 | 7 | 99870325 | 99929819 | 160 | 59 | 0.75 | .23 | 160 | 59 | 0.30 | .38 |
| *EPHA1* | ENSG00000146904 | 7 | 143077382 | 143140985 | 488 | 111 | -0.20 | .58 | 489 | 113 | -0.24 | .60 |
| *CTSB* | ENSG00000164733 | 8 | 11690033 | 11761957 | 1056 | 154 | -0.55 | .71 | 1054 | 136 | -0.13 | .55 |

**Supplementary Table 6 continued.** Gene-based results for AD genes previously identified in non-Hispanic Whites^33^

| Gene | ID | Chr | Start BP  (hg37) | Stop BP  (hg37) | Model 1 | | | | Model 2 | | | |
| --- | --- | --- | --- | --- | --- | --- | --- | --- | --- | --- | --- | --- |
|  |  |  |  |  | No. of SNPs | No. of Parameters | *Z* | *P* | No. of SNPs | No. of Parameters | *Z* | *P* |
| *PTK2B* | ENSG00000120899 | 8 | 27133999 | 27326903 | 1765 | 143 | -0.90 | .82 | 1757 | 142 | -0.20 | .58 |
| *CLU* | ENSG00000120885 | 8 | 27444434 | 27507548 | 531 | 122 | 0.25 | .40 | 529 | 121 | -0.28 | .61 |
| *SHARPIN* | ENSG00000179526 | 8 | 145143536 | 145198027 | 364 | 59 | -0.51 | .70 | 364 | 57 | -0.14 | .56 |
| *ABCA1* | ENSG00000165029 | 9 | 107533283 | 107725518 | 1744 | 267 | 0.39 | .35 | 1735 | 263 | 0.46 | .32 |
| *USP6NL* | ENSG00000148429 | 10 | 11485945 | 11688753 | 1450 | 101 | -0.95 | .83 | 1453 | 104 | -0.41 | .66 |
| *ANK3* | ENSG00000151150 | 10 | 61776056 | 62528248 | 6782 | 360 | -1.01 | .84 | 6779 | 356 | -0.73 | .77 |
| *TSPAN14* | ENSG00000108219 | 10 | 82178922 | 82302879 | 1118 | 134 | 0.39 | .35 | 1116 | 138 | -0.34 | .63 |
| *BLNK* | ENSG00000095585 | 10 | 97941458 | 98066344 | 932 | 158 | -0.21 | .58 | 933 | 151 | -0.76 | .78 |
| *PLEKHA1* | ENSG00000107679 | 10 | 124099212 | 124201867 | 815 | 110 | 0.16 | .44 | 812 | 109 | -0.11 | .54 |
| *SPI1* | ENSG00000066336 | 11 | 47366411 | 47435127 | 505 | 88 | 1.68 | .05 | 502 | 75 | 0.88 | .19 |
| *MS4A4A* | ENSG00000110079 | 11 | 60013014 | 60086445 | 595 | 94 | -0.21 | .58 | 593 | 95 | -0.57 | .72 |
| *EED* | ENSG00000074266 | 11 | 85920586 | 85999855 | 650 | 62 | 0.00 | .50 | 648 | 67 | -0.25 | .60 |
| *SORL1* | ENSG00000137642 | 11 | 121287912 | 121514402 | 1525 | 223 | 0.67 | .25 | 1520 | 219 | 0.10 | .46 |
| *TPCN1* | ENSG00000186815 | 12 | 113623855 | 113746390 | 866 | 75 | -1.03 | .85 | 861 | 75 | -1.39 | .92 |
| *FERMT2* | ENSG00000073712 | 14 | 53313986 | 53454153 | 1137 | 127 | -0.77 | .78 | 1129 | 102 | -0.04 | .51 |
| *SLC24A4* | ENSG00000140090 | 14 | 92753925 | 92972596 | 2142 | 243 | 0.77 | .22 | 2134 | 248 | 1.11 | .13 |
| *SPPL2A* | ENSG00000138600 | 15 | 50989506 | 51093005 | 868 | 103 | -0.42 | .66 | 866 | 95 | -0.82 | .79 |
| *MINDY2* | ENSG00000128923 | 15 | 59028391 | 59164099 | 1049 | 115 | 0.38 | .35 | 1048 | 108 | 0.51 | .31 |
| *APH1B* | ENSG00000138613 | 15 | 63533217 | 63611325 | 545 | 123 | 0.50 | .31 | 545 | 118 | 0.83 | .20 |
| *SNX1* | ENSG00000028528 | 15 | 64351322 | 64448289 | 724 | 74 | 0.49 | .31 | 721 | 71 | 0.52 | .30 |
| *CTSH* | ENSG00000103811 | 15 | 79203400 | 79276916 | 653 | 108 | -1.13 | .87 | 651 | 110 | -1.86 | .97 |
| *DOC2A* | ENSG00000149927 | 16 | 30006830 | 30069591 | 339 | 56 | 0.11 | .46 | 340 | 60 | -0.42 | .66 |
| *BCKDK* | ENSG00000103507 | 16 | 31082428 | 31134110 | 252 | 76 | -0.84 | .80 | 251 | 76 | -0.47 | .68 |
| *IL34* | ENSG00000157368 | 16 | 70578798 | 70704585 | 1084 | 84 | 0.65 | .26 | 1084 | 102 | 0.86 | .19 |
| *MAF* | ENSG00000178573 | 16 | 79609740 | 79669611 | 543 | 150 | -0.18 | .57 | 541 | 149 | 0.23 | .41 |
| *PLCG2* | ENSG00000197943 | 16 | 81737702 | 82001899 | 3615 | 383 | -0.66 | .75 | 3602 | 392 | -0.38 | .65 |
| *FOXF1* | ENSG00000103241 | 16 | 86509133 | 86558076 | 657 | 139 | 0.34 | .37 | 656 | 129 | 1.02 | .15 |
| *PRDM7* | ENSG00000126856 | 16 | 90112974 | 90193480 | 514 | 102 | 0.17 | .43 | 514 | 102 | -0.75 | .77 |
| *WDR81* | ENSG00000167716 | 17 | 1584817 | 1651893 | 431 | 100 | 1.04 | .15 | 431 | 94 | 0.66 | .26 |
| *SCIMP* | ENSG00000161929 | 17 | 5102256 | 5173155 | 639 | 113 | 0.28 | .39 | 639 | 113 | 0.48 | .32 |
| *MYO15A* | ENSG00000091536 | 17 | 17977020 | 18093116 | 786 | 133 | 0.16 | .43 | 785 | 126 | 0.05 | .48 |

**Supplementary Table 6 continued.** Gene-based results for AD genes previously identified in non-Hispanic Whites^33^

| Gene | ID | Chr | Start BP  (hg37) | Stop BP  (hg37) | Model 1 | | | | Model 2 | | | |
| --- | --- | --- | --- | --- | --- | --- | --- | --- | --- | --- | --- | --- |
|  |  |  |  |  | No. of SNPs | No. of Parameters | *Z* | *P* | No. of SNPs | No. of Parameters | *Z* | *P* |
| *GRN* | ENSG00000030582 | 17 | 42387614 | 42440470 | 312 | 99 | 0.24 | .41 | 309 | 98 | 0.30 | .38 |
| *WNT3* | ENSG00000108379 | 17 | 44829872 | 44945520 | 722 | 139 | 0.87 | .19 | 720 | 158 | 1.11 | .13 |
| *ABI3* | ENSG00000108798 | 17 | 47252589 | 47310587 | 420 | 75 | -1.63 | .95 | 419 | 78 | -1.76 | .96 |
| *TSPOAP1* | ENSG00000005379 | 17 | 56368592 | 56441152 | 520 | 118 | 0.35 | .36 | 516 | 116 | 0.10 | .46 |
| *ACE* | ENSG00000264813 | 17 | 61527184 | 61609209 | 648 | 152 | -0.43 | .67 | 651 | 153 | -0.07 | .53 |
| *ABCA7* | ENSG00000064687 | 19 | 1005102 | 1075571 | 765 | 116 | 3.14 | 8.46×10^-4^ | 765 | 116 | 3.78 | 7.76×10^-5^ |
| *KLF16* | ENSG00000129911 | 19 | 1842399 | 1898567 | 402 | 70 | 1.35 | .09 | 400 | 73 | 2.03 | .02 |
| *SIGLEC11* | ENSG00000161640 | 19 | 50442242 | 50499429 | 572 | 68 | -0.50 | .69 | 574 | 69 | -0.36 | .64 |
| *LILRB2* | ENSG00000131042 | 19 | 54767675 | 54820039 | 401 | 62 | -0.17 | .57 | 401 | 62 | 0.54 | .30 |
| *RBCK1* | ENSG00000125826 | 20 | 353142 | 421610 | 627 | 143 | 1.48 | .07 | 623 | 149 | 1.46 | .07 |
| *CASS4* | ENSG00000087589 | 20 | 54952168 | 55044396 | 777 | 170 | 0.13 | .45 | 777 | 176 | 0.06 | .48 |
| *SLC2A4RG* | ENSG00000125520 | 20 | 62336214 | 62384858 | 322 | 63 | -0.88 | .81 | 320 | 63 | -0.48 | .69 |
| *APP* | ENSG00000142192 | 21 | 27242861 | 27578446 | 2792 | 270 | 0.73 | .23 | 2780 | 269 | 0.17 | .43 |
| *ADAMTS1* | ENSG00000154734 | 21 | 28198066 | 28252728 | 475 | 94 | 0.34 | .37 | 475 | 94 | 0.69 | .25 |

^a^Model 1 is adjusted for PCs, age, sex

^b^Model 2 is adjusted for PCs, age, sex, APOE genotype

### Supplementary Table 7. Gene-based results for AD genes previously identified in African Americans but not NHW^18,28-32,34-36^

| Gene | ID | Chr | Start BP  (hg37) | Stop BP  (hg37) | Model 1 | | | | Model 2 | | | |
| --- | --- | --- | --- | --- | --- | --- | --- | --- | --- | --- | --- | --- |
|  |  |  |  |  | No. of SNPs | No. of Parameters | *Z* | *P* | No. of SNPs | No. of Parameters | *Z* | *P* |
| *SIPA1L2* | ENSG00000116991 | 1 | 232523711 | 232732304 | 1565 | 184 | -1.45 | .93 | 1825 | 205 | -0.59 | .72 |
| *TSSC1* | ENSG00000032389 | 2 | 3182696 | 3416653 | 2032 | 152 | -0.56 | .71 | 2027 | 164 | 0.09 | .46 |
| *KIDINS220* | ENSG00000134313 | 2 | 8855408 | 9012760 | 778 | 109 | 0.49 | .31 | 773 | 111 | 0.33 | .37 |
| *SLC4A1AP* | ENSG00000163798 | 2 | 27851338 | 27927840 | 486 | 76 | -0.49 | .69 | 485 | 68 | 0.10 | .46 |
| *EDEM1* | ENSG00000134109 | 3 | 5194331 | 5271642 | 687 | 64 | -0.08 | .53 | 683 | 61 | 0.02 | .49 |
| *SRGAP3* | ENSG00000196220 | 3 | 9012275 | 9439737 | 3312 | 418 | 0.86 | .19 | 3300 | 422 | 0.38 | .35 |
| *RBMS3* | ENSG00000144642 | 3 | 29287473 | 30061886 | 7285 | 455 | -0.49 | .69 | 7280 | 467 | 0.05 | .48 |
| *TRANK1* | ENSG00000168016 | 3 | 36858311 | 37021548 | 1074 | 137 | 2.36 | .009 | 1078 | 137 | 3.26 | 5.50×10^-4^ |
| *ALCAM* | ENSG00000170017 | 3 | 105050753 | 105305744 | 1510 | 109 | 0.31 | .38 | 1743 | 129 | -0.72 | .76 |
| *POLN* | ENSG00000130997 | 4 | 2063645 | 2278848 | 1606 | 111 | 1.09 | .14 | 1605 | 112 | 0.91 | .18 |
| *UNC5C* | ENSG00000182168 | 4 | 96073655 | 96505357 | 3260 | 368 | -0.48 | .68 | 418 | 57 | 2.34 | .01 |
| *FABP2* | ENSG00000145384 | 4 | 120228405 | 120278545 | 409 | 28 | 1.82 | .03 | 435 | 33 | 2.80 | .003 |
| *WDR70* | ENSG00000082068 | 5 | 37344314 | 37763537 | 2974 | 106 | -0.84 | .80 | 2969 | 108 | -0.80 | .79 |
| *PFDN1* | ENSG00000113068 | 5 | 139614624 | 139717706 | 565 | 90 | -0.34 | .63 | 562 | 116 | -0.36 | .64 |
| *HBEGF* | ENSG00000113070 | 5 | 139702428 | 139761216 | 330 | 110 | -0.50 | .69 | 329 | 110 | -0.61 | .73 |
| *NSG2* | ENSG00000170091 | 5 | 173437607 | 173680504 | 1778 | 239 | -0.53 | .70 | 1776 | 249 | -0.39 | .65 |
| *MSX2* | ENSG00000120149 | 5 | 174116536 | 174167896 | 366 | 93 | 0.37 | .36 | 365 | 91 | 0.97 | .17 |
| *IYD* | ENSG00000009765 | 6 | 150655028 | 150737105 | 817 | 100 | -2.93 | 1.00 | 900 | 115 | -1.76 | .96 |
| *PLEKHG1* | ENSG00000120278 | 6 | 150885999 | 151174799 | 2220 | 221 | -3.16 | 1.00 | 2510 | 247 | -1.02 | .85 |
| *SDK1* | ENSG00000146555 | 7 | 3306080 | 4318632 | 14031 | 354 | -2.08 | .98 | 14007 | 343 | -1.78 | .96 |
| *COBL* | ENSG00000106078 | 7 | 51073909 | 51419515 | 2763 | 192 | 0.91 | .18 | 2765 | 186 | 0.04 | .48 |
| *AKAP9* | ENSG00000127914 | 7 | 91535181 | 91749987 | 1126 | 61 | 0.24 | .41 | 1124 | 60 | 0.56 | .29 |
| *TSRM* | ENSG00000236294 | 7 | 113056127 | 113101457 | 298 | 74 | 2.77 | .003 | 298 | 74 | 2.99 | .001 |
| *CNTNAP2* | ENSG00000174469 | 7 | 145778453 | 148128090 | 22730 | 795 | 1.07 | .14 | 22665 | 807 | 1.16 | .12 |
| *MMP16* | ENSG00000156103 | 8 | 89034237 | 89375254 | 2357 | 206 | -1.35 | .91 | 2347 | 205 | -0.57 | .71 |
| *ZC3H3* | ENSG00000014164 | 8 | 144509825 | 144658623 | 1623 | 124 | 0.95 | .17 | 1604 | 129 | 0.81 | .21 |
| *MPDZ* | ENSG00000107186 | 9 | 13095703 | 13314589 | 1723 | 149 | 0.27 | .39 | 1719 | 158 | 1.09 | .14 |
| *USP6NL* | ENSG00000148429 | 10 | 11485945 | 11688753 | 1450 | 101 | -0.95 | .83 | 1453 | 104 | -0.41 | .66 |
| *ECHDC3* | ENSG00000134463 | 10 | 11749365 | 11816069 | 720 | 60 | 0.25 | .40 | 720 | 60 | -1.11 | .87 |
| *API5* | ENSG00000166181 | 11 | 43298513 | 43376079 | 486 | 92 | 0.30 | .38 | 480 | 85 | 0.19 | .42 |
| *ARAP1* | ENSG00000186635 | 11 | 72386114 | 72539644 | 1113 | 144 | 2.96 | .002 | 1115 | 152 | 2.57 | .005 |

**Supplementary Table 7 continued.** Gene-based results for AD genes previously identified in African Americans but not NHW^18,28-32,34-36^

| Gene | ID | Chr | Start BP  (hg37) | Stop BP  (hg37) | Model 1 | | | | Model 2 | | | |
| --- | --- | --- | --- | --- | --- | --- | --- | --- | --- | --- | --- | --- |
|  |  |  |  |  | No. of SNPs | No. of Parameters | *Z* | *P* | No. of SNPs | No. of Parameters | *Z* | *P* |
| *STARD10* | ENSG00000214530 | 11 | 72455774 | 72539726 | 558 | 101 | 2.93 | .002 | 560 | 109 | 2.46 | .007 |
| *ACER3* | ENSG00000078124 | 11 | 76536911 | 76747841 | 1123 | 56 | 1.19 | .12 | 1125 | 55 | 1.18 | .12 |
| *PIK3C2G* | ENSG00000139144 | 12 | 18365548 | 18811348 | 3904 | 226 | 1.22 | .11 | 3900 | 227 | 1.37 | .09 |
| *TMTC1* | ENSG00000133687 | 12 | 29643773 | 29972692 | 2965 | 297 | 0.77 | .22 | 2952 | 284 | 0.41 | .34 |
| *ASCL1* | ENSG00000139352 | 12 | 103316464 | 103364294 | 440 | 76 | -1.90 | .97 | 438 | 76 | -0.85 | .80 |
| *RNF6* | ENSG00000127870 | 13 | 26696253 | 26831791 | 1287 | 171 | -1.22 | .89 | 1287 | 183 | -1.15 | .87 |
| *ENOX1* | ENSG00000120658 | 13 | 43777654 | 44396044 | 4512 | 257 | -1.40 | .92 | 4509 | 264 | -1.30 | .90 |
| *GPC6* | ENSG00000183098 | 13 | 93844095 | 95069655 | 9843 | 544 | -0.07 | .53 | 9815 | 544 | -0.34 | .63 |
| *STK24* | ENSG00000102572 | 13 | 99092455 | 99265194 | 1647 | 135 | 1.31 | .10 | 1639 | 134 | 0.76 | .22 |
| *SLC10A2* | ENSG00000125255 | 13 | 103686350 | 103754196 | 614 | 122 | 1.09 | .14 | 613 | 122 | 0.99 | .16 |
| *ARRDC4* | ENSG00000140450 | 15 | 98427784 | 98527068 | 1043 | 145 | -1.36 | .91 | 1040 | 143 | -1.15 | .87 |
| *IGF1R* | ENSG00000140443 | 15 | 99157200 | 99517759 | 3176 | 317 | -1.06 | .86 | 3168 | 310 | -0.89 | .81 |
| *RBFOX1* | ENSG00000078328 | 16 | 6034095 | 7773340 | 28264 | 952 | -1.11 | .87 | 28188 | 953 | -1.05 | .85 |
| *CNTNAP4* | ENSG00000152910 | 16 | 76276176 | 76603135 | 3994 | 249 | 0.43 | .34 | 3977 | 243 | 0.29 | .39 |
| *TANC2* | ENSG00000170921 | 17 | 61051917 | 61515060 | 2574 | 103 | -0.26 | .60 | 2573 | 105 | 0.34 | .37 |
| *SPHK1* | ENSG00000176170 | 17 | 74337665 | 74393941 | 389 | 70 | 3.72 | .0001 | 387 | 69 | 3.39 | 3.47×10^-4^ |
| *GRIN3B* | ENSG00000116032 | 19 | 965418 | 1019731 | 647 | 112 | 2.38 | .009 | 648 | 110 | 3.01 | .001 |
| *HMHA1* | ENSG00000180448 | 19 | 1030922 | 1096627 | 702 | 117 | 3.33 | 4.32×10^-4^ | 700 | 108 | 3.95 | 3.91×10^-5^ |
| *NFIC* | ENSG00000141905 | 19 | 3324561 | 3479215 | 1182 | 207 | 0.46 | .32 | 1178 | 200 | -0.91 | .82 |
| *VRK3* | ENSG00000105053 | 19 | 50469724 | 50564203 | 1011 | 27 | 0.18 | .43 | 1012 | 27 | 0.10 | .46 |

^a^Model 1 is adjusted for PCs, age, sex

^b^Model 2 is adjusted for PCs, age, sex, APOE genotype

### Supplementary Table 8. Sequence variants in ADSP Hispanic individuals within 1Mb distance to GWAS and CADD-PHRED score ≥20

| **GWAS Locus** | **Sequence Variant** | **Ref**  **Allele** | **Alt**  **Allele** | **AFF AC** | **AFF AN** | **UNAFF AC** | **UNAFF AN** | **MAF** | **χ^2^** | ***P*** | **Distance to top locus** | **D' with GWAS top variant** | **Consequence** | **SYMBOL** | **CADD_**  **Score** |
| --- | --- | --- | --- | --- | --- | --- | --- | --- | --- | --- | --- | --- | --- | --- | --- |
| *IYD/ PLEKHG1* | 6:151738529 | C | T | 7 | 4012 | 25 | 2082 | 0.005 | 25.71 | 3.97E-07 | 919,077 | 1 | missense_variant | *RMND1* | 24.4 |
| *MPDZ* | 9:12709125 | T | G | 35 | 4010 | 44 | 2082 | 0.013 | 15.52 | 8.15E-05 | 511,393 | 1 | stop_gained | *TYRP1* | 37 |
| *ASCL1* | 12:104380824 | A | C | 14 | 4010 | 29 | 2082 | 0.007 | 19.84 | 8.42E-06 | 810,451 |  | missense_variant | *TDG* | 21.5 |
| *CNTNAP4* | 16:75590013 | G | A | 17 | 4012 | 29 | 2072 | 0.008 | 16.07 | 6.12E-05 | 945,057 |  | missense_variant | *TMEM231* | 21.1 |
| *CNTNAP4* | 16:75575299 | T | C | 17 | 4010 | 29 | 2082 | 0.008 | 15.90 | 6.67E-05 | 959,771 |  | missense_variant | *TMEM231* | 21.4 |
| *TANC2* | 17:61611569 | A | C | 9 | 3928 | 33 | 1732 | 0.007 | 43.60 | 4.02E-11 | 322,711 |  | missense_variant | *KCNH6* | 24.7 |
| *TANC2* | 17:61512597 | A | C | 7 | 3980 | 26 | 1894 | 0.006 | 30.80 | 2.86E-08 | 223,739 |  | missense_variant | *CYB561* | 23 |
| *TANC2* | 17:60522268 | G | A | 2 | 4012 | 16 | 2082 | 0.003 | 21.66 | 3.26E-06 | 766,590 |  | missense_variant | *METTL2A* | 28.9 |
| *TANC2* | 17:62049954 | A | G | 7 | 4010 | 20 | 2078 | 0.004 | 17.50 | 2.87E-05 | 761,096 | 1 | missense_variant | *SCN4A* | 23 |

### Supplementary Figure 1. Regional association plots for the (A) two novel common and (B) ten rare loci identified in single-variant meta-analysis. The SNPs labeled on each regional plot had the lowest P value at each locus and are represented by a purple diamond. Each dot represents a SNP and dot colors indicate strength of LD with the labeled SNP. Blue vertical lines show the recombination rate marked on the right-hand y-axis of each regional plot.

**A)**

chr2:3070309 (rs78857220, Model1)

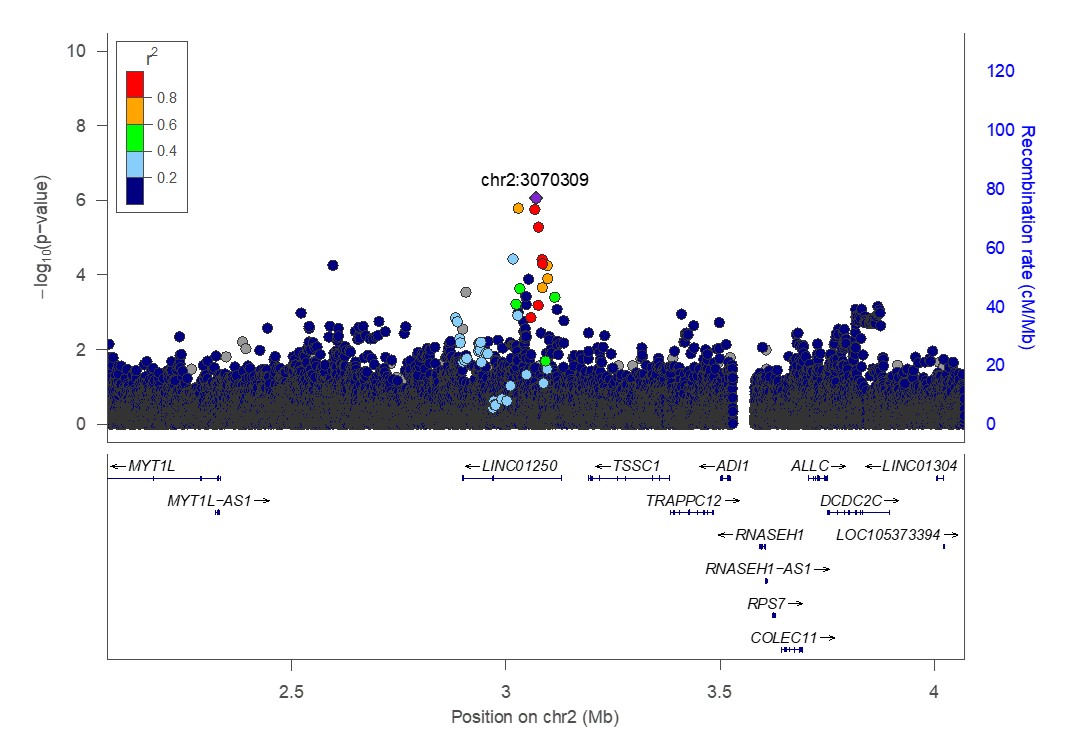

chr3:921529 (rs396323, Model 1)

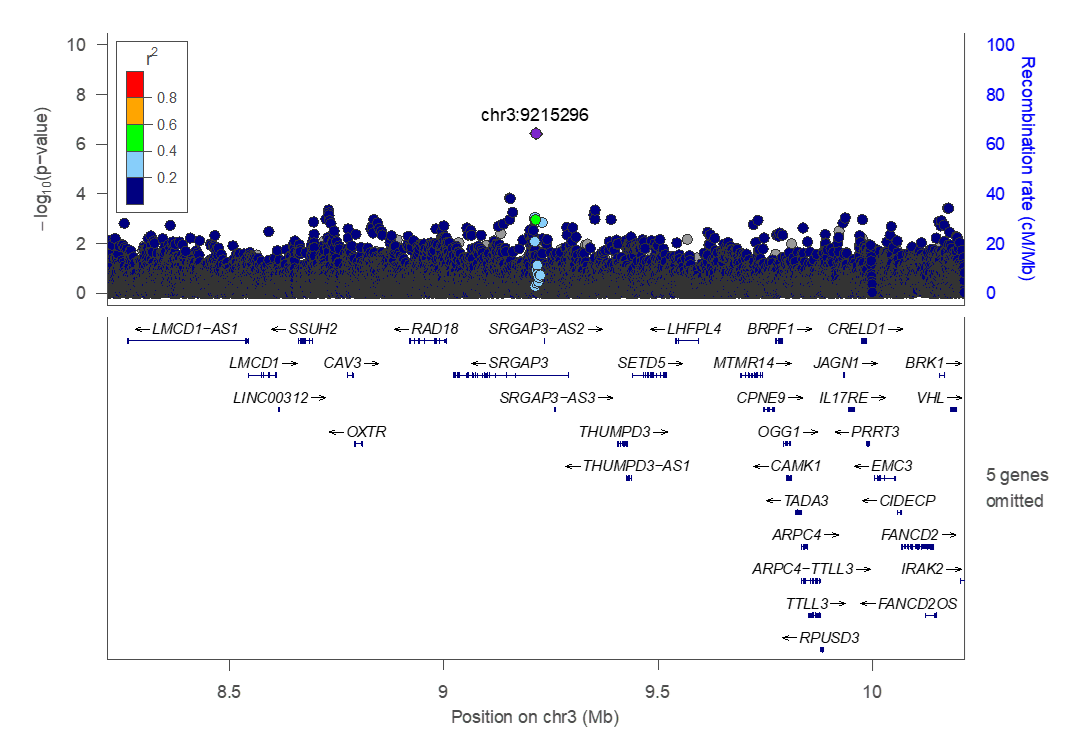

**B)**

chr2:8806098 (rs183088158, Model 2)

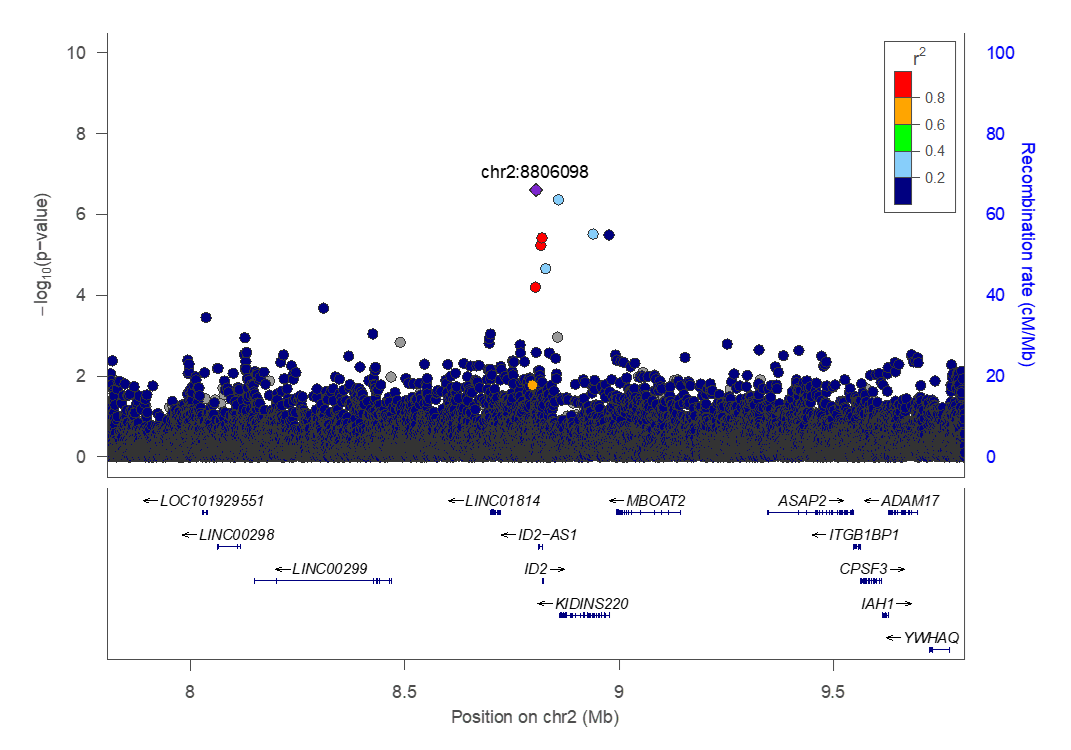

chr2:145902343 (rs191256330, Model 2)

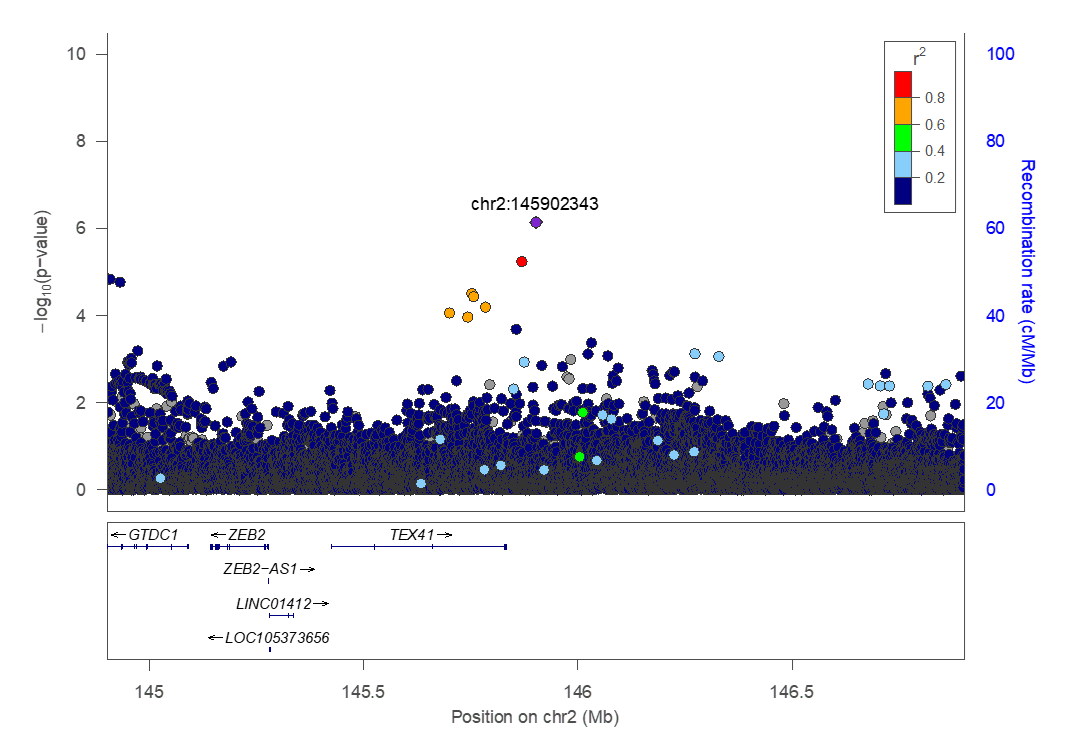

chr4:96471106 (rs969240869, Model 2)

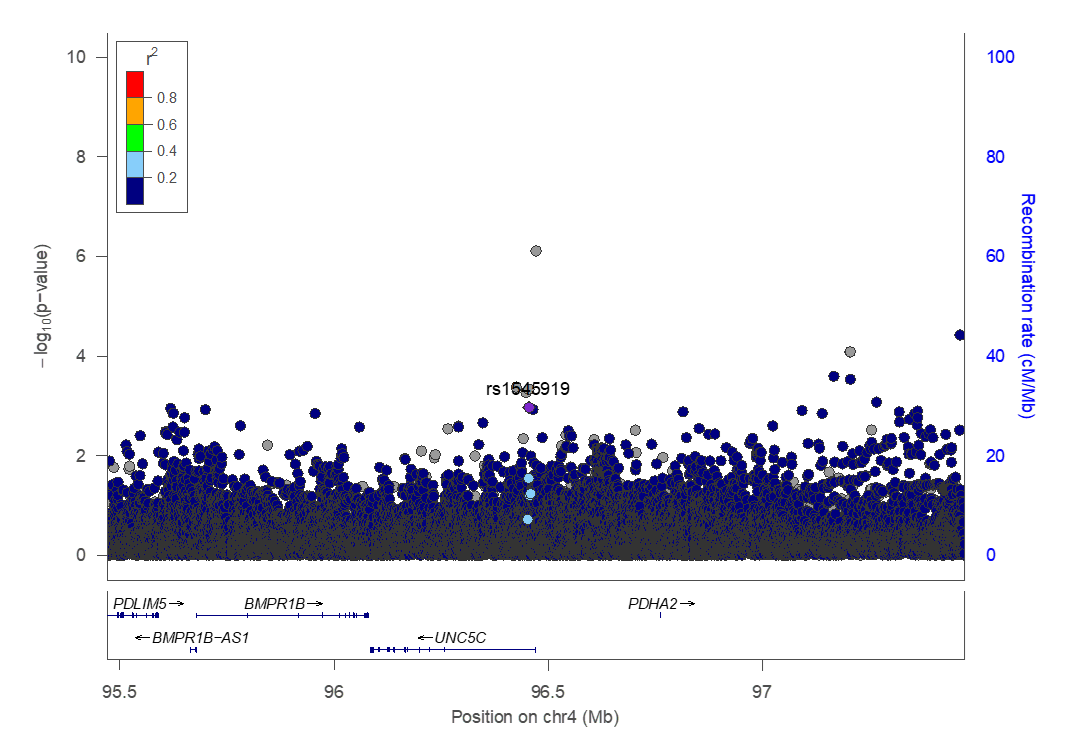

chr6:150819452 (rs117762284, Model 1)

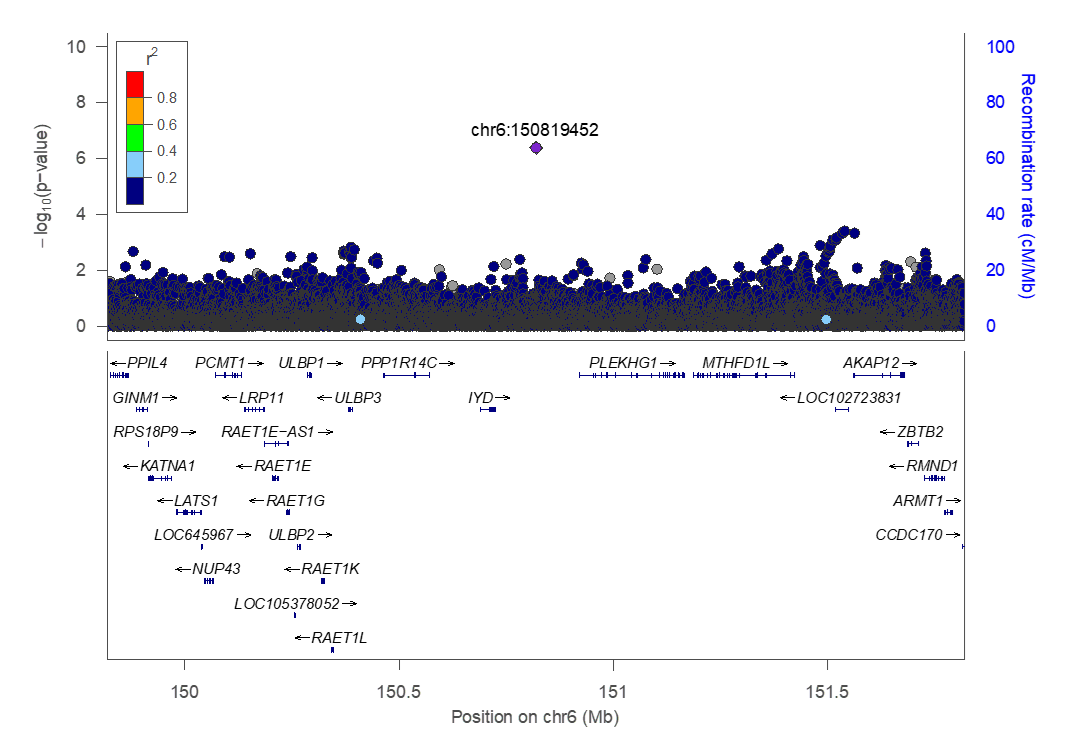

chr7:4268872 (rs181767458, Model 2)

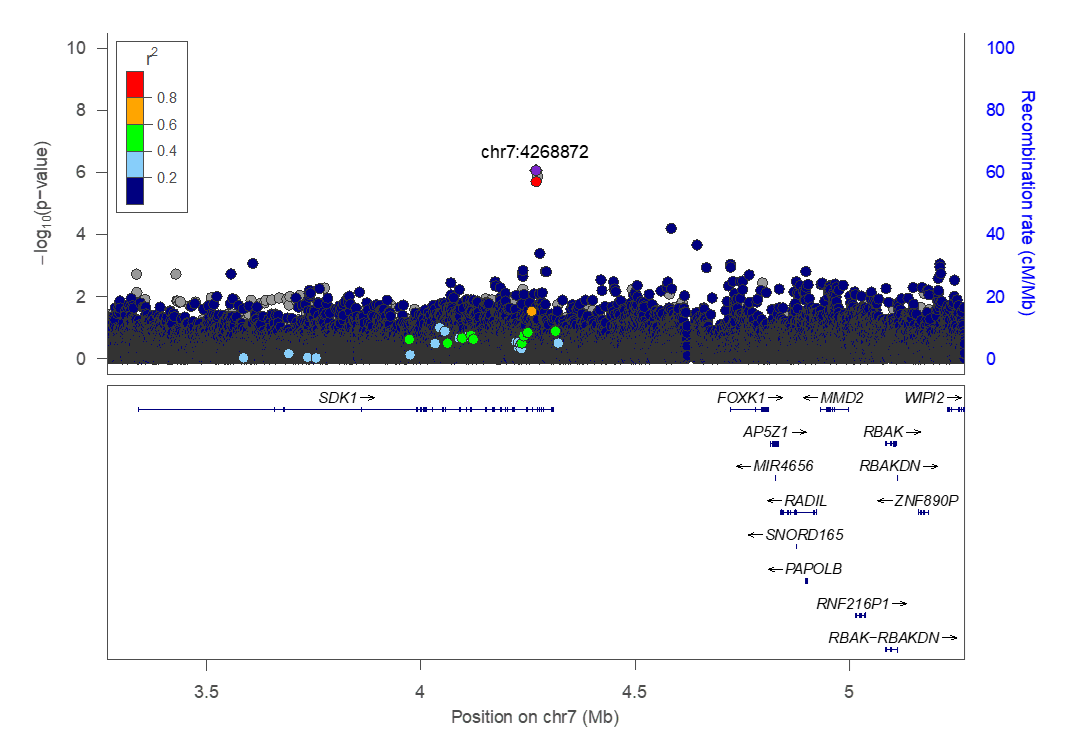

chr8:90157165 (rs138744190, Model 2)

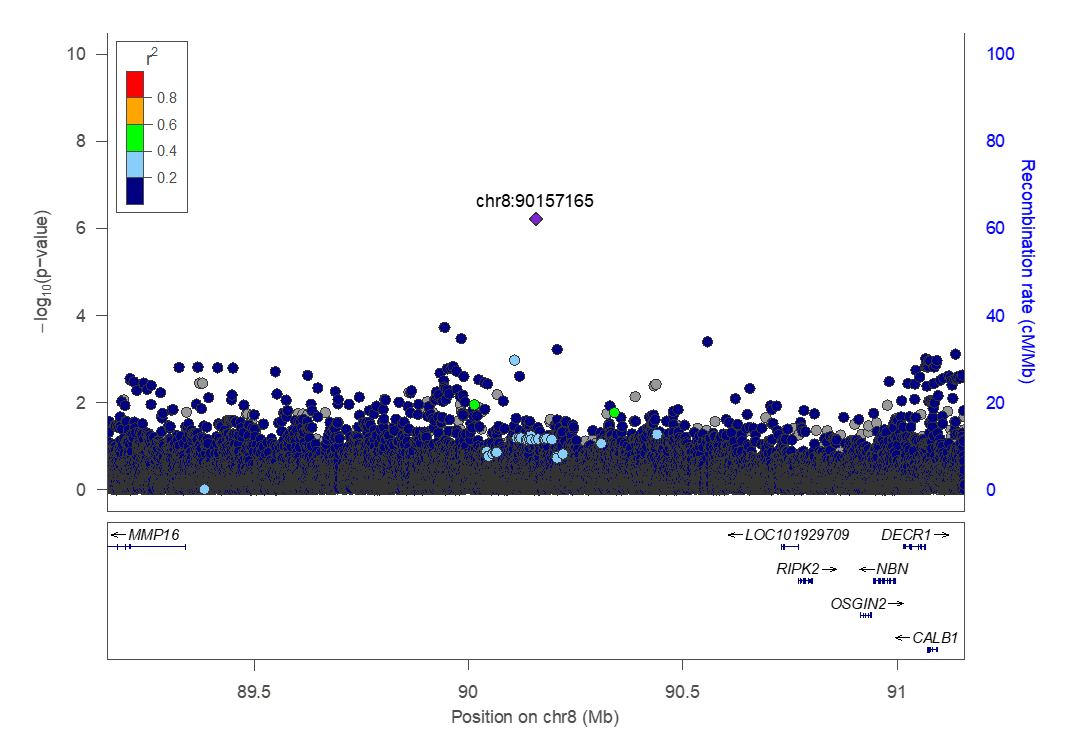

chr9:13220518 (rs141610415, Model 2)

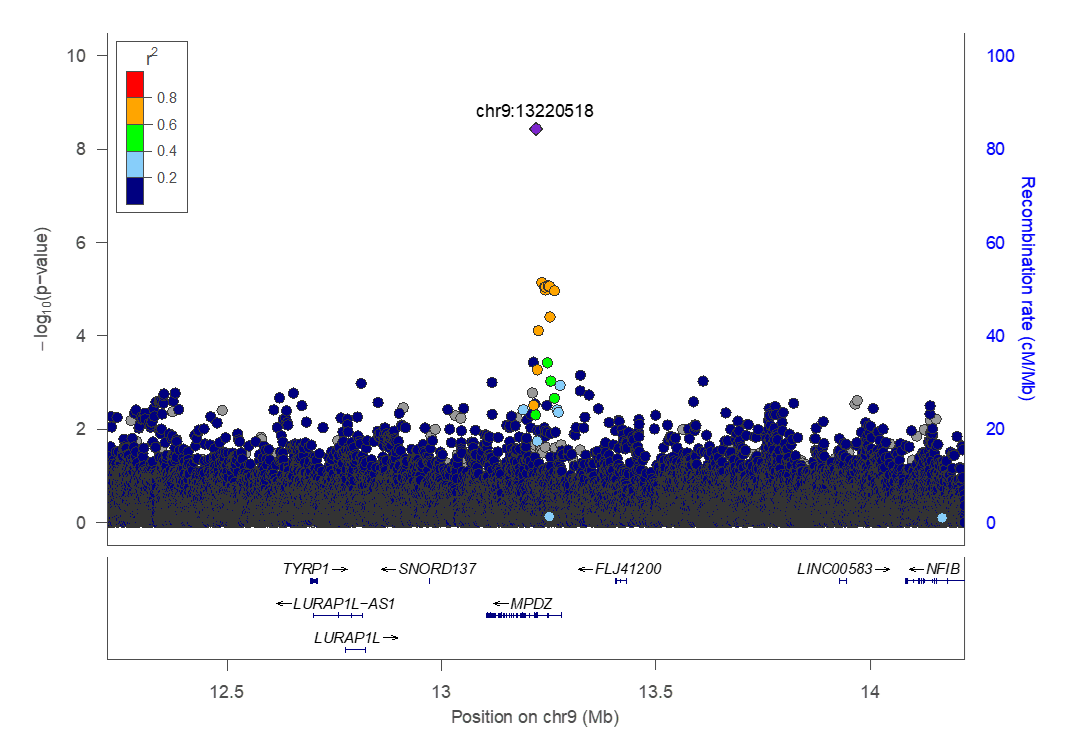

chr12:103570373 (rs556001137, Model 1)

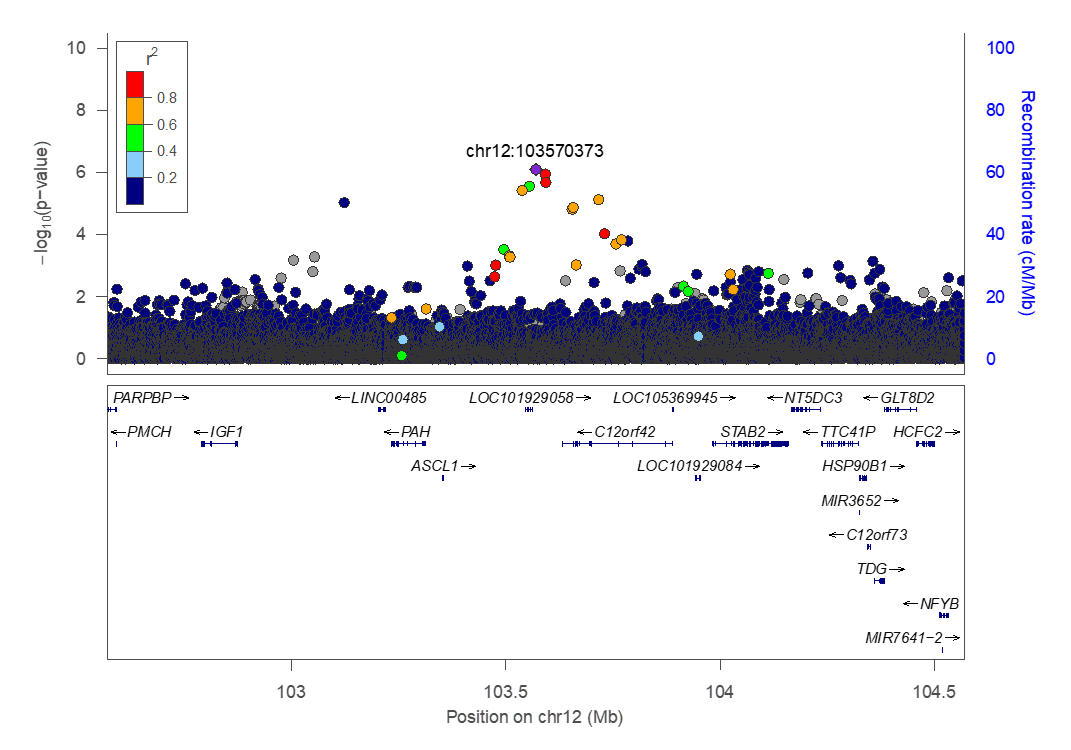

chr12:103716273 (rs138206541, Model 2)

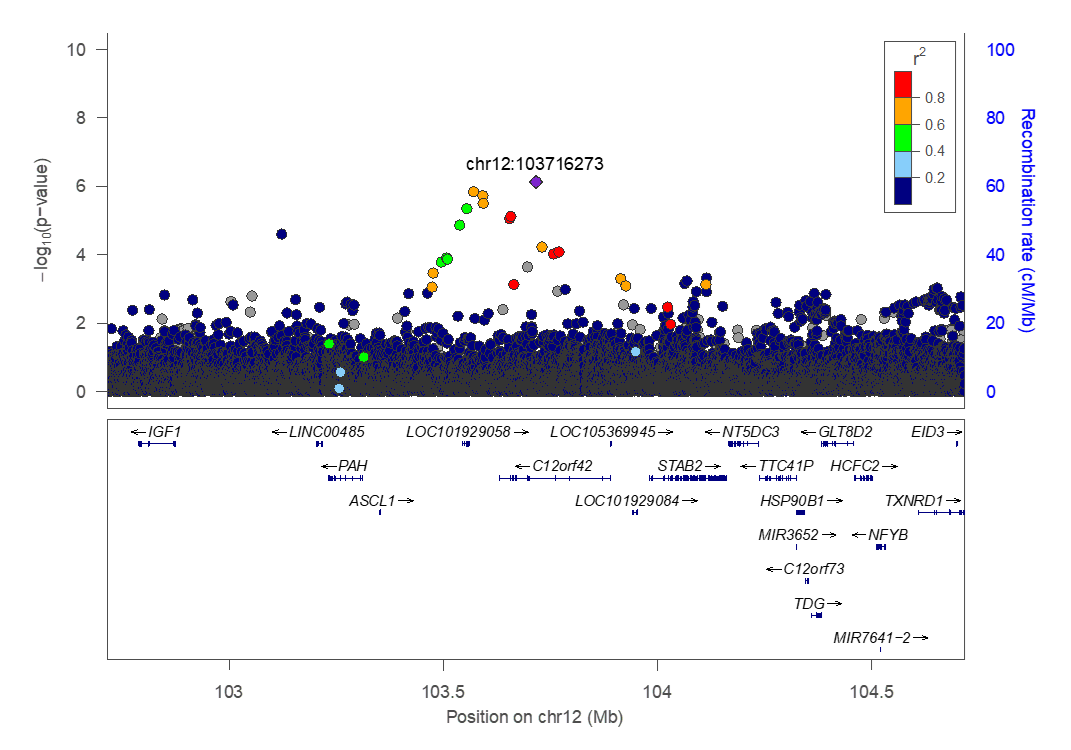

chr16:76535070 (rs1010752317, Model 2)

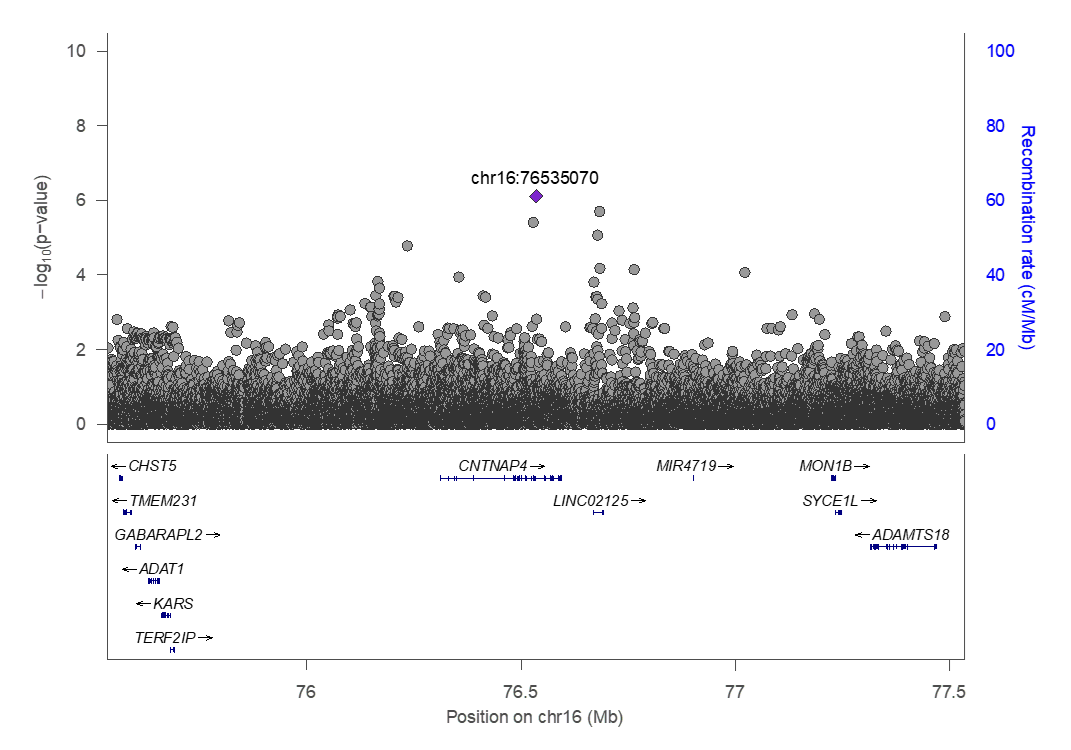

chr17:61288858 (rs56285182, Model 2)

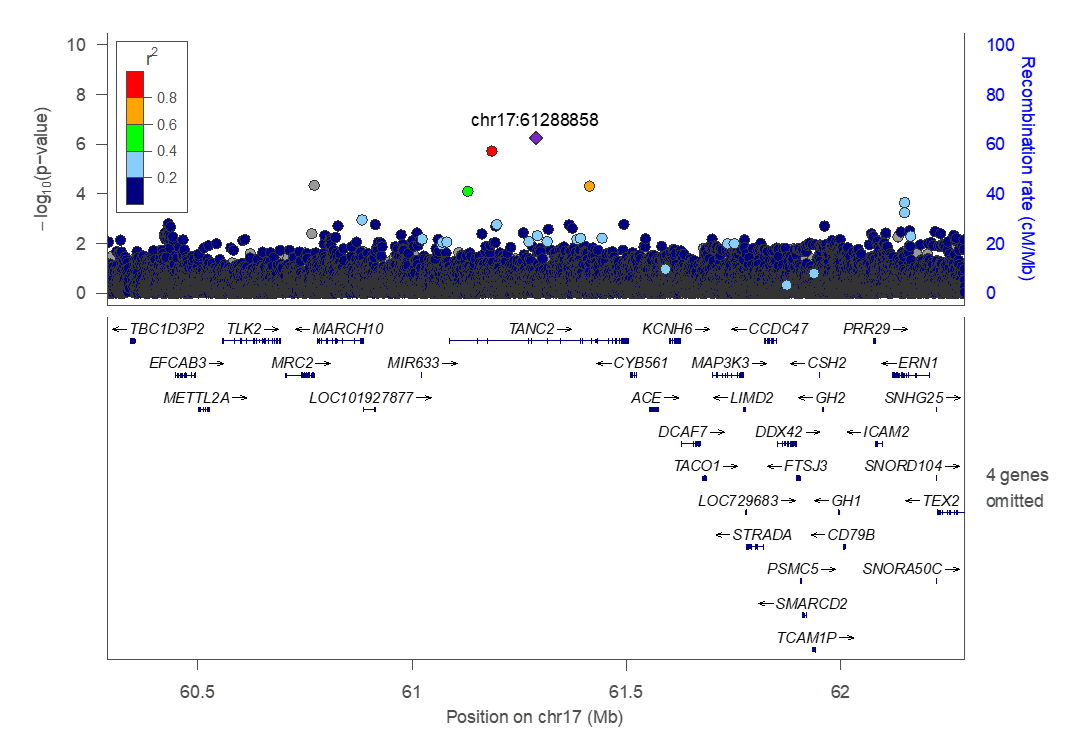

### Supplementary Figure 2. Forest Plots of Odds Ratios (ORs) and standard errors (SEs) for the (A) two novel common and (B) ten rare loci identified in single-variant meta-analysis

chr2:3070309 (rs78857220, Model1)

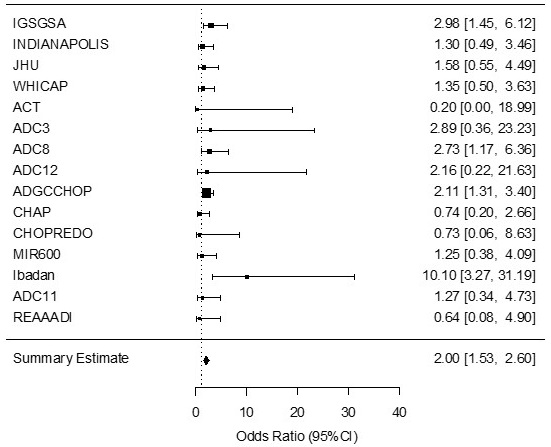

chr3:921529 (rs396323, Model 1)

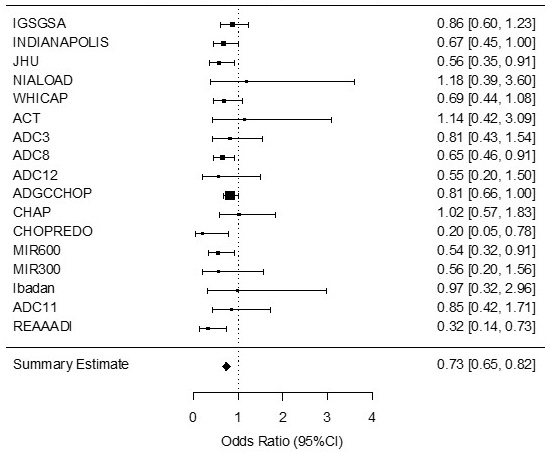

**B)**

chr2:8806098 (rs183088158, Model 2)

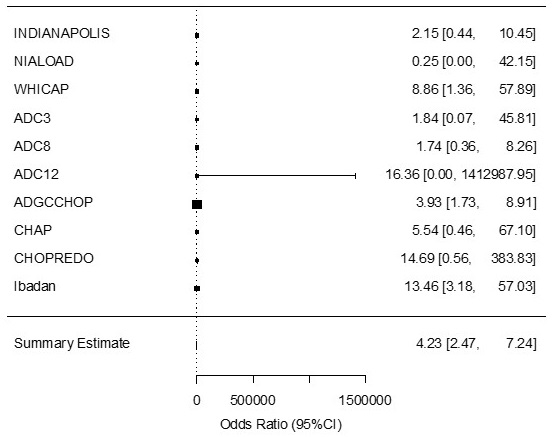

chr2:145902343 (rs191256330, Model 2)

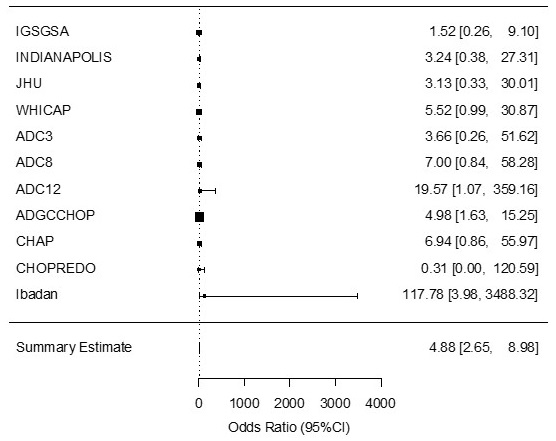

chr4:96471106 (rs969240869, Model 2)

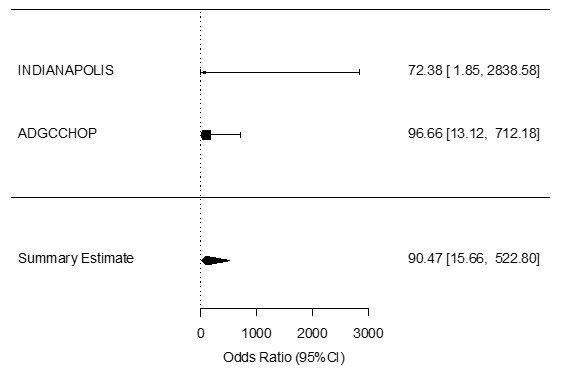

chr6:150819452 (rs117762284, Model 1)

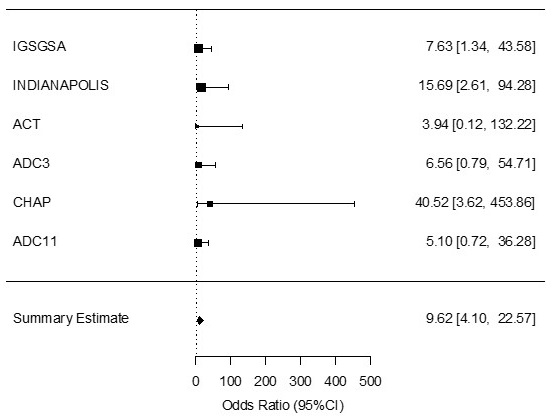

chr7:4268872 (rs181767458, Model 2)

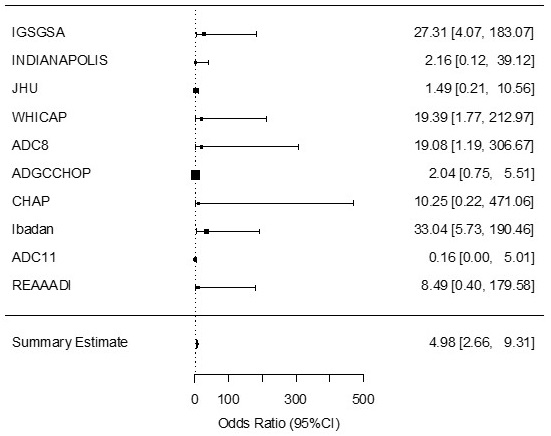

chr8:90157165 (rs138744190, Model 2)

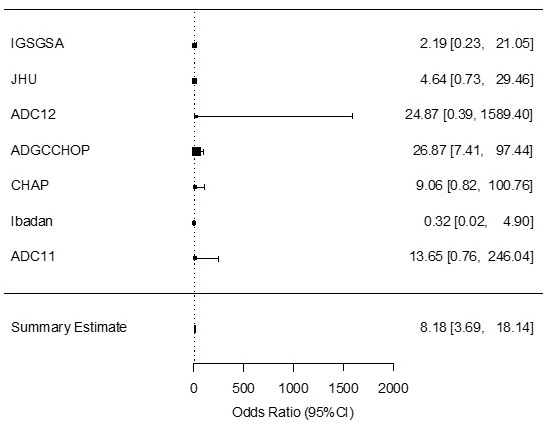

chr9:13220518 (rs141610415, Model 2)

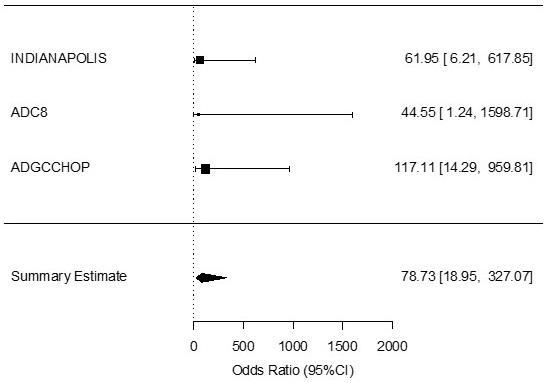

chr12:103570373 (rs556001137, Model 1)

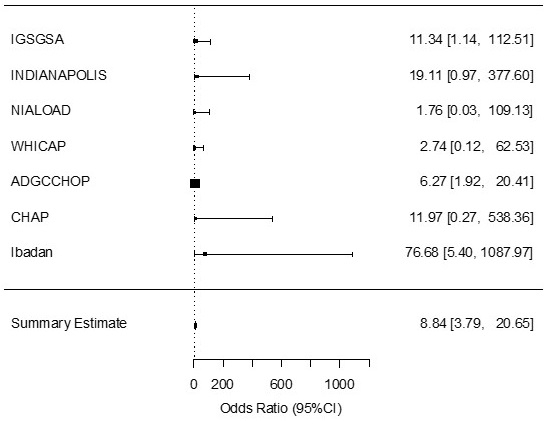

chr12:103716273 (rs138206541, Model 2)

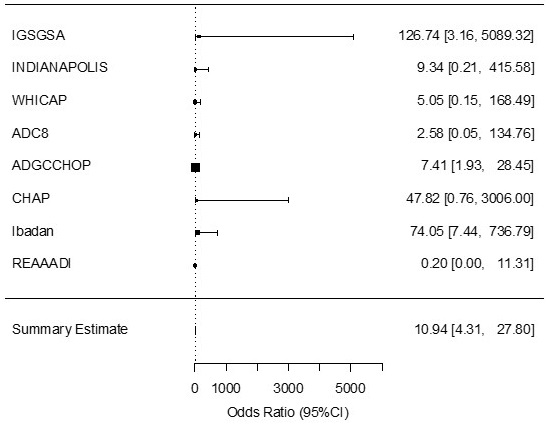

chr16:76535070 (rs1010752317, Model 2)

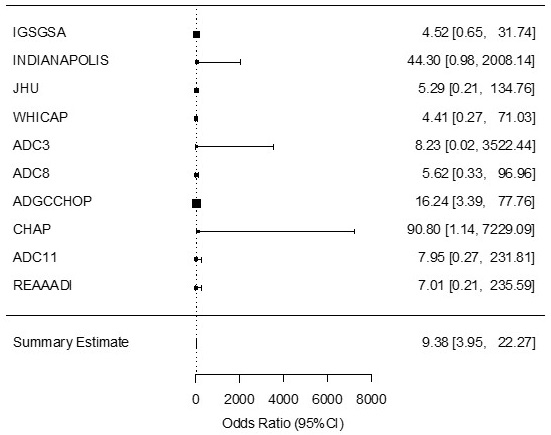

chr17:61288858 (rs56285182, Model 2)

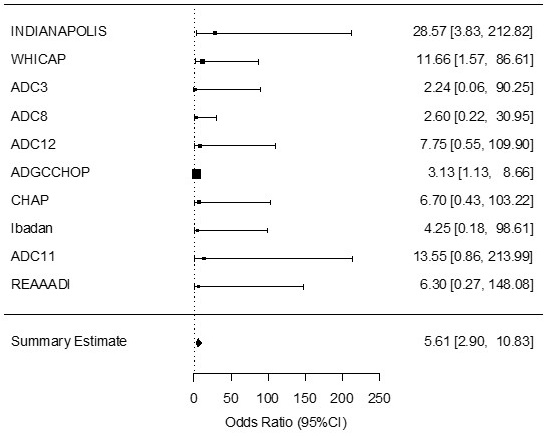

### Supplementary Figure 3. Quantile-quantile plots for single marker association analyses based (A) on the model adjusted for age, sex and population stratification and (B) age, sex, population stratification and *APOE* dosage showing the deviation of observed from expected p-values.

A) Model 1

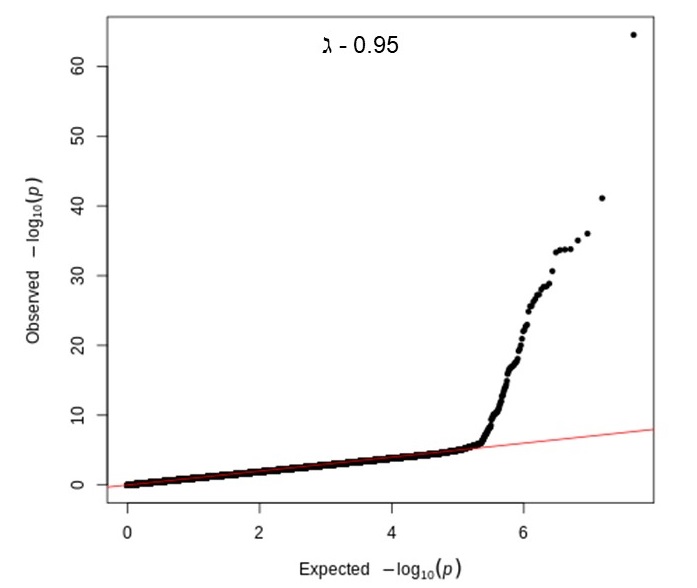

B) Model 2

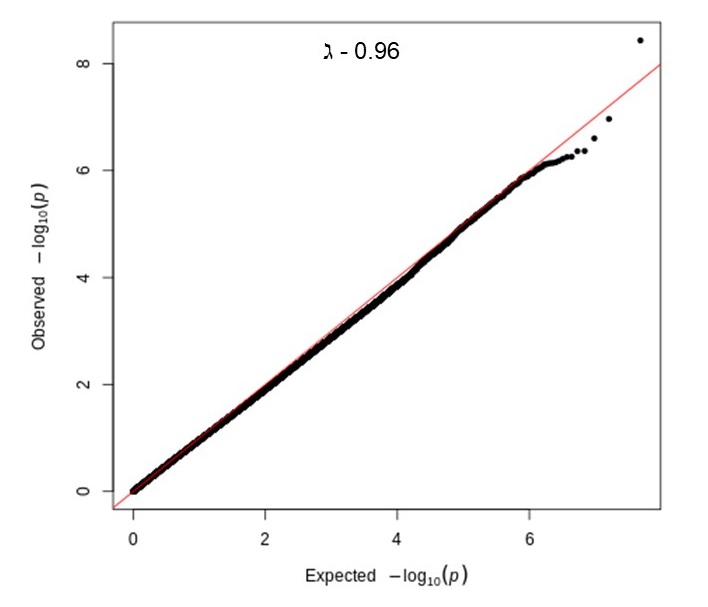

### Supplemental Figure 4. PCA plots for all individual datasets compared to data from the HapMap project^37^. CEU = Central European ancestry (from Utah, US); JPT_CHB = Japanese ancestry (from Tokyo) and Chinese ancestry (from Bejing); YRI = Yoruban ancestry (from Nigeria, West Africa).

**A)** ACT

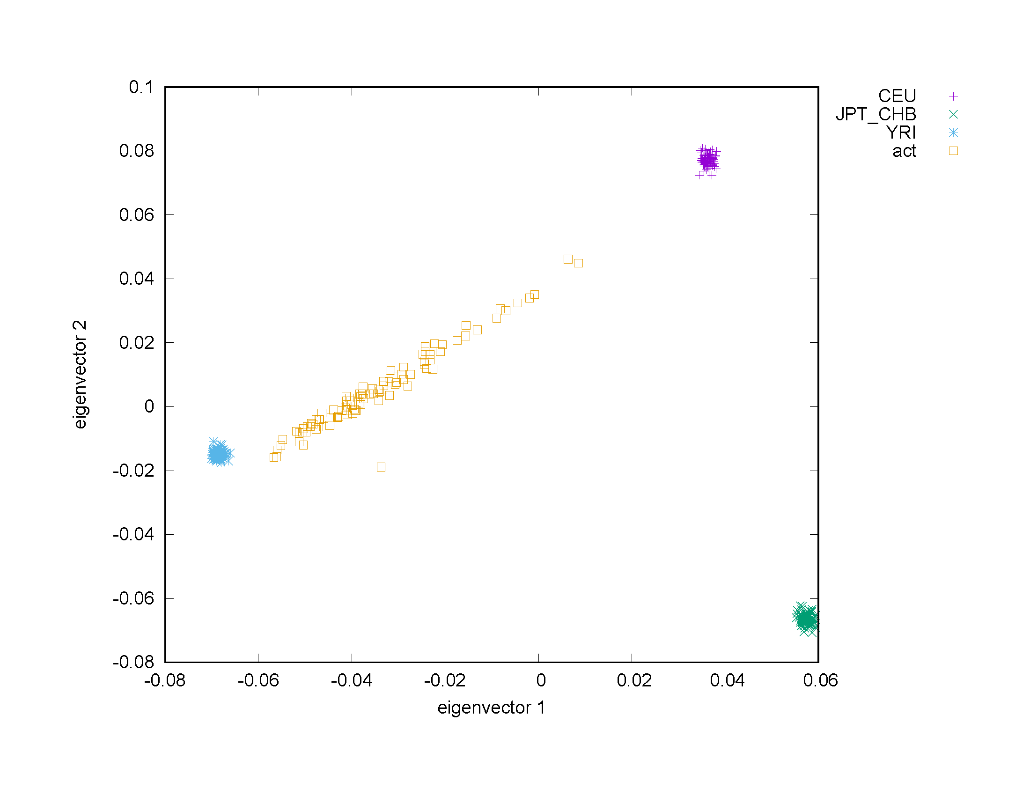

**B)** ADC 1/2

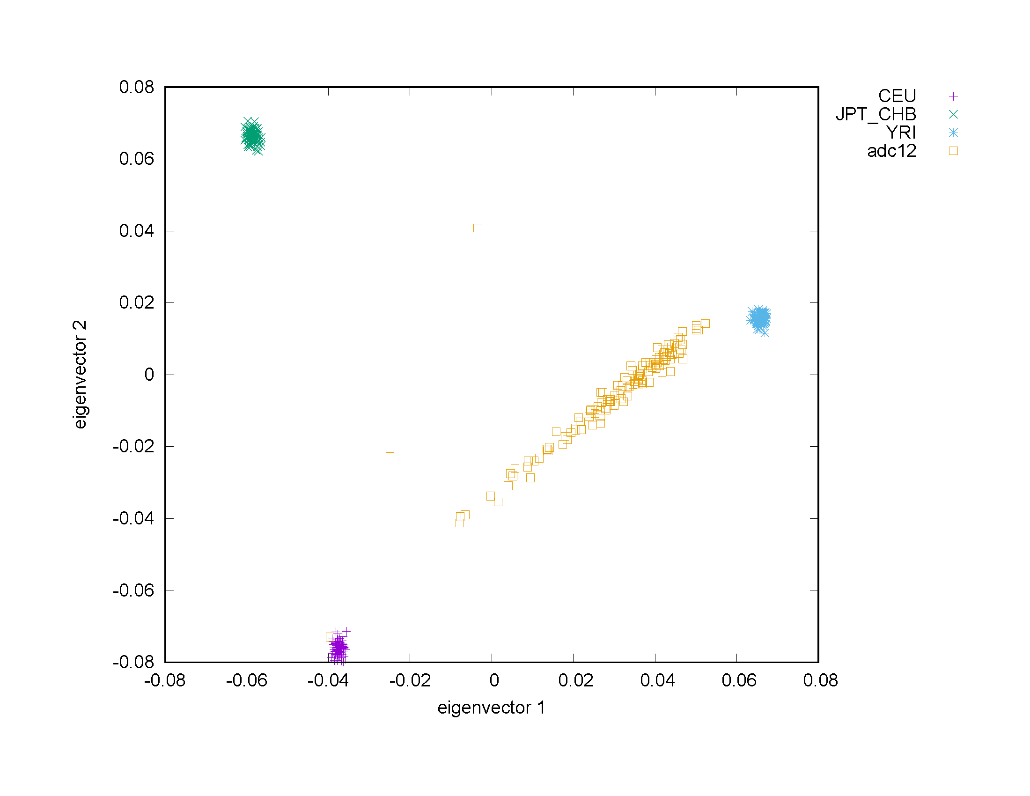

**C)** ADC 3

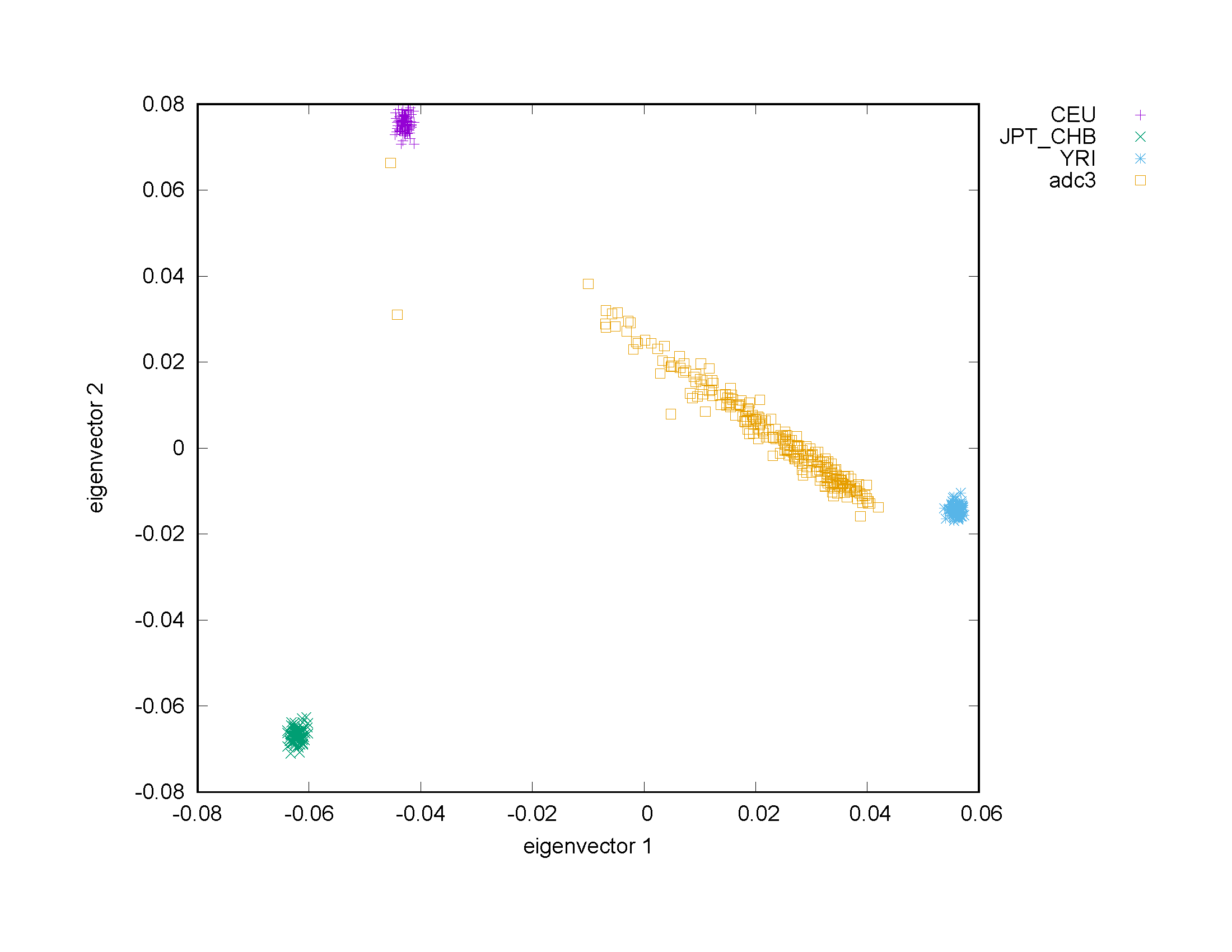

**D)** ADC 8

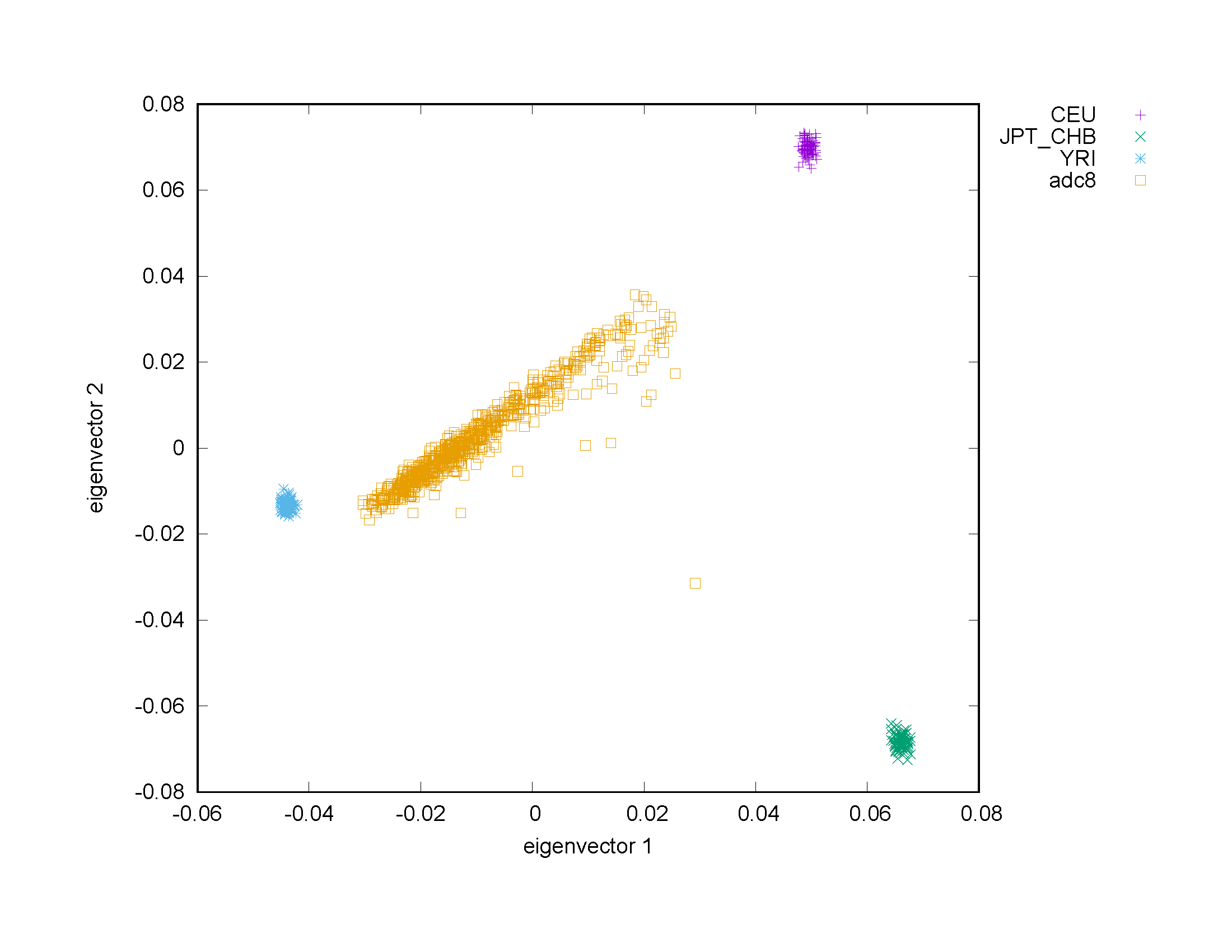

**E)** ADC 11

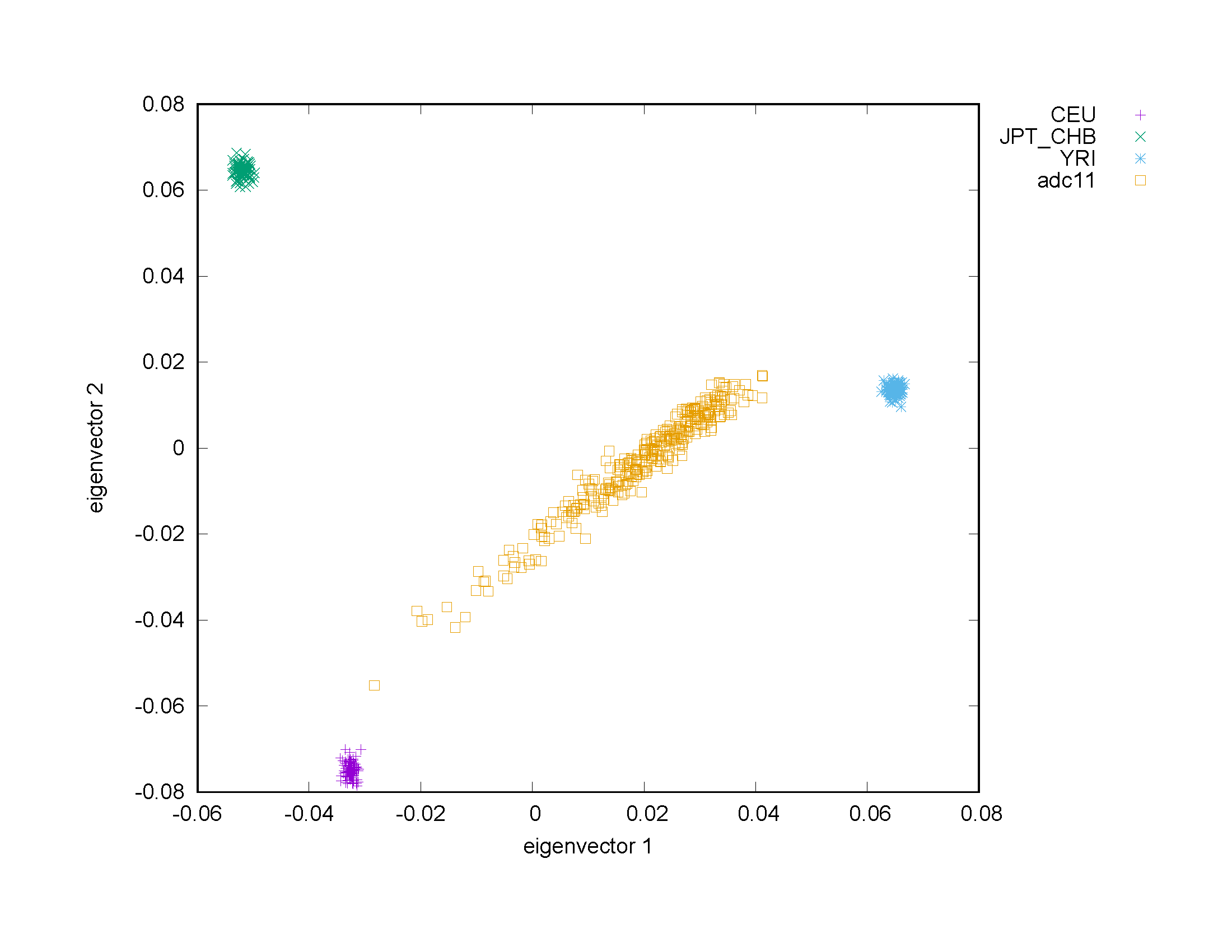

**F)** CHAP

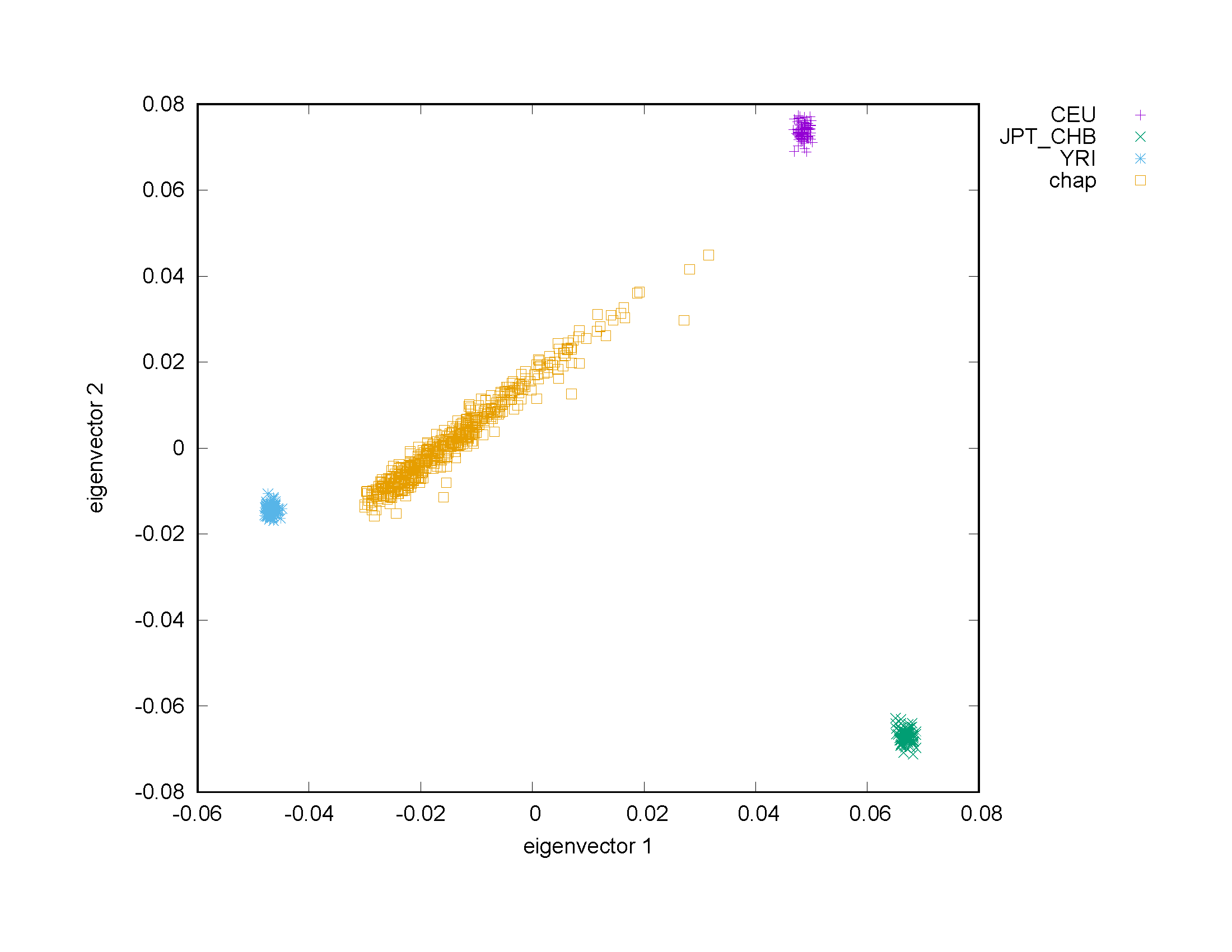

**G)** Indianapolis

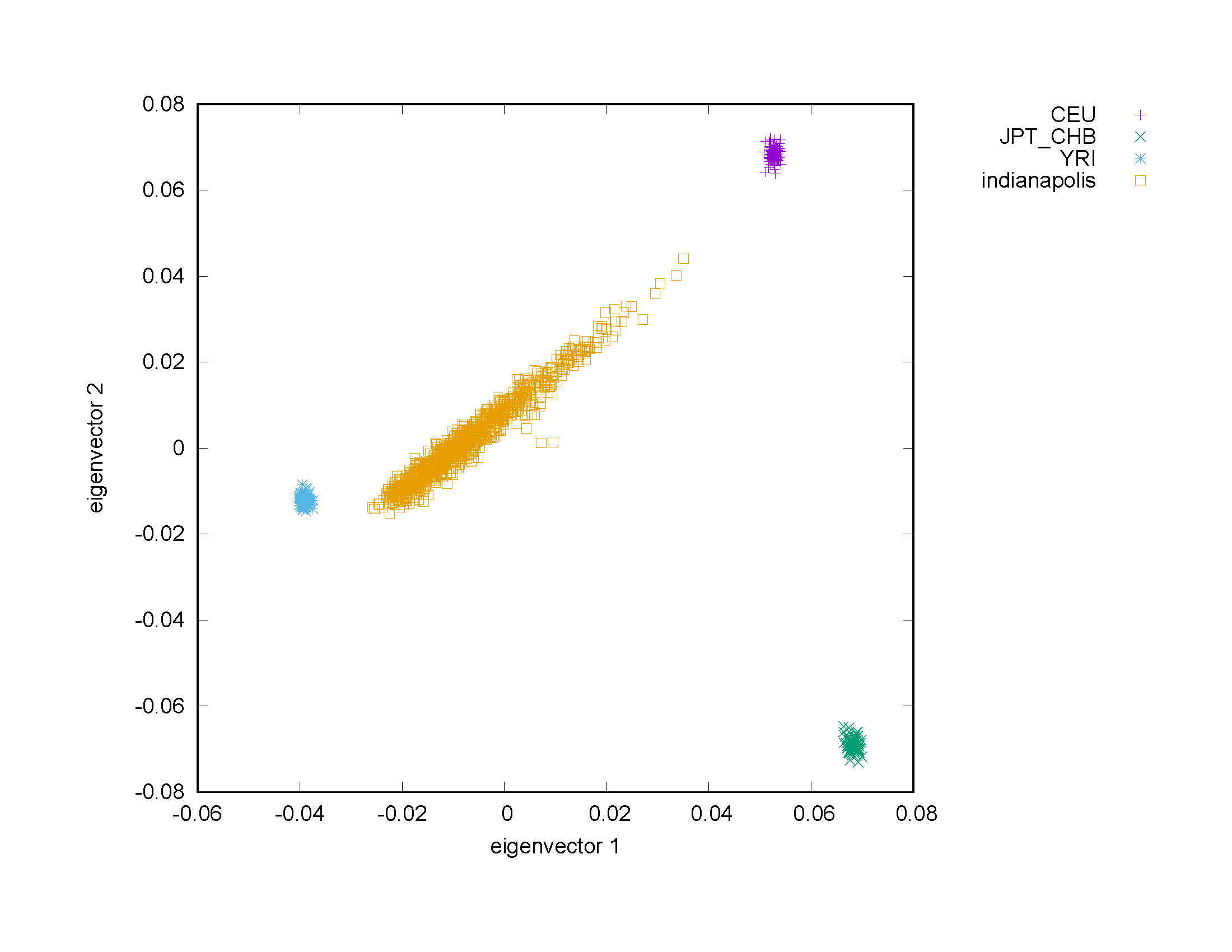

**H)** NIA-FBS/NCRAD

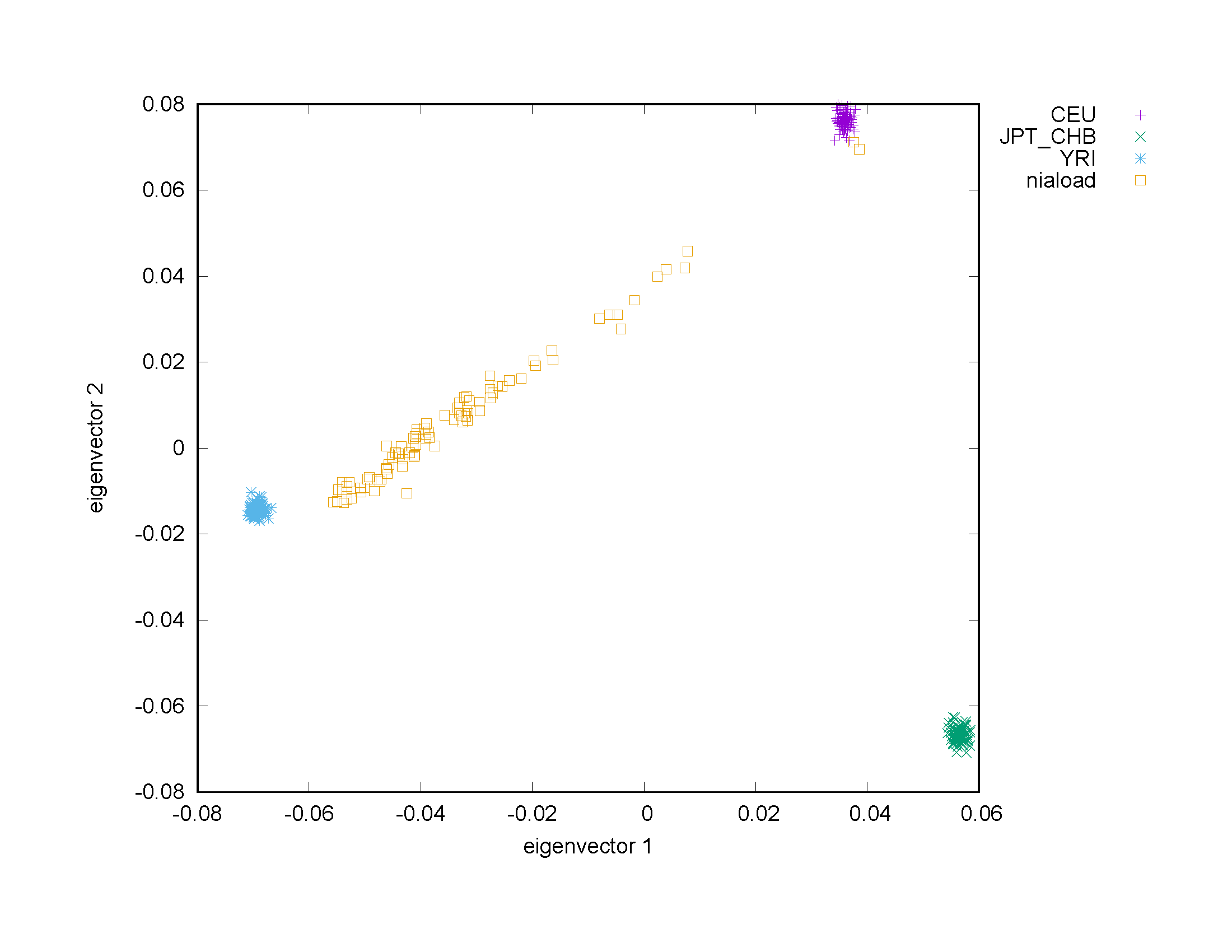

**I)** ADGC [2013]

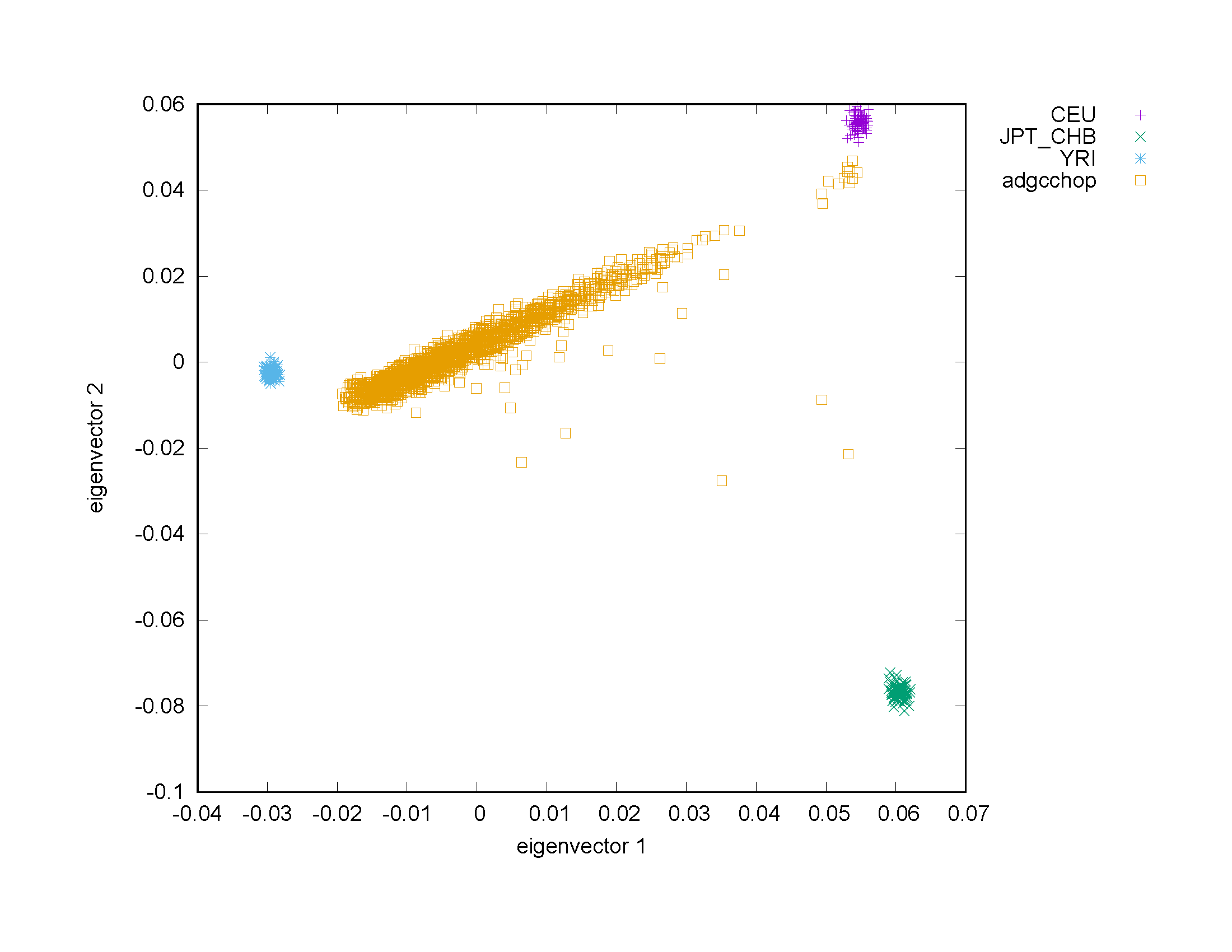

**J)** ADGC [2018a]

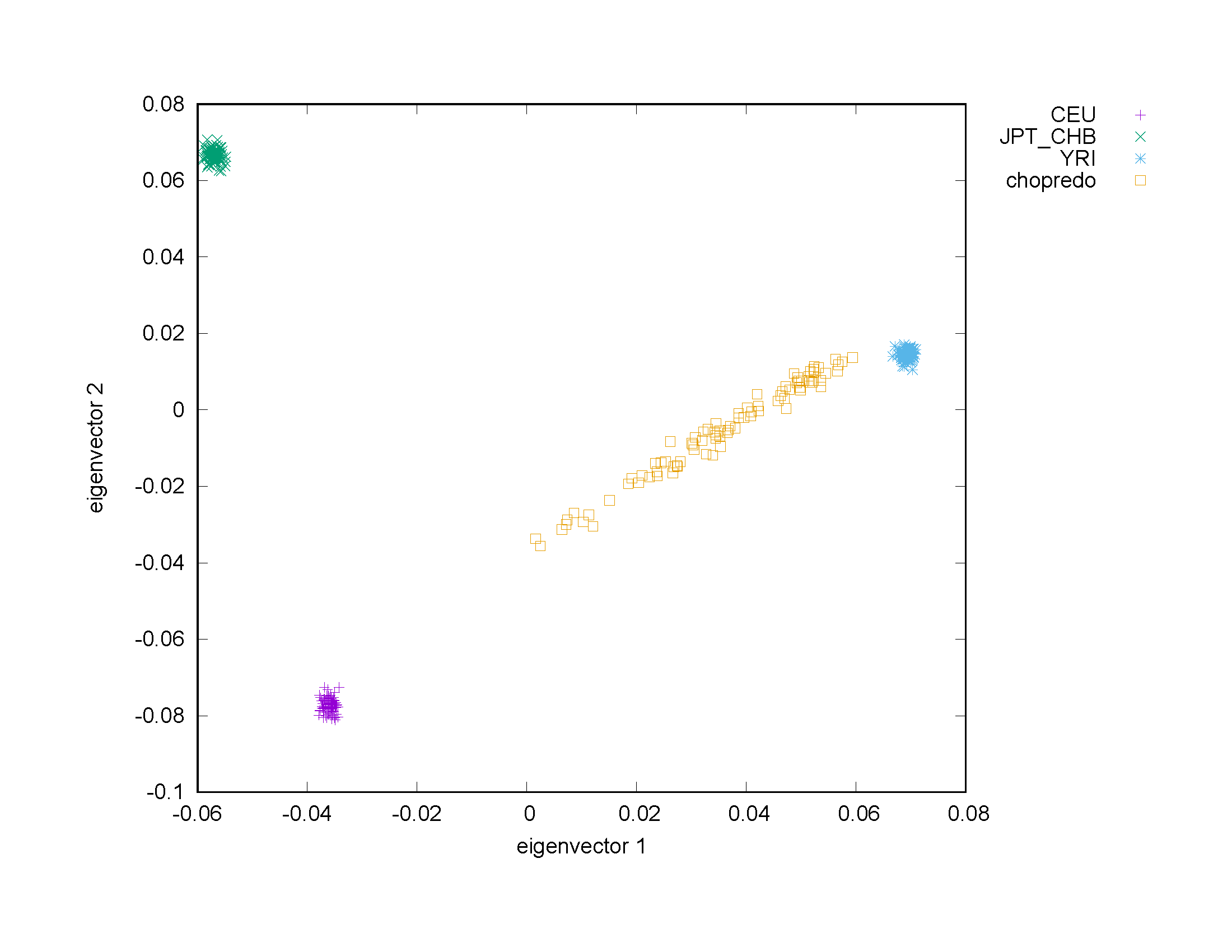

**K)** Mirage 300k

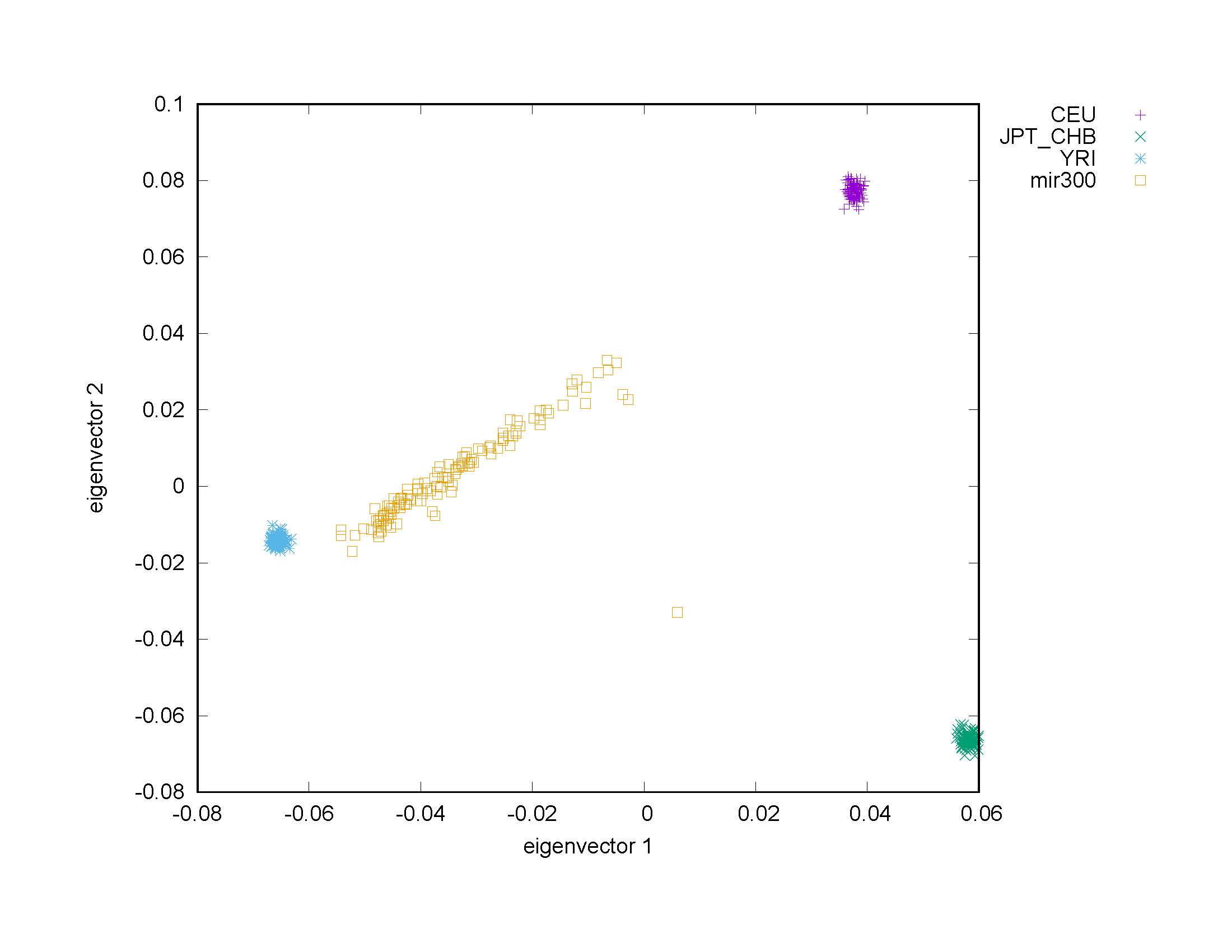

**L)** Mirage 660k

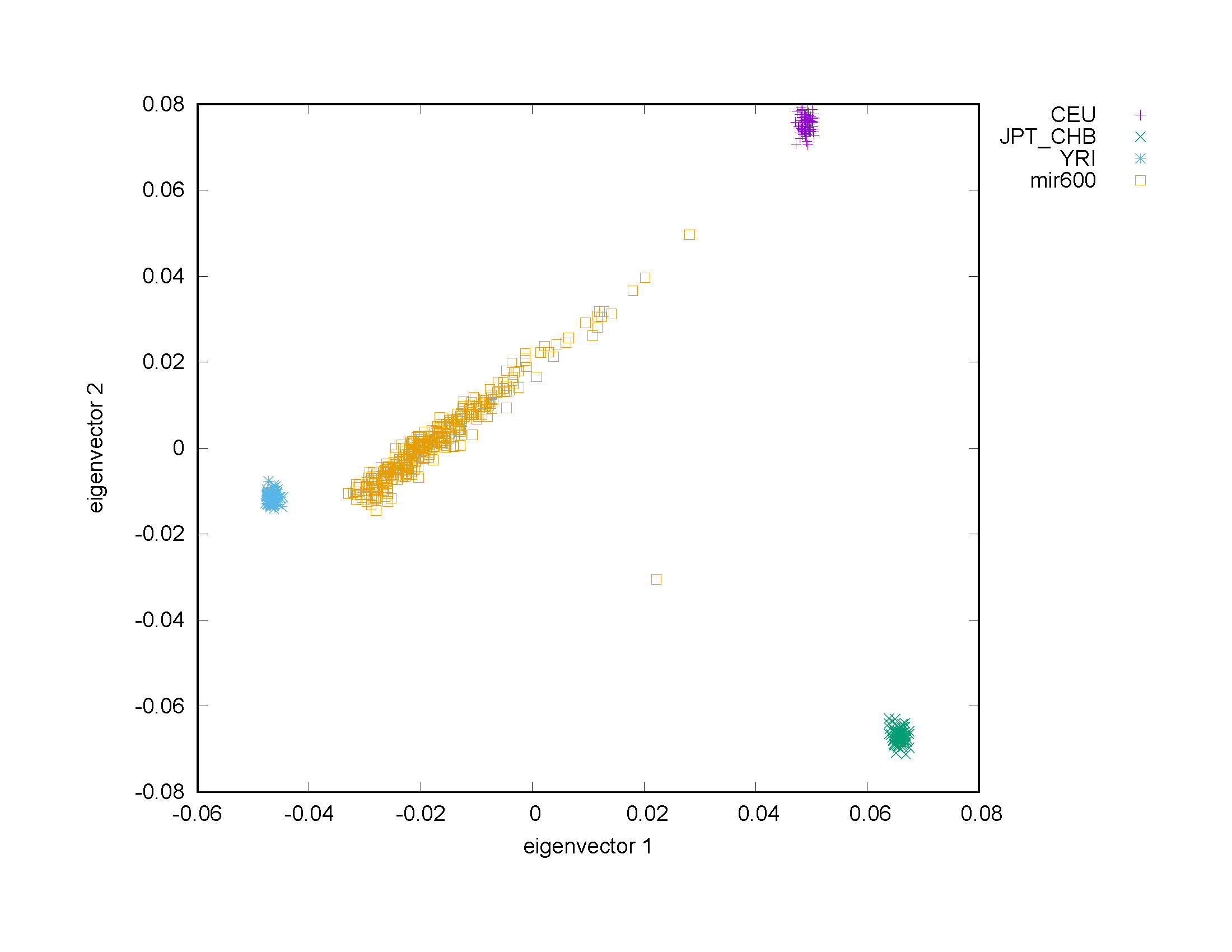

**M)** GenerAAtions

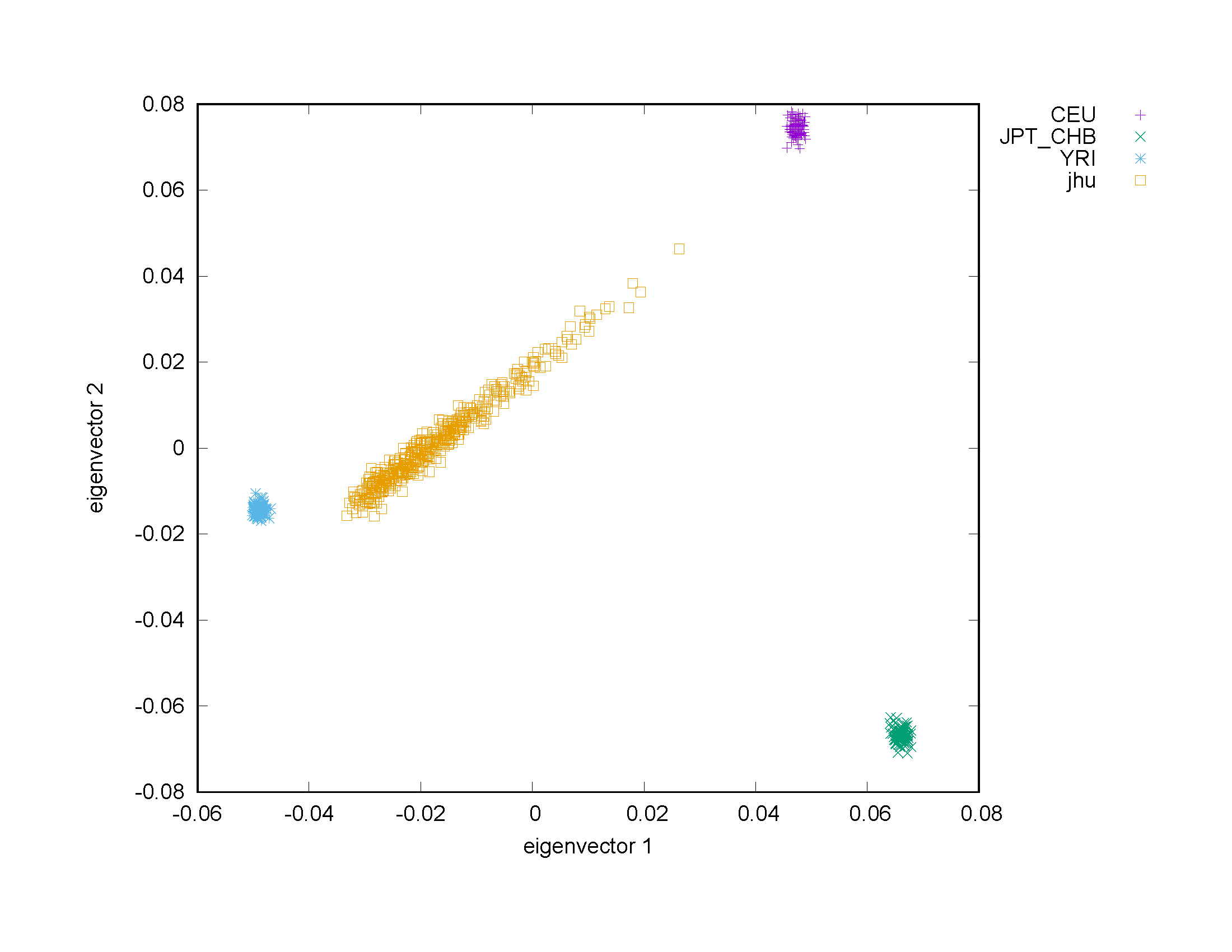

**N)** ADGC [2018b]

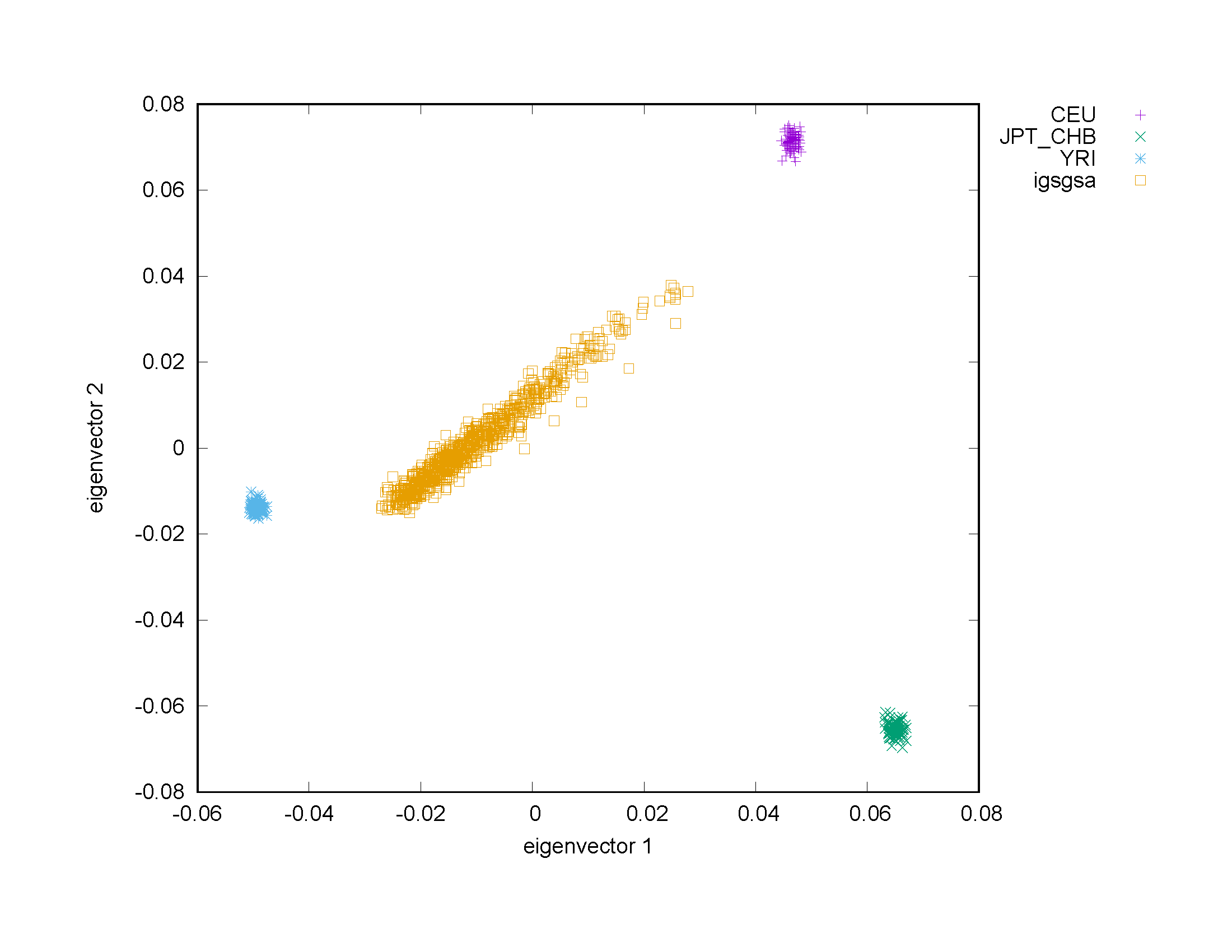

**O)** WHICAP

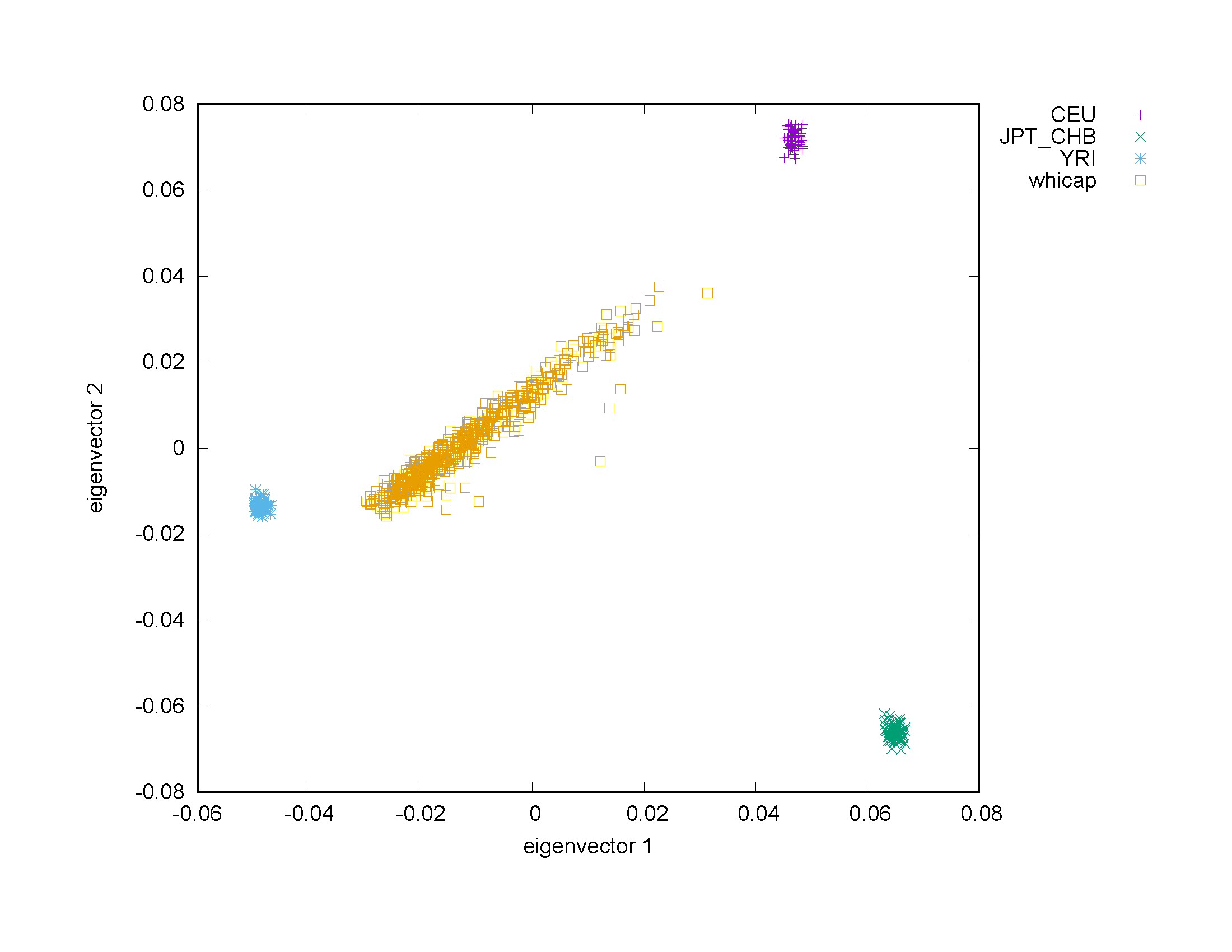

**P)** REAAADI

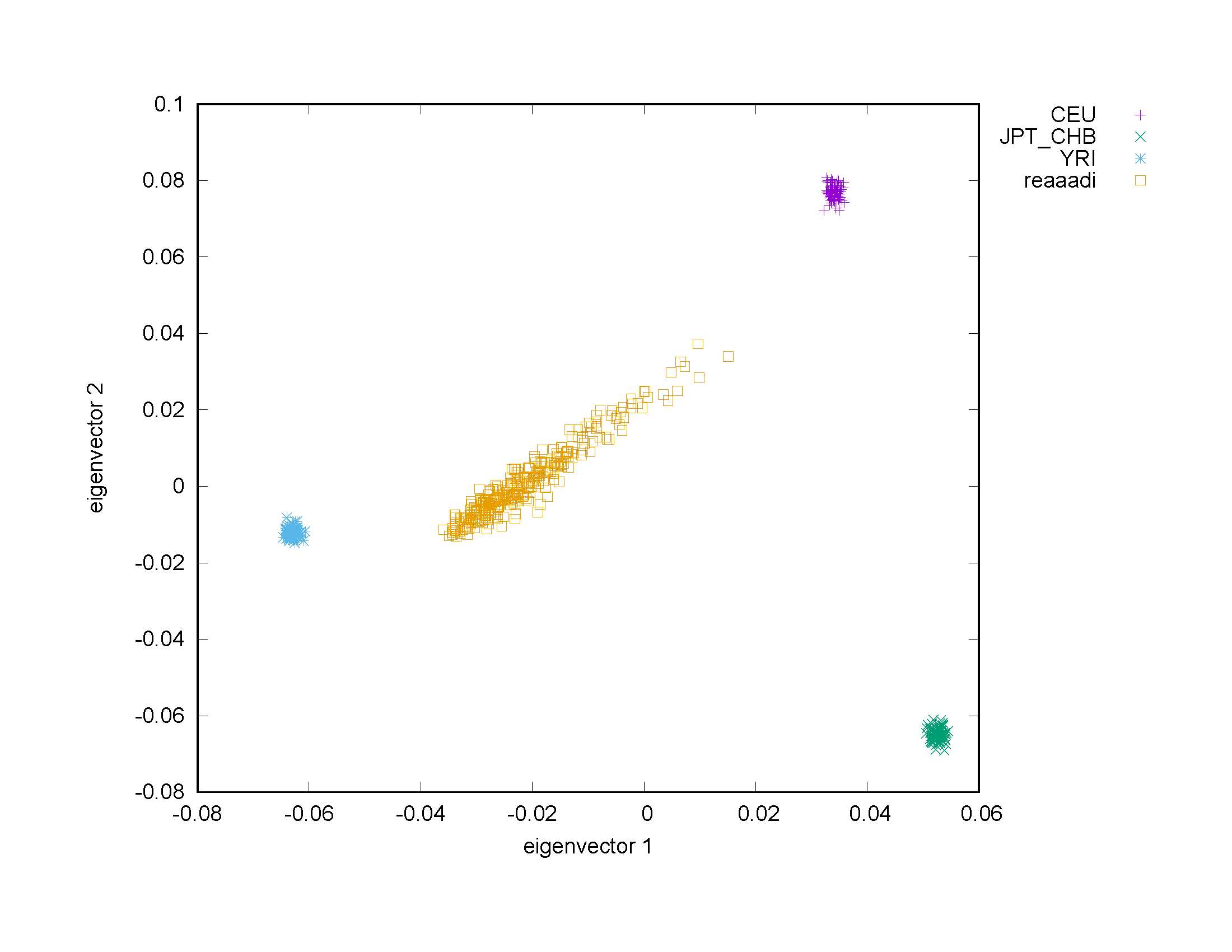

**Q)** Ibadan

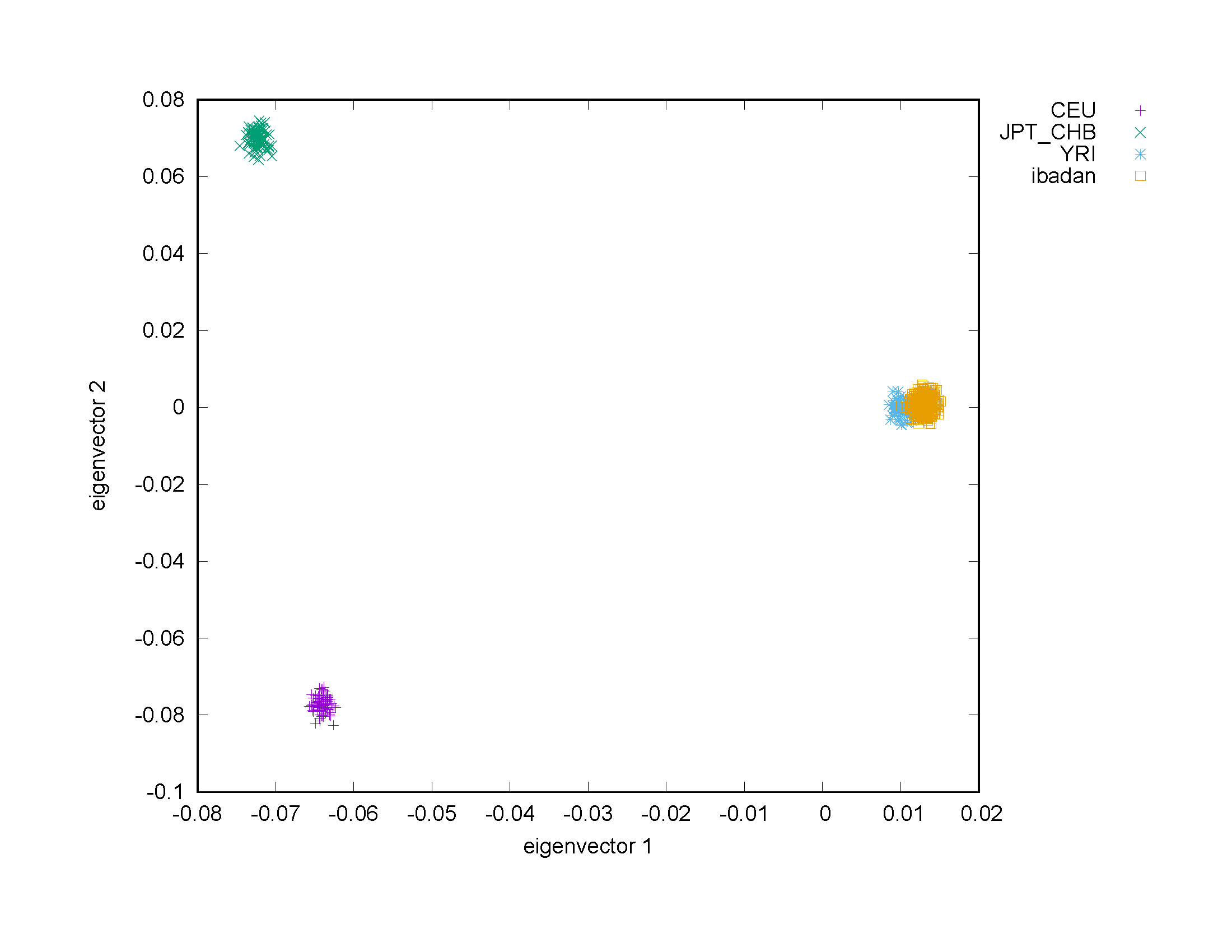

### Supplementary Figure 5. Individual locus zoom plots for A) chr2:3070309, B) chr2:145902343, and C) chr12:103570373 for the dataset with high African ancestry (e.g. Ibadan) and the African American datasets.

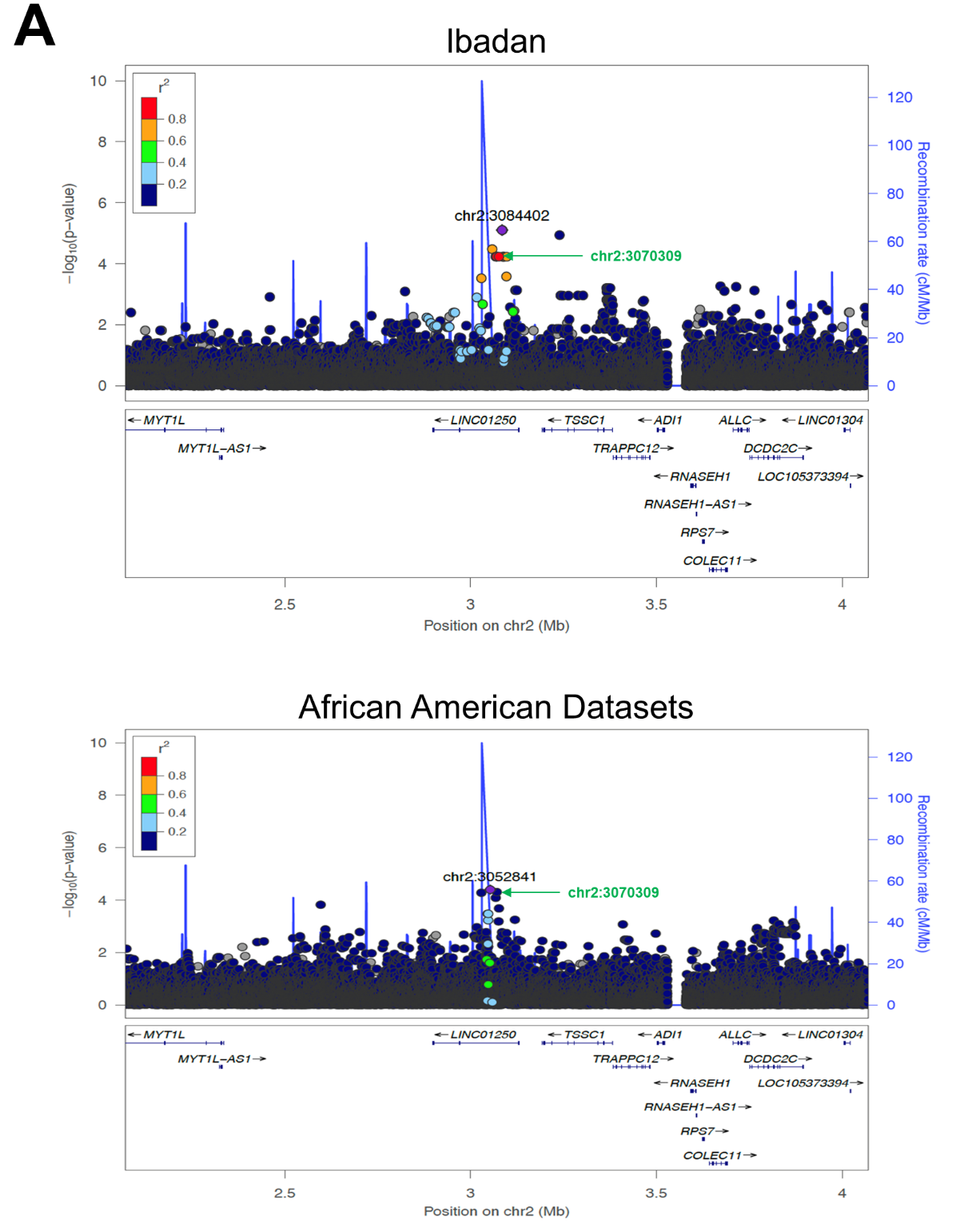

The top SNP reported in our results from the meta-analysis across all datasets (Ibadan and African American) is shown in green.

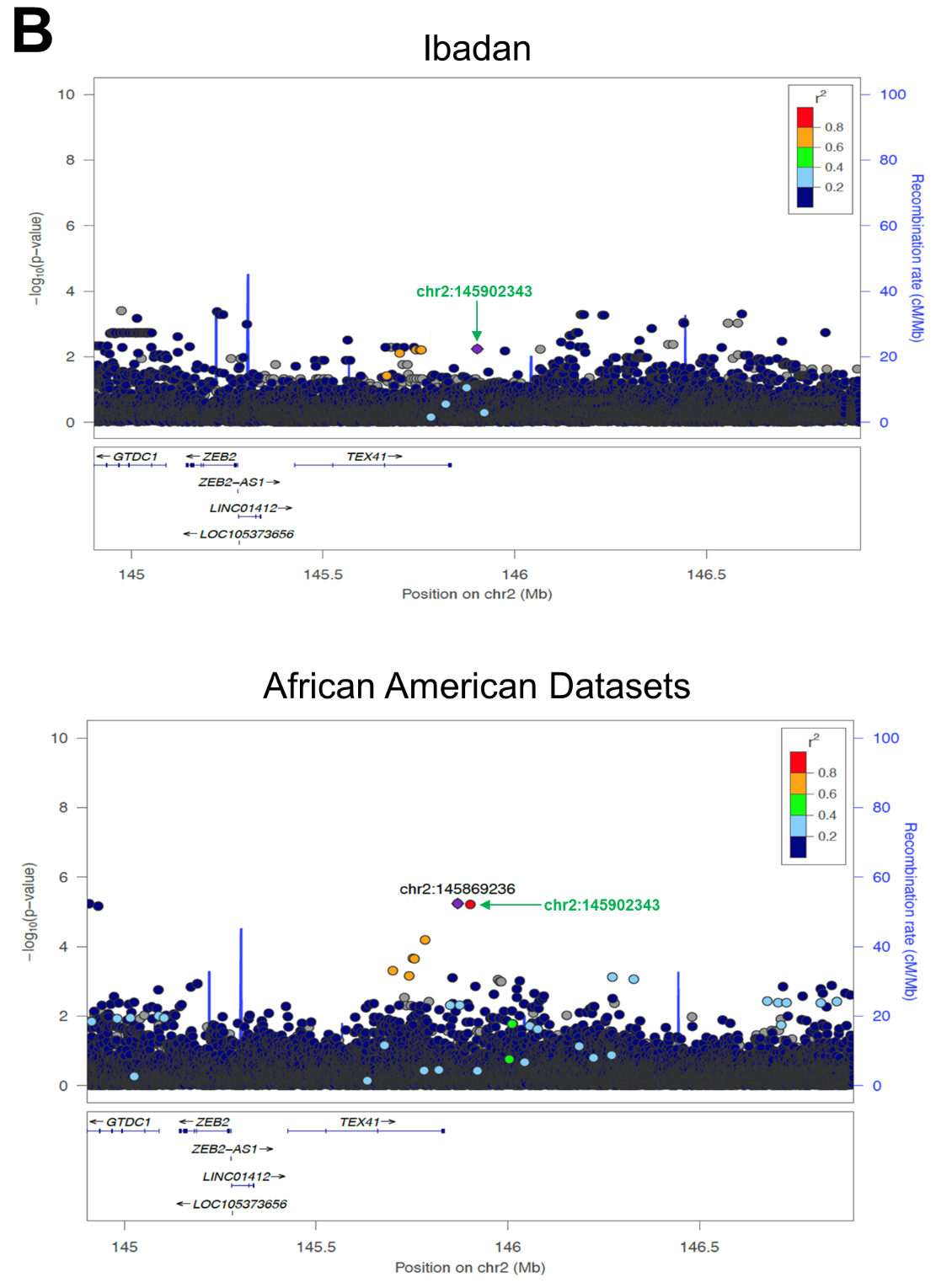

The top SNP reported in our results from the meta-analysis across all datasets (Ibadan and African American) is shown in green.

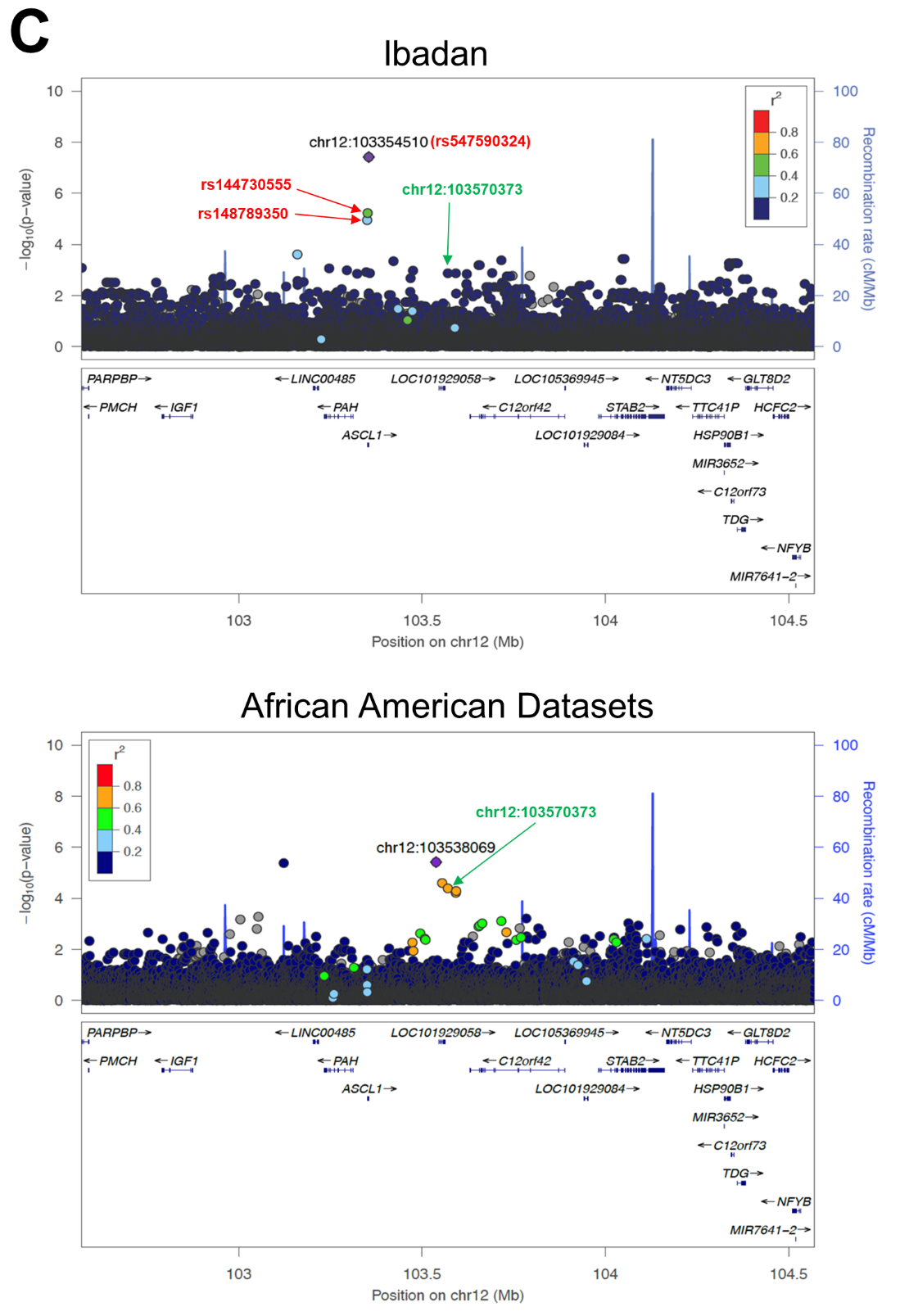

The top SNP reported in our results from the meta-analysis across all datasets (Ibadan and African American) is shown in green. The rsID numbers for the top three SNPs from the African ancestry dataset (Ibadan) are shown in red.

### Supplementary Figure 6. Manhattan plot of gene-based analysis results. Model 1 (a) is adjusted for age, sex and population stratification; Model 2 (b) is adjusted for age, sex, population stratification, and *APOE*.

A) Model 1

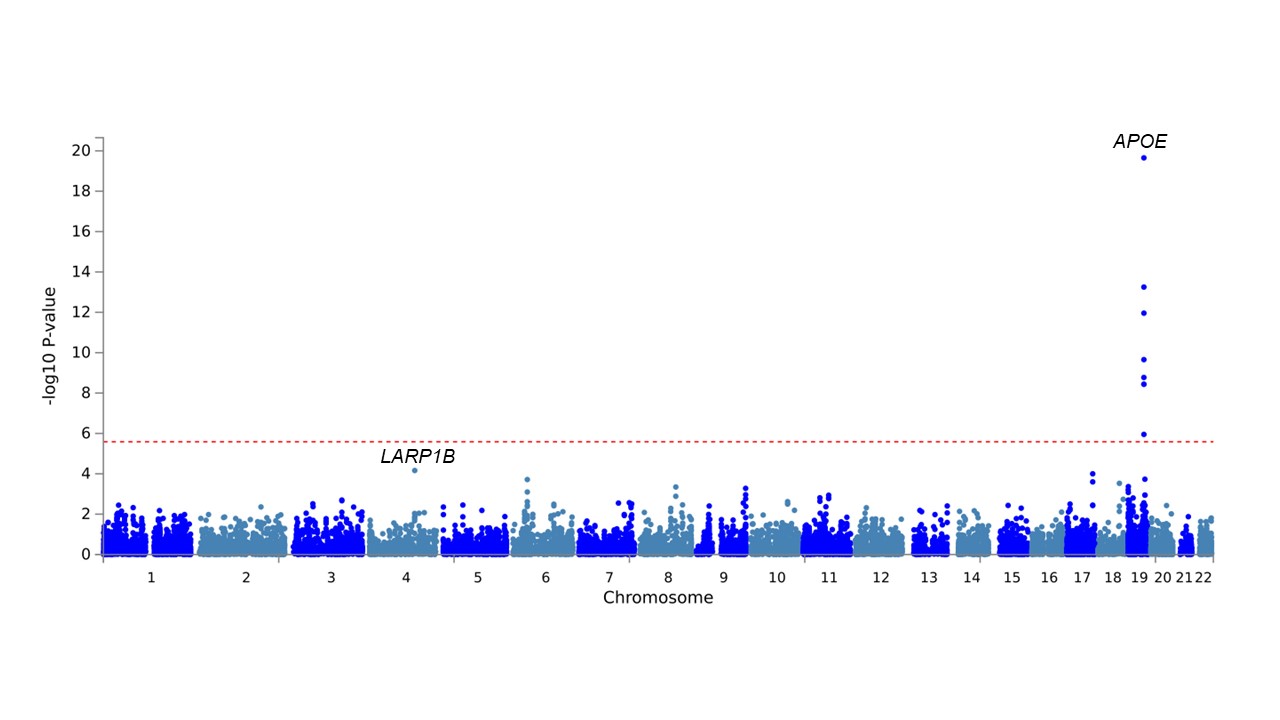

B) Model 2

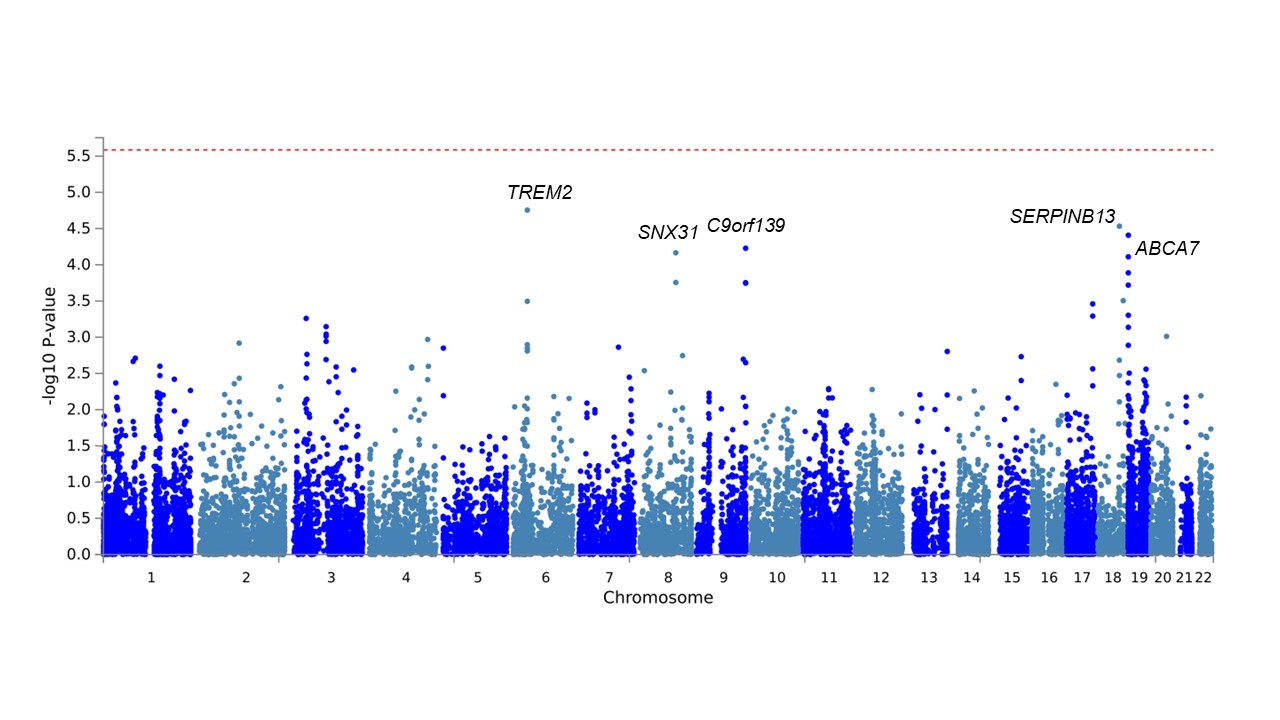

### Supplementary Figure 7. Quantile-quantile plots for gene-based analysis (A) on the model adjusted for age, sex and population stratification and (B) age, sex, population stratification and APOE showing the deviation of observed from expected p-values.

A) Model 1

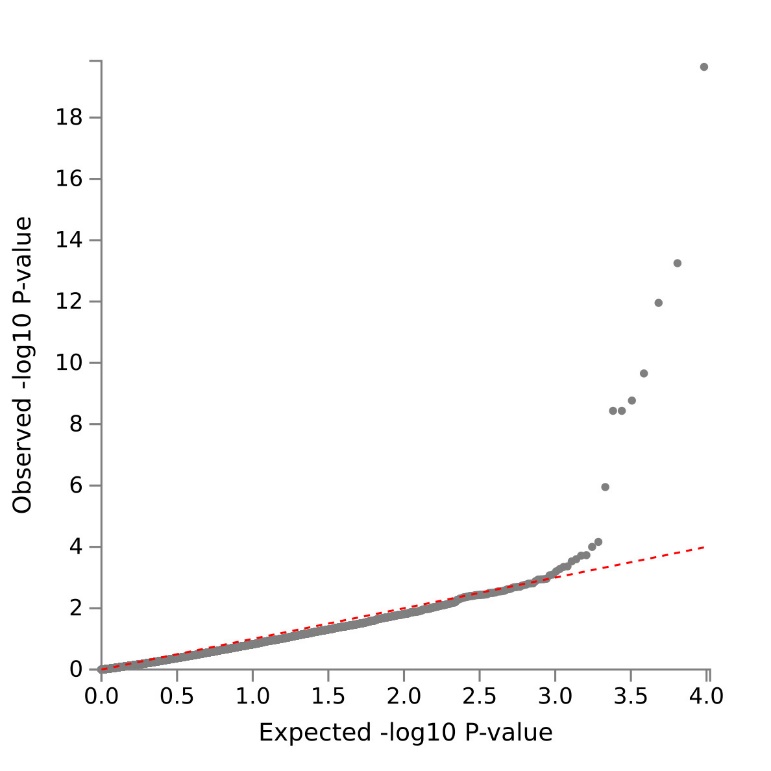

B) Model 2

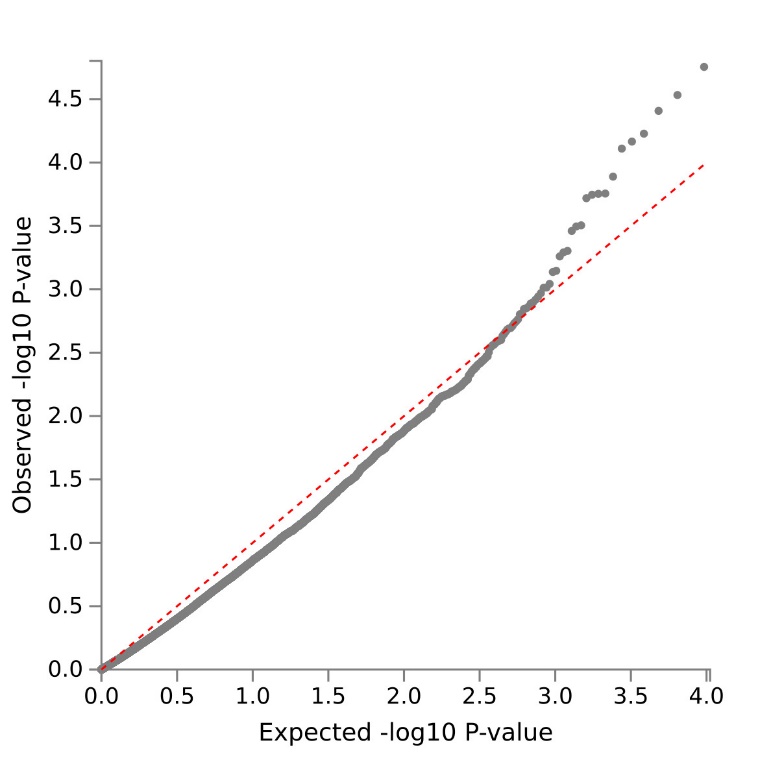

### Supplementary Figure 8. Differential brain gene expression between AD cases and controls of genes at top loci. Results are based on RNA-seq data from over 2100 samples from post-mortem brains of more than 1100 individuals from three human cohort studies (ROSMAP^38^, Mayo Clinic^39^, Mount Sinai Brain Bank (MSBB)^40^), and were generated from nine distinct brain regions (Anterior Cingulate Cortex (ACC), Dorsolateral Prefrontal Cortex (DLPFC), Posterior Cingulate Cortex (PCC), Cerebellum (CBE), Temporal Cortex (TCX), Frontal Pole (FP), Inferior Frontal Gyrus (IFG), Parahippocampal Gyrus (PHG), Superior Temporal Gyrus (STG)).

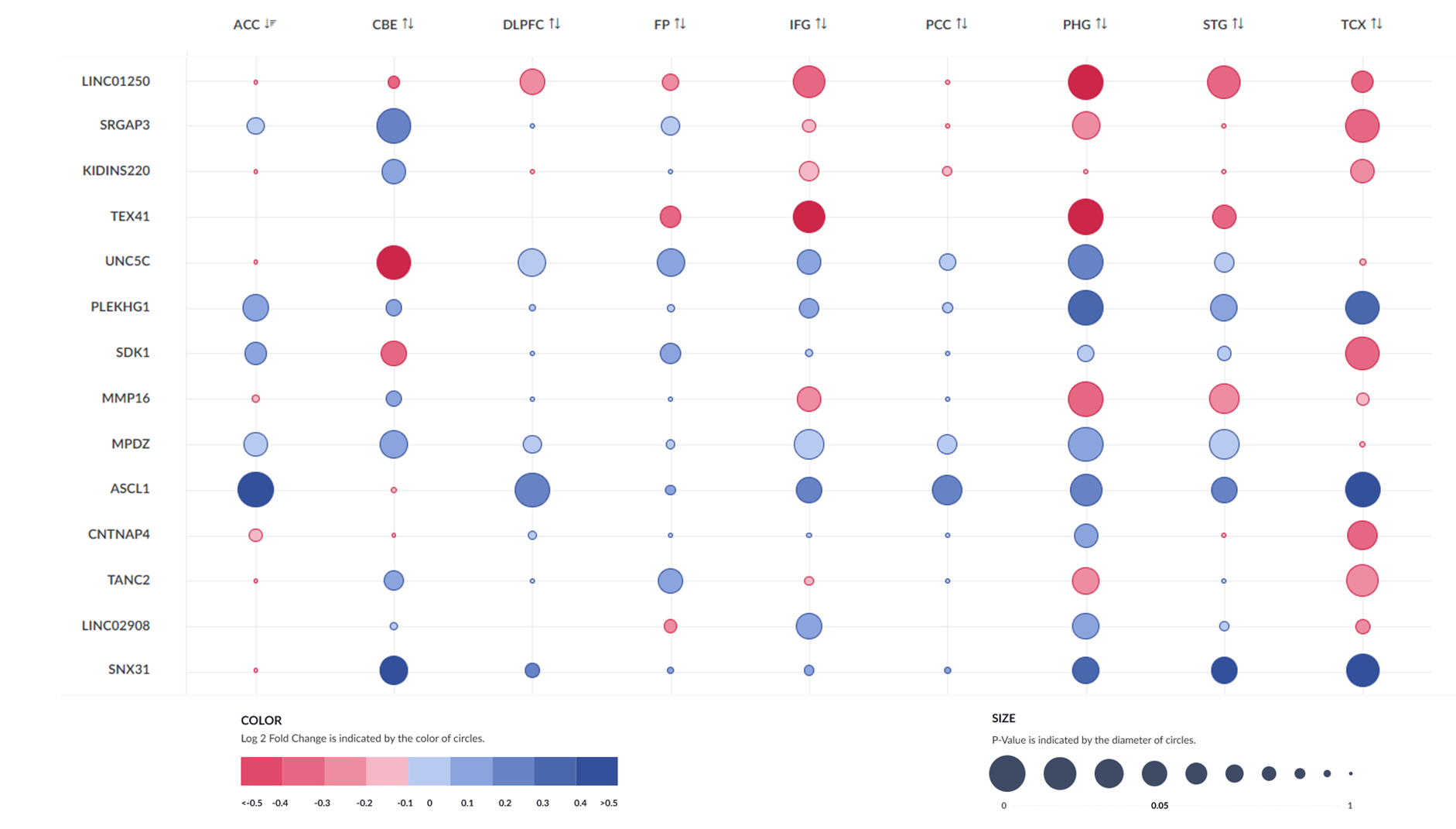

### Supplementary Figure 9. Results of bulk RNA sequence analysis in zebrafish brain (gene expression after amyloid toxicity compared to control).

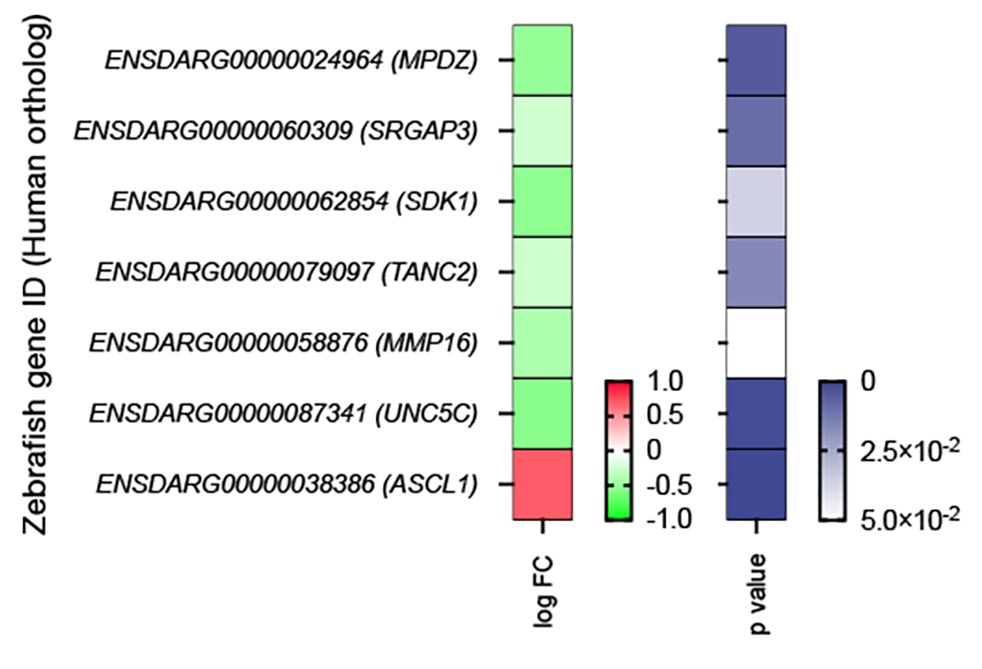

Differential gene expression for GWAS hits with identifiable zebrafish orthologs (green: downregulation, red: upregulation) and *P* values are shown. Zebrafish Ensembl IDs are given. Human gene names are shown in parentheses.

### Supplementary Figure 10. *MPDZ* splicing variants from GTEx human brain data^41^. Our top GWAS hit is shown here in red (chr9:13220518; rs141610415).

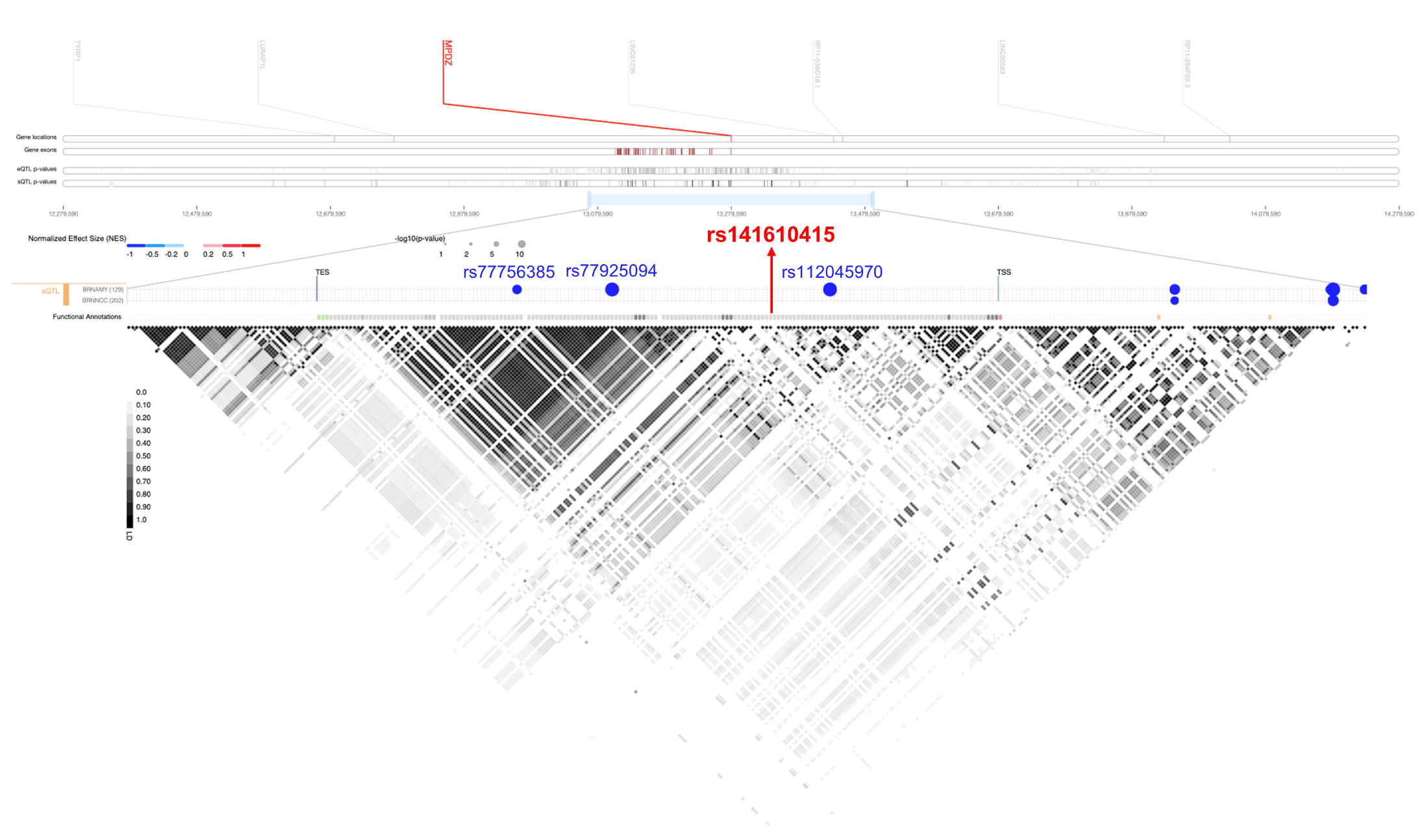

### Alzheimer’s Disease Genetics Consortium (ADGC) members

Erin Abner^1^, Perrie M. Adams^2^, Alyssa Aguirre^3^, Marilyn S. Albert^4^, Roger L. Albin^5,6,7^, Mariet Allen^8^, Lisa Alvarez^9^, Lia G. Apostolova^10,11,12,13^, Steven E. Arnold^14^, Sanjay Astha^15,16,17^, Craig S. Atwood^16,17,18^, Gayle Ayres^3^, Robert C. Barber^9^, Lisa L. Barnes^19,20,21^, Sandra Barral^22,23,24^, Jackie Bartlett^25^, Thomas G. Beach^26^, James T. Becker^27^, Gary W. Beecham^28,29^, Penelope Benchek^25^, David A. Bennett^19,21^, John Bertelson^30^, Sarah A. Biber^31^, Thomas D. Bird^32,33^, Deborah Blacker^34,35^, Bradley F. Boeve^36^, James D. Bowen^37^, Adam Boxer^38^, James B. Brewer^39^, James R. Burke^40^, Jeffrey M. Burns^41^, William S. Bush^25^, Joseph D. Buxbaum^42,43,44^, Goldie Byrd^45^, Laura B. Cantwell^46^, Chuanhai Cao^47^, Cynthia M. Carlsson^15,16,17^, Minerva M. Carrasquillo^8^, Kwun C. Chan^31^, Scott Chasse^48^, Yen-Chi Chen^31^, Marie-Francoise Chesselet^49^, thaniel A. Chin^15,16^, Hele C. Chui^50^, Jaeyoon Chung Jaeyoon Chung^51^, Suzanne Craft^52^, Paul K. Crane^53^, Carlos Cruchaga^54^, Michael L. Cuccaro^28,29,55^, Jessica Culhane^31^, C. Cullum^2,56,57^, Eveleen Darby^58^, Barbara Davis^57^, Charles DeCarli^59^, Vivian M. Deerlin^60^, John C. DeToledo^61^, Dennis W. Dickson^8^, Nic Dobbins^31^, Ranjan Duara^62^, Linda J. Eldik^63^, Nilufer Ertekin-Taner^8,64^, Denis A. Evans^65^, Kelley M. Faber^11^, Thomas J. Fairchild^66^, Daniele Fallin^67^, Kenneth B. Fallon^68^, David W. Fardo^69^, Martin R. Farlow^13^, John Farrell^51^, Lindsay A. Farrer^70,71,51,72,73^, Victoria Ferndez-Herndez^54^, Tatia M. Foroud^11^, Matthew P. Frosch^74^, Douglas R. Galasko^39^, Adria Gamboa^75^, Peter George-Hyslop^76,77^, Daniel H. Geschwind^49^, Berrdino Ghetti^78^, Alison M. Goate^42^, Thomas Grabowski^32,79^, Neill R. Graff-Radford^8,64^, Anthony R. Griswold^28^, Jothan L. Haines^25^, Hakon Hakorson^80^, Kathleen Hall^11^, James R. Hall^9^, Rold L. Hamilton^81^, Kara L. Hamilton-Nelson^28^, Xudong Han^51^, John Hardy^82^, Lindy E. Harrell^83^, Elizabeth Head^84^, Victor Henderson^85,86^, Michelle Herndez^61^, Lawrence S. Honig^22^, Ryan M. Huebinger^56^, Matthew J. Huentelman^87^, Christine M. Hulette^88^, Bradley T. Hyman^14^, Linda Hyn^57^, Laura Ibanez^54^, Philip L. Jager^89,90^, Gail P. Jarvik^91,92^, Suman Jayadev^32^, Lee-Way Jin^93^, Kimberly Johnson^61^, Leigh Johnson^9^, Gyungah Jun^70,73,94^, M. Kamboh^95,96^, Moon Il Kang^51^, An Karydas^38^, Gauthreaux M. Kathryn^31^, Mindy J. Katz^97^, John S.K. Kauwe^98^, Jeffrey A. Kaye^99,100^, C. Keene^101^, Benjamin Keller^31^, Aisha Khaleeq^58^, Rold Kim^84^, Janice Knebl^75^, Neil W. Kowall^72,102^, Joel H. Kramer^103^, Walter A. Kukull^31^, Brian W. Kunkle^28^, Amanda P. Kuzma^46^, Frank M. LaFerla^104^, James J. Lah^105^, Eric B. Larson^53,106^, Melissa Lerch^31^, Alan J. Lerner^25^, Yuk Leung^46^, James B. Leverenz^107^, Allan I. Levey^105^, Donghe Li^51^, Andrew P. Lieberman^108^, Richard B. Lipton^97^, Oscar L. Lopez^96^, Kathryn L. Lunetta^70^, Constantine G. Lyketsos^109^, Douglas Mains^110,111^, Jennifer Manly^22^, Logue Mark^51,112,113^, David Marquez^19^, Daniel C. Marson^83^, Eden R. Martin^28,29^, Eliezer Masliah^39,114^, Paul Massman^58^, Arjun V. Masurkar^115^, Richard Mayeux^22,23^, Wayne C. McCormick^53^, Susan M. McCurry^116^, Stefan McDonough^117^, Ann C. McKee^72,102^, Marsel Mesulam^118,119^, Jesse Mez^72^, Bruce L. Miller^38^, Carol A. Miller^50^, Charles Mock^31^, Abhay Moghekar^109^, Thomas J. Montine^120^, Edwin Monuki^84,121^, Sean D. Mooney^31^, John C. Morris^122,123^, Shubhabrata Mukherjee^53^, Amanda J. Myers^55^, Adam C. j^46^, Trung Nguyen^57^, Sid E. O’Bryant^75^, Kyle Ormsby^31^, Marcia Ory^124^, Raymond Palmer^125^, Joseph E. Parisi^126^, Henry L. Paulson^127^, Valory Pavlik^58^, David Paydarfar^3^, Victoria Perez^61^, Margaret A. Pericak-Vance^28,29^, Rold C. Petersen^36^, Marsha Polk^125^, Liming Qu^46^, Mary Quiceno^2^, Joseph F. Quinn^99,100^, Ashok Raj^47^, Farid Rajabli^28^, Vijay Raman^36^, Eric M. Reiman^87,128,129,130^, Joan S. Reisch^57^, Christiane Reitz^22,23,24,131^, John M. Ringman^50^, Erik D. Roberson^83^, Monica Rodriguear^58^, Ekateri Rogaeva^76^, Howard J. Rosen^38^, Roger N. Rosenberg^132^, Dold R. Royall^133^, Mary Sano^43^, Andrew J. Saykin^10,11^, Gerard D. Schellenberg^46^, Julie A. Schneider^19,21,134^, Lon S. Schneider^50,135^, William W. Seeley^38^, Richard M. Sherva^51^, Dean K. Shibata^31^, Scott Small^22,24^, Amanda G. Smith^47^, Janet Smith^57^, Yeunjoo Song^25^, Salvatore Spi^38^, Robert A. Stern^72^, Alan Stevens^124^, Stephen Strittmatter^136^, David Sultzer^137^, Russell H. Swerdlow^41^, Jeffrey Tilson^48^, Giuseppe Tosto^22,24^, John Q. Trojanowski^60^, Juan C. Troncoso^138^, Debby W. Tsuang^33,139^, Otto Valladares^46^, Jeffery M. Vance^28^, Badri N. Vardarajan^22,23,24^, Robert Vassar^140,119^, Harry V. Vinters^141,142^, Jean Vonsattel^143^, Li-San Wang^46^, Sandra Weintraub^118,140^, Kathleen A. Welsh-Bohmer^40,144,145^, Nick Wheeler^25^, Ellen Wijsman^91,92,146^, Kirk C. Wilhelmsen^48^, Scott Williams^25^, Benjamin Williams^57^, Jennifer Williamson^22^, Henrick Wilms^61^, Thomas S. Wingo^105^, Thomas Wisniewski^145,115^, Randall L. Woltjer^147^, Martin Woon^30^, Steven G. Younkin^8^, Lei Yu^19,21^, Yi Zhao^46^, Xiongwei Zhou^25^, Congcong Zhu^51^

^1^Sanders-Brown Center on Aging, College of Public Health, Department of Epidemiology, University of Kentucky, Lexington, Kentucky, ^2^Department of Psychiatry, University of Texas Southwestern Medical Center, Dallas, Texas, ^3^Department of Neurology, University of Texas at Austin/Dell Medical School, Austin, Texas, ^4^Department of Neurology, Johns Hopkins University, Baltimore, Maryland, ^5^Department of Neurology, University of Michigan, Ann Arbor, Michigan, ^6^Geriatric Research, Education and Clinical Center (GRECC), VA Ann Arbor Healthcare System (VAAAHS), Ann Arbor, Michigan, ^7^Michigan Alzheimer Disease Center, Ann Arbor, Michigan, ^8^Department of Neuroscience, Mayo Clinic, Jacksonville, Florida, ^9^Department of Pharmacology and Neuroscience, University of North Texas Health Science Center, Fort Worth, Texas, ^10^Department of Radiology, India University, Indiapolis, India, ^11^Department of Medical and Molecular Genetics, India University, Indiapolis, India, ^12^India Alzheimer's Disease Center, India University, Indiapolis, India, ^13^Department of Neurology, India University, Indiapolis, India, ^14^Department of Neurology, Massachusetts General Hospital/Harvard Medical School, Boston, Massachusetts, ^15^Geriatric Research, Education and Clinical Center (GRECC), University of Wisconsin, Madison, Wisconsin, ^16^Department of Medicine, University of Wisconsin, Madison, Wisconsin, ^17^Wisconsin Alzheimer's Disease Research Center, Madison, Wisconsin, ^18^Wisconsin Alzheimer's Disease Research Center, ^19^Department of Neurological Sciences, Rush University Medical Center, Chicago, Illinois, ^20^Department of Behavioral Sciences, Rush University Medical Center, Chicago, Illinois, ^21^Rush Alzheimer's Disease Center, Rush University Medical Center, Chicago, Illinois, ^22^Taub Institute on Alzheimer's Disease and the Aging Brain, Department of Neurology, Columbia University, New York, New York, ^23^Gertrude H. Sergievsky Center, Columbia University, New York, New York, ^24^Department of Neurology, Columbia University, New York, New York, ^25^Department of Epidemiology and Biostatistics, Case Western Reserve University, Cleveland, Ohio, ^26^Civin Laboratory for Neuropathology, Banner Sun Health Research Institute, Phoenix, Arizo, ^27^Departments of Psychiatry, Neurology, and Psychology, University of Pittsburgh School of Medicine, Pittsburgh, Pennsylvania, ^28^The John P. Hussman Institute for Human Genomics, University of Miami, Miami, Florida, ^29^Dr. John T. Macdold Foundation Department of Human Genetics, University of Miami, Miami, Florida, ^30^Department of Psychiatry, University of Texas at Austin/Dell Medical School, Austin, Texas, ^31^Department of Epidemiology, University of Washington, Seattle, Washington, ^32^Department of Neurology, University of Washington, Seattle, Washington, ^33^VA Puget Sound Health Care System/GRECC, Seattle, Washington, ^34^Department of Epidemiology, Harvard School of Public Health, Boston, Massachusetts, ^35^Department of Psychiatry, Massachusetts General Hospital/Harvard Medical School, Boston, Massachusetts, ^36^Department of Neurology, Mayo Clinic, Rochester, Minnesota, ^37^Swedish Medical Center, Seattle, Washington, ^38^Department of Neurology, University of California San Francisco, San Francisco, California, ^39^Department of Neurosciences, University of California San Diego, La Jolla, California, ^40^Department of Medicine, Duke University, Durham, North Caroli, ^41^University of Kansas Alzheimer's Disease Center, University of Kansas Medical Center, Kansas City, Kansas, ^42^Department of Neuroscience, Mount Sii School of Medicine, New York, New York, ^43^Department of Psychiatry, Mount Sii School of Medicine, New York, New York, ^44^Departments of Genetics and Genomic Sciences, Mount Sii School of Medicine, New York, New York, ^45^Social Sciences & Health Policy, Wake Forest School of Medicine, Winston-Salem, North Caroli, ^46^Penn Neurodegeneration Genomics Center, Department of Pathology and Laboratory Medicine, University of Pennsylvania Perelman School of Medicine, Philadelphia, Pennsylvania, ^47^USF Health Byrd Alzheimer's Institute, University of South Florida, Tampa, Florida, ^48^Department of Genetics, University of North Caroli Chapel Hill, Chapel Hill, North Caroli, ^49^Neurogenetics Program, University of California Los Angeles, Los Angeles, California, ^50^Department of Neurology, University of Southern California, Los Angeles, California, ^51^Department of Medicine (Biomedical Genetics), Boston University, Boston, Massachusetts, ^52^Gerontology and Geriatric Medicine Center on Diabetes, Obesity, and Metabolism, Wake Forest School of Medicine, Winston-Salem, North Caroli, ^53^Department of Medicine, University of Washington, Seattle, Washington, ^54^Department of Psychiatry and Hope Center Program on Protein Aggregation and Neurodegeneration, Washington University School of Medicine, St. Louis, Missouri, ^55^Department of Psychiatry and Behavioral Sciences, Miller School of Medicine, University of Miami, Miami, Florida, ^56^Department of Surgery, University of Texas Southwestern Medical Center, Dallas, Texas, ^57^Department of Clinical Sciences, University of Texas Southwestern Medical Center, Dallas, Texas, ^58^Alzheimer's Disease and Memory Disorders Center, Baylor College of Medicine, Houston, Texas, ^59^Department of Neurology, University of California Davis, Sacramento, California, ^60^Department of Pathology and Laboratory Medicine, University of Pennsylvania Perelman School of Medicine, Philadelphia, Pennsylvania, ^61^Departments of Neurology, Pharmacology & Neuroscience, Texas Tech University Health Science Center, Lubbock, Texas, ^62^Wien Center for Alzheimer's Disease and Memory Disorders, Mount Sii Medical Center, Miami Beach, Florida, ^63^Sanders-Brown Center on Aging, Department of Atomy and Neurobiology, University of Kentucky, Lexington, Kentucky, ^64^Department of Neurology, Mayo Clinic, Jacksonville, Florida, ^65^Rush Institute for Healthy Aging, Department of Interl Medicine, Rush University Medical Center, Chicago, Illinois, ^66^Office of Strategy and Measurement, University of North Texas Health Science Center, Fort Worth, Texas, ^67^Department of Public Health, Rollins School of Public Health, Johns Hopkins University, Baltimore, Maryland, ^68^Department of Pathology, University of Alabama at Birmingham, Birmingham, Alabama, ^69^Sanders-Brown Center on Aging, Department of Biostatistics, University of Kentucky, Lexington, Kentucky, ^70^Department of Biostatistics, Boston University, Boston, Massachusetts, ^71^Department of Epidemiology, Boston University, Boston, Massachusetts, ^72^Department of Neurology, Boston University, Boston, Massachusetts, ^73^Department of Ophthalmology, Boston University, Boston, Massachusetts, ^74^C.S. Kubik Laboratory for Neuropathology, Massachusetts General Hospital, Charlestown, Massachusetts, ^75^Interl Medicine, Division of Geriatrics, University of North Texas Health Science Center, Fort Worth, Texas, ^76^Tanz Centre for Research in Neurodegenerative Disease, University of Toronto, Toronto, Ontario, ^77^Cambridge Institute for Medical Research and Department of Clinical Neurosciences, University of Cambridge, Cambridge, United Kingdom, ^78^Department of Pathology and Laboratory Medicine, India University, Indiapolis, India, ^79^Department of Radiology, University of Washington, Seattle, Washington, ^80^Center for Applied Genomics, Children's Hospital of Philadelphia, Philadelphia, Pennsylvania, ^81^Department of Pathology (Neuropathology), University of Pittsburgh, Pittsburgh, Pennsylvania, ^82^Institute of Neurology, University College London, Queen Square, London, United Kingdom, ^83^Department of Neurology, University of Alabama at Birmingham, Birmingham, Alabama, ^84^Department of Pathology and Laboratory Medicine, University of California Irvine, Irvine, California, ^85^Department of Epidemiology and Population Health, Stanford University, Stanford, California, ^86^Department of Neurology & Neurological Sciences, Stanford University, Stanford, California, ^87^Neurogenomics Division, Translatiol Genomics Research Institute, Phoenix, Arizo, ^88^Department of Pathology, Duke University, Durham, North Caroli, ^89^Program in Translatiol Neuro-Psychiatric Genomics, Institute for the Neurosciences, Department of Neurology & Psychiatry, Brigham and Women's Hospital and Harvard Medical School, Boston, Massachusetts, ^90^Program in Medical and Population Genetics, Broad Institute, Cambridge, Massachusetts, ^91^Department of Genome Sciences, University of Washington, Seattle, Washington, ^92^Department of Medicine (Medical Genetics), University of Washington, Seattle, Washington, ^93^Department of Pathology and Laboratory Medicine, University of California Davis, Sacramento, California, ^94^Department of Medicine (Genetics Program), Boston University, Boston, Massachusetts, ^95^Department of Human Genetics, University of Pittsburgh, Pittsburgh, Pennsylvania, ^96^University of Pittsburgh Alzheimer's Disease Research Center, Pittsburgh, Pennsylvania, ^97^Department of Neurology, Albert Einstein College of Medicine, New York, New York, ^98^Department of Biology, Brigham Young University, Provo, Utah, ^99^Department of Neurology, Oregon Health & Science University, Portland, Oregon, ^100^Department of Neurology, Portland Veterans Affairs Medical Center, Portland, Oregon, ^101^Department of Pathology, University of Washington, Seattle, Washington, ^102^Department of Pathology, Boston University, Boston, Massachusetts, ^103^Department of Neuropsychology, University of California San Francisco, San Francisco, California, ^104^Department of Neurobiology and Behavior, University of California Irvine, Irvine, California, ^105^Department of Neurology, Emory University, Atlanta, Georgia, ^106^Group Health Research Institute, Group Health, Seattle, Washington, ^107^Cleveland Clinic Lou Ruvo Center for Brain Health, Cleveland Clinic, Cleveland, Ohio, ^108^Department of Pathology, University of Michigan, Ann Arbor, Michigan, ^109^Department of Psychiatry, Johns Hopkins University, Baltimore, Maryland, ^110^Department of Health Behavior and Health Systems, University of North Texas Health Science Center, Fort Worth, Texas, ^111^Health Magement and Policy Department, School of Public Health, University of North Texas Health Science Center, Fort Worth, Texas, ^112^tiol Center for PTSD at Boston VA Healthcare System, Boston, Massachusetts, ^113^Department of Psychiatry, Boston University School of Medicine, Boston, Massachusetts, ^114^Department of Pathology, University of California San Diego, La Jolla, California, ^115^Department of Psychiatry, New York University, New York, New York, ^116^School of Nursing Northwest Research Group on Aging, University of Washington, Seattle, Washington, ^117^PharmaTherapeutics Clinical Research, Pfizer Worldwide Research and Development, Cambridge, Massachusetts, ^118^Department of Pathology, Northwestern University Feinberg School of Medicine, Chicago, Illinois, ^119^Department of Neurology, Northwestern University Feinberg School of Medicine, Chicago, Illinois, ^120^Department of Pathology, Stanford University School of Medicine, Stanford, California, USA., ^121^Department of Developmental and Cell Biology, UC Irvine, Irvine, ^122^Department of Neurology, Washington University, St. Louis, Missouri, ^123^Department of Pathology and Immunology, Washington University, St. Louis, Missouri, ^124^Center for Population Health & Aging, Texas A&M University Health Science Center, Lubbock Texas, ^125^Department of Family and Community Medicine, University of Texas Health Science Center - San Antonio, San Antonio, Texas, ^126^Department of Laboratory Medicine and Pathology, Mayo Clinic, Rochester, Minnesota, ^127^Michigan Alzheimer's Disease Center, Department of Neurology, University of Michigan, Ann Arbor, Michigan, ^128^Arizo Alzheimer's Consortium, Phoenix, Arizo, ^129^Banner Alzheimer's Institute, Phoenix, Arizo, ^130^Department of Psychiatry, University of Arizo, Phoenix, Arizo, ^131^Department of Epidemiology, Columbia University, New York, New York, ^132^Department of Neurology, University of Texas Southwestern, Dallas, Texas, ^133^Departments of Psychiatry, Medicine, Family & Community Medicine, South Texas Veterans Health Administration Geriatric Research Education & Clinical Center (GRECC), UT Health Science Center at San Antonio, San Antonio, Texas, ^134^Department of Pathology (Neuropathology), Rush University Medical Center, Chicago, Illinois, ^135^Department of Psychiatry, University of Southern California, Los Angeles, California, ^136^Program in Cellular Neuroscience, Neurodegeneration & Repair, Yale University, New Haven, Connecticut, ^137^Department of Psychiatry & Human Behavior, University of California Irvine, Irvine, California, ^138^Department of Pathology, Johns Hopkins University, Baltimore, Maryland, ^139^Department of Psychiatry and Behavioral Sciences, University of Washington School of Medicine, Seattle, Washington, ^140^Cognitive Neurology and Alzheimer's Disease Center, Northwestern University Feinberg School of Medicine, Chicago, Illinois, ^141^Department of Neurology, University of California Los Angeles, Los Angeles, California, ^142^Department of Pathology & Laboratory Medicine, University of California Los Angeles, Los Angeles, California, ^143^Taub Institute on Alzheimer's Disease and the Aging Brain, Department of Pathology, Columbia University, New York, New York, ^144^Department of Psychiatry & Behavioral Sciences, Duke University, Durham, North Caroli, ^145^Center for Cognitive Neurology and Departments of Neurology, New York University, School of Medicine, New York, ^146^Department of Biostatistics, University of Washington, Seattle, Washington, ^147^Department of Pathology, Oregon Health & Science University, Portland, Oregon
